# Modeling islet enhancers using deep learning identifies candidate causal variants at loci associated with T2D and glycemic traits

**DOI:** 10.1101/2022.05.13.22275035

**Authors:** Sanjarbek Hudaiberdiev, D. Leland Taylor, Wei Song, Narisu Narisu, Redwan M. Bhuiyan, Henry J. Taylor, Tingfen Yan, Amy J. Swift, Lori L. Bonnycastle, DIAMANTE Consortium, Michael L. Stitzel, Michael R. Erdos, Ivan Ovcharenko, Francis S. Collins

## Abstract

Genetic association studies have identified hundreds of independent signals associated with type 2 diabetes (T2D) and related traits. Despite these successes, the identification of specific causal variants underlying a genetic association signal remains challenging. In this study, we describe a deep learning method to analyze the impact of sequence variants on enhancers. Focusing on pancreatic islets, a T2D relevant tissue, we show that our model learns islet-specific transcription factor (TF) regulatory patterns and can be used to prioritize candidate causal variants. At 101 genetic signals associated with T2D and related glycemic traits where multiple variants occur in linkage disequilibrium, our method nominates a single causal variant for each association signal, including three variants previously shown to alter reporter activity in islet-relevant cell types. For another signal associated with blood glucose levels, we biochemically test all candidate causal variants from statistical fine-mapping using a pancreatic islet beta cell line and show biochemical evidence of allelic effects on TF binding for the model-prioritized variant. To aid in future research, we publicly distribute our model and islet enhancer perturbation scores across ∼67 million genetic variants. We anticipate that deep learning methods like the one presented in this study will enhance the prioritization of candidate causal variants for functional studies.

## Introduction

Over the past 20 years, immense progress has been made towards unraveling the genetic basis of diseases and traits, with tens of thousands of genetic associations identified to date (1). These associations could guide advances towards effective treatment and prevention of disease by shedding light on the underlying disease etiology and pinpointing specific genes, cell types, and molecular pathways that contribute to a disease. However, despite a few notable examples (2, 3), only modest progress has been made in translating genetic discoveries about common, complex diseases into therapies.

This challenge of translation is driven in part by the difficulties in (i) identifying which variants influence disease, since most disease-associated genetic signals are composed of many candidate causal SNPs due to linkage disequilibrium (LD), and (ii) establishing how these variants mechanistically function. To date, a variety of approaches (reviewed in (4)) have been developed to prioritize candidate causal SNPs by weighting SNPs according to their statistical evidence of association (statistical fine-mapping) and by the functional/epigenomic signals overlapping a SNP (functional fine-mapping). However, such methods often fail to nominate a feasible number of SNP candidates to test in the laboratory due to (i) limited statistical power to disentangle the effects of correlated SNPs even at large sample sizes and (ii) no clear metric to weight epigenomic overlaps in the case of functional fine-mapping.

Type 2 diabetes (T2D) is an exemplary case of the challenges of identifying causal variants and effector genes. T2D is a disease characterized by pancreatic islet beta cell dysfunction and insulin resistance in peripheral tissues (5). In a recent fine-mapping analysis of 898,130 European-descent participants, 243 loci were associated with T2D. These loci contained 403 distinct association signals (multiple, independent signals per locus), of which 18 signals could be narrowed down to one SNP based on statistical fine-mapping (6). Given the strong enrichment of T2D genetic signals in regulatory regions (e.g., enhancers) active in islets (reviewed in (7)), the authors performed functional fine-mapping of these 403 signals using islet epigenomic information and refined this list to 23 signals with one SNP, leaving much room for improvement.

In this study (overview in Fig. 1), we report a deep learning (DL) method that models both shared and tissue-specific genomic and epigenomic signals. Using this method, we analyze the impact of mutational profiles on pancreatic islet enhancers within the context of their local, surrounding DNA sequence. We show that our model learns islet-specific transcription factor (TF) regulatory patterns and can be used to refine fine-mapping results and prioritize candidate causal SNPs. By applying our model to prioritize SNPs from statistical fine-mapping results for T2D and related traits, we nominate a single candidate SNP that likely affects pancreatic islet enhancers at 101 signals containing more than one 99% credible set SNP from statistical fine-mapping. For three signals, previous studies validate our SNP predictions by showing these SNPs induce allelic activity in reporter assays in islet-relevant cell types. For another signal associated with blood glucose levels (near *PSMA1*), we biochemically demonstrate using a pancreatic islet beta cell line that the SNP prioritized by our model shows the greatest allelic effects on TF binding among all SNPs in the 99% credible set. We believe models like the one presented in this study will aid in refining candidate causal SNPs from fine-mapping for further functional studies across a wide variety of diseases/traits.

**Fig. 1.**
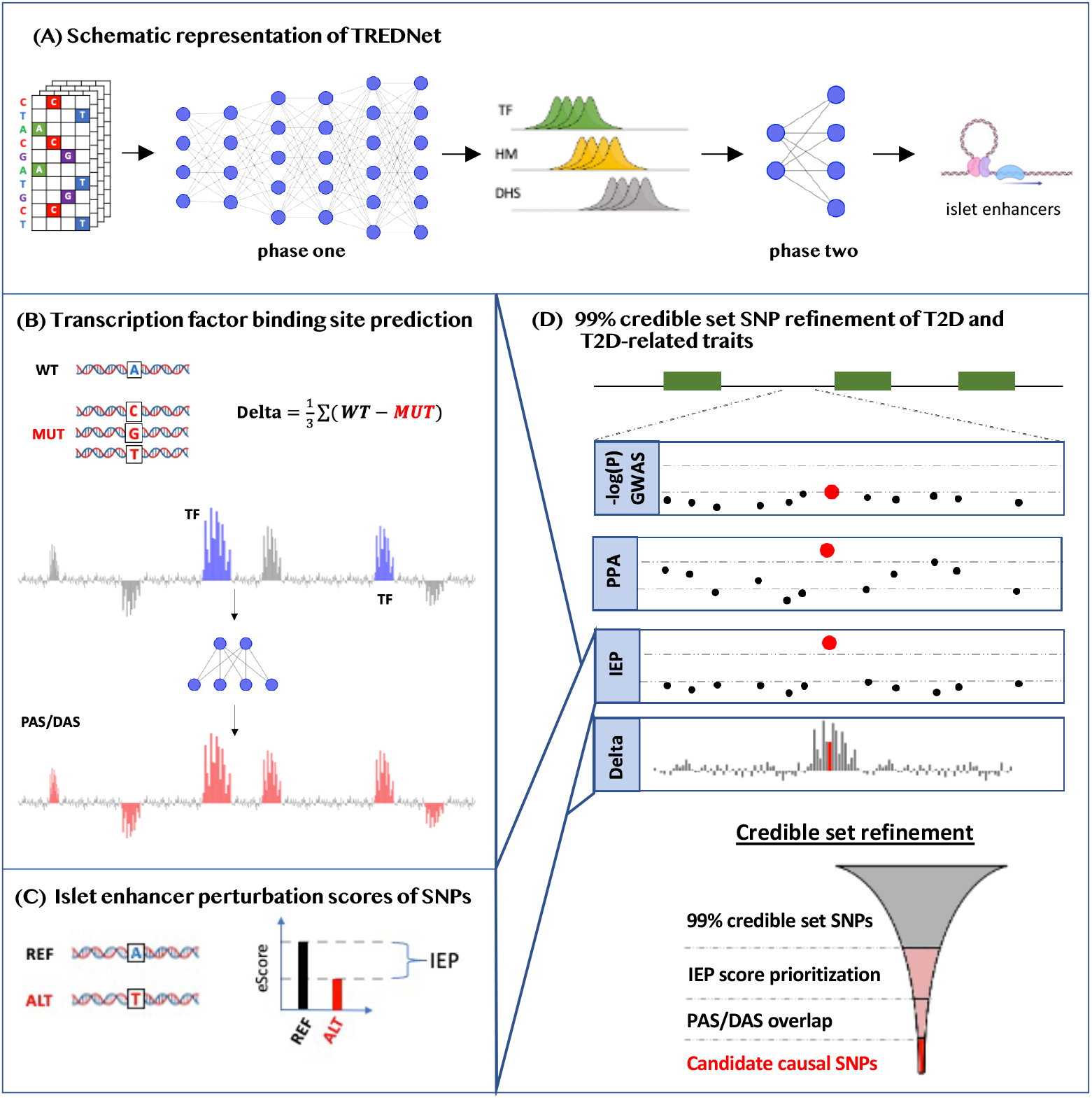
Graphical overview of this study. (A) Overview of TREDNet. TREDNet consists of two separate convolutional neural networks (CNNs; mesh of gray lines and blue circles). The first CNN is trained on genomic regions in one-hot encoded representation to predict peaks of epigenomic features, including transcription factors (TFs), histone modifications (HMs), and DNase I hypersensitivity sites (DHSs). The second CNN is trained on the output from the first CNN to predict enhancer regions. Enhancer graphic created with BioRender.com. (B) Saturated mutagenesis analysis using TREDNet produces delta scores, which are used to predict transcription factor binding sites (TFBSs), corresponding to peaks (peak active sites; PASs) and dips (dip active sites; DASs) in delta scores. Bars depict delta scores of each genomic position (x-axis). Blue bars show positions corresponding to known TFBSs. Red bars show TFBSs predicted by a CNN (mesh of gray lines and blue circles) using delta scores. (C) Allelic differences in TREDNet enhancer probability scores are used to calculate islet enhancer perturbation (IEP) scores for each SNP. (D) Schematic locus zoom example at a genetic signal where a candidate causal SNP is identified. Green boxes depict gene coding regions along the genome (x-axis). Subsequent facets show different signals for each SNP (points): the −*log*_10_(*P*) of the genetic association, the posterior probability of association (PPA) from statistical fine-mapping, IEP scores, and delta scores. Funnel schematic describes the framework used to identify candidate causal SNPs. SNPs from 99% credible sets are prioritized using IEP scores. Subsequently, SNPs are prioritized by PAS/DAS overlap.

## Results

### TREDNet: a deep learning model for enhancer prediction

To predict enhancers based on DNA sequence, we developed TREDNet, a two phase DL framework consisting of two consecutive convolutional neural networks (CNNs): the first to predict epigenomic signals across the genome and the second to predict enhancers (Methods; Fig. 1A).

For the phase one model, we trained a CNN that uses tiled DNA sequences of 2,000 base pairs (bp) to predict DNase I hypersensitive sites (DHSs), histone modifications (HMs), and TF binding sites (TFBSs) across 127 human cell types and tissues (biospecimens) from the ENCODE (8) and NIH Roadmap (9) studies (1,924 features in total). We excluded signals on chromosomes 8 and 9 from training and used them to test the model’s accuracy. We found that the phase one TREDNet model was highly accurate, achieving an average area under the receiver operating characteristic (auROC) of 0.93, 0.88, and 0.96 for DHSs, HMs, and TFBSs respectively (Fig. S1). We compared TREDNet’s phase one model to other methods that predict epigenomic signals from DNA sequences—ExPecto (10), DeepSEA (11), and Basset (12)—and found TREDNet performed similarly to these previous models (Fig. S1).

Using the vectors of 1,924 epigenomic predictions generated by the phase one model for each 2,000bp DNA sequence, we trained a second (phase two) CNN to predict pancreatic islet enhancers—defined using chromatin accessibility profiles (ATAC-seq peaks) and H3K27ac histone marks (Methods). We trained two similar models to predict HepG2 and K562 enhancers (one for each cell line) to validate our approach using datasets available for these two cell lines only. The result of this two phase learning framework is the enhancer probability of a 2,000bp DNA sequence. We used enhancer coordinates from chromosomes 8 and 9 withheld from training for validation and found that the phase two TREDNet model achieved an auROC of 0.92, 0.89, and 0.85 for islets, HepG2, and K562, respectively (Fig. 2A). As a benchmark, we compared TREDNet to other models that predict enhancers: BiRen (13), Tan et al. (14), and SVM (15). We found that the phase two TREDNet model outperformed the other models consistently (Fig. 2A).

**Fig. 2.**
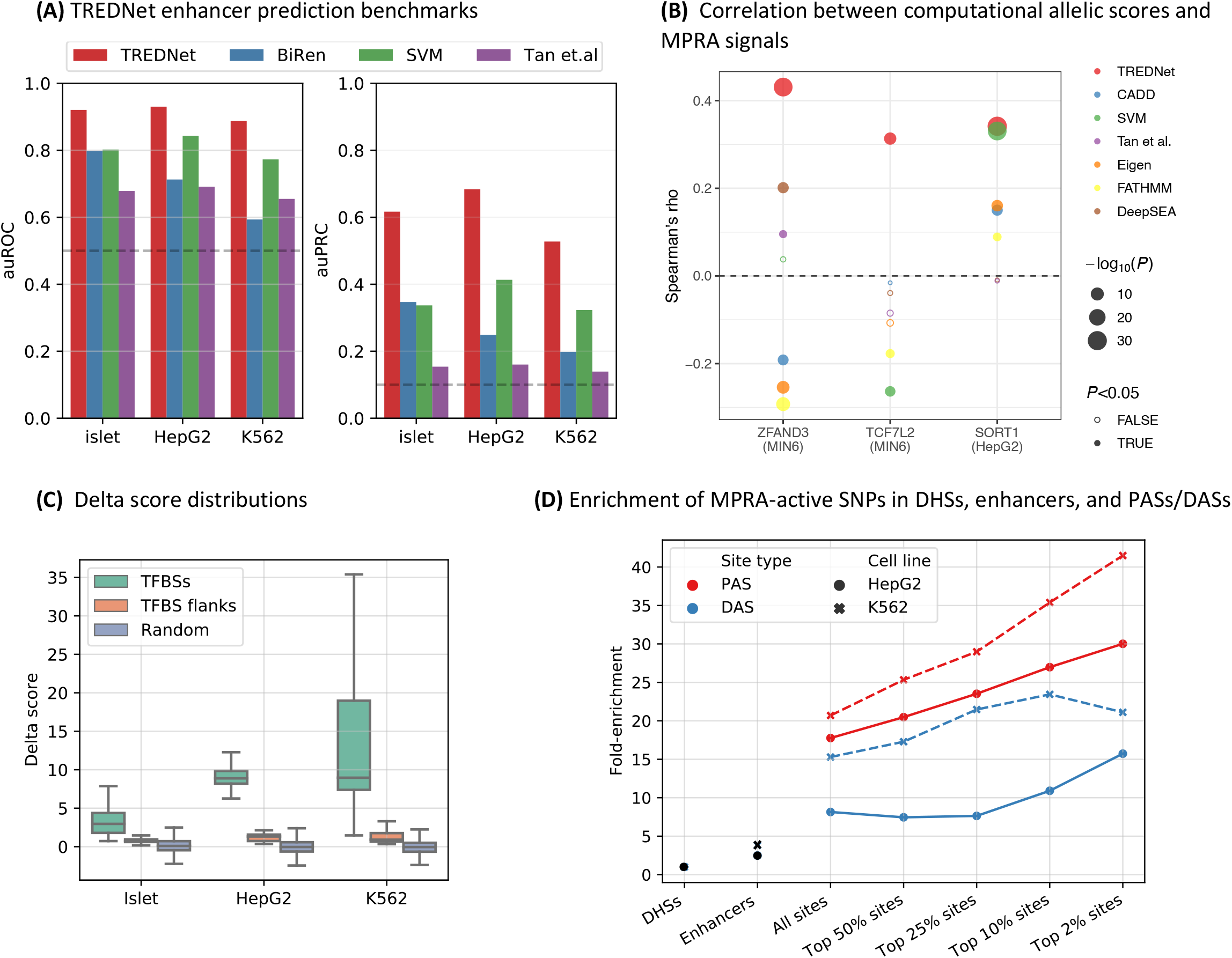
Characterization of TREDNet, TREDNet delta scores, and PASs/DASs. (A) Phase two TREDNet enhancer prediction accuracy across biospecimens (x-axis) compared to other models (colors) using area under the receiver operating characteristic (auROC; left) and area under the precision recall curve (auPRC; right) metrics (y-axis). Dashed horizontal lines show the performance of a random classifier: auROC=0.5 and auPRC=0.09. (B) Correlation (Spearman’s rho; y-axis) between allelic scores derived from computational methods (colors) and MPRA signals at different enhancers in biospecimens (x-axis). (C) Distribution of TREDNet delta scores (y-axis) in TFBSs, TFBS flanking regions, and random genomic regions outside of TFBSs (colors) across biospecimens (x-axis). (D) Enrichment (y-axis) of MPRA-active SNPs from HepG2 and K562 experiments (point shape and linetype) in PASs/DASs (colors), enhancers, and DHSs (x-axis). PASs/DASs are binned into five groups by their average delta score.

To further validate TREDNet’s enhancer probability scores, we used massively parallel reporter assay (MPRA) experiments that tested the allelic effects of mutations in enhancer regions of MIN6, a mouse beta cell line, and HepG2 (16). At each sequence position tested, we compared the allelic differences of the *in vitro* gene expression MPRA results to the allelic differences of the enhancer probabilities generated by TREDNet (Methods). We observed a strong, positive correlation between TREDNet’s predictions and the measured MPRA effects (minimum rho=0.31, *P*<1×10^−8^ across all enhancer regions, Spearman’s rank-order correlation; Fig. 2B). To benchmark TREDNet, we compared the MPRA allelic signals to allelic scores from other computational models (Methods), including DeepSEA (11), CADD (17), Eigen (18), FATHMM (19), Tan et al. (14), and SVM (15). We found that TREDNet consistently performed better than these alternative methods (Fig. 2B).

### *In silico* saturated mutagenesis of enhancers reveals TF regulatory patterns

In order to probe the regulatory structure of enhancers, we performed an *in silico* saturated mutagenesis experiment across the DNA sequences of all islet, HepG2, and K562 enhancers, predicting the effects of nucleotide mutations on the overall enhancer probability of the surrounding 2,000bp region (Methods). For every DNA sequence position within an enhancer, we calculated TREDNet delta scores, defined as the average difference in the enhancer probability of the reference nucleotide (GRCh37) and the enhancer probabilities of all other possible nucleotides. A positive delta score indicates a negative change in enhancer probability (enhancer-damaging), while a negative delta score indicates a positive change in enhancer probability (enhancer-strengthening).

Next, we asked if delta scores mark TFBSs, as has been suggested by previous studies that used similar metrics (reviewed in (20)). Using TFBSs from HepG2 ChIP-seq experiments (8), K562 ChIP-seq experiments (8), and islet ATAC-seq footprints (21) (since islets do not have as comprehensive of TF ChIP-seq profiles like HepG2 and K562), we compared the absolute value of delta scores within TFBSs to 20bp regions immediately flanking each TFBS as well as randomly sampled enhancer regions (Methods). We found that the delta scores of regions within TFBSs were much greater than flanking regions (an average 7-fold increase; *P*<1×10^−100^, Wilcoxon rank sum test) or randomly sampled regions (an average 35-fold increase; *P*<1×10^−100^, Wilcoxon rank sum test; Fig. 2C)—confirming that elevated delta scores differentiate TFBSs.

To explore if the TFBSs identified by delta scores are relevant to a tissue/cell type, we focused on islets and ranked each TF footprint by the ratio of the average delta score within the TF footprint motif to the flanking region (Table S1). The top five TFs were all known to play an important regulatory role in islets: TCF7L2 (22), the FOX family of TFs (23, 24), the C/EBP family of TFs (25), the HNF family of TFs (26), and DBP (27). Moreover, across all islet TFBSs, we found that the delta scores were strongly correlated with the per-nucleotide information content of each TFBS motif (average Spearman’s rho=0.45)—much more than the evolutionary sequence conservation (average Spearman’s rho=0.20; *P*=1.8×10^−14^, Wilcoxon rank sum test; Fig. S2; Methods). These trends held true for both HepG2 and K562 (Fig. S2B). Combined, these results strongly suggest that TFBSs with the largest delta scores demarcate TFBSs important to a cell type and thereby allow one to predict regions within enhancers with the greatest effect when altered in the relevant cell type.

Given the observed link between delta scores and TFBSs, we designed another DL model to predict TFBSs directly from delta score profiles (Methods). For each biospecimen, we trained two separate models to predict TFBSs from either ChIP-seq experiments (HepG2 and K562) or ATAC-seq footprints (islets) as short stretches (>=3bp; average length 13.7bp) of either enhancer-damaging regions (peak active sites; PASs) or enhancer-strengthening regions (dip active sites; DASs; Fig. S3). The resulting models were highly accurate at predicting TFBSs on chromosomes 8 and 9 (excluded from training), achieving an average auROC of 0.92 for PASs and 0.84 for DASs (Fig. S4). To further validate the PAS/DAS predictions, we calculated the enrichment of SNPs shown to have regulatory activity in HepG2 and K562 MPRA experiments (28) across DHSs, enhancer regions, and PASs/DASs defined in the relevant cell line (Methods). We observed a striking increase in the enrichment of regulatory SNPs overlapping PASs or DASs, especially those in the top 2% of PASs/DASs ranked by their average delta scores (Fig. 2D), suggesting that PASs and DASs capture meaningful regulatory information.

In total we identified 412,408 PASs and 348,439 DASs across islets, HepG2, and K562 (Fig. S5), 93.7% of which occur in only one biospecimen. The identified PASs were on average 14.7bp long with an average delta of 4. The DASs were 12.4bp long with an average delta of −3. Focusing on islets, within the 9,918 islet enhancers, we identified 74,073 PASs with an average length of 19.5bp (average delta of 2.8) and 67,142 DASs with an average length of 12.2bp (average delta of −2.3), which we used to aid in the interpretation of candidate causal SNPs of signals associated with T2D and glycemic traits.

### The impact of SNPs on islet enhancers

Having determined that TREDNet captures meaningful regulatory patterns across several biospecimens, we applied TREDNet to predict the effect of 67,226,155 SNPs from the genome aggregation database (29) on islet enhancers by calculating an islet enhancer perturbation score (IEP score; Methods) to guide the identification of candidate causal SNPs at T2D-associated genetic signals. This score weights each SNP based on the probability that the surrounding genomic region is an islet enhancer and the predicted effect of each allele on the islet enhancer probability.

To validate the IEP scores, we collected SNPs known to affect various islet features including gene expression (expression quantitative trait loci; eQTLs), exon expression (exonQTLs), chromatin accessibility (caQTLs), and MPRA signals from MIN6 beta cells (Methods). We note that of these validation data, all of the MPRA signals are at SNP level resolution while for the other datasets from population genetic studies, the truly causal SNP(s) is not known in many cases due to linkage disequilibrium (LD). Next, using progressively strict IEP score percentile cutoffs to select groups of SNPs, we calculated the enrichment of SNPs among the islet features, controlling for the distance of a SNP to the nearest gene and the number of SNPs in LD (Methods). We found a strong and progressive enrichment in SNPs shown to activate transcription in MIN6 MPRA experiments and islet chromatin accessibility (caQTLs; Fig. 3A), but not in eQTL or exonQTL signals. These results suggest the IEP score captures meaningful biological effects of SNPs on enhancers and are consistent with studies that report gene expression genetic associations are more strongly enriched in promoter regions than in distal enhancer regions (21, 30).

**Fig. 3.**
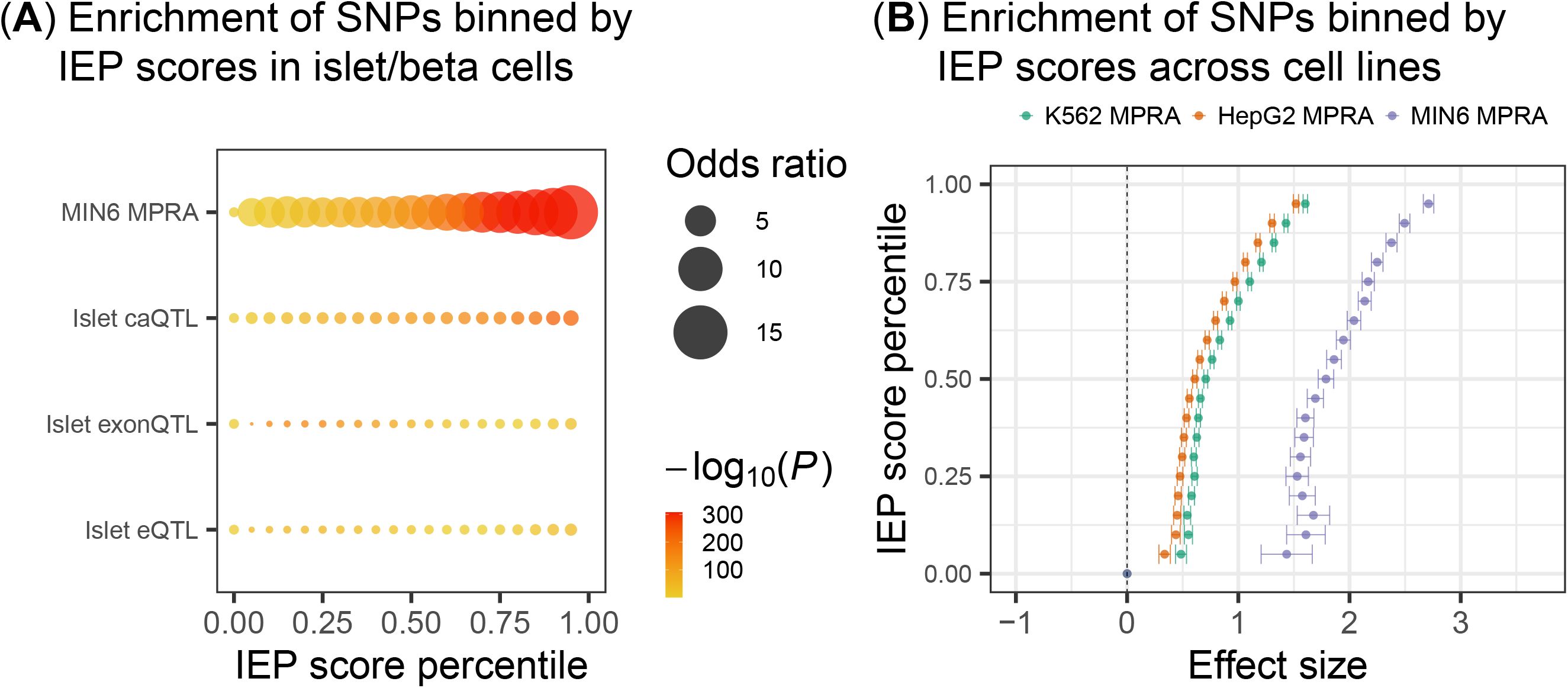
Validation of islet enhancer perturbation (IEP) SNP scores. (A) Enrichment (point size and color) of SNPs grouped by IEP percentile (x-axis) in islet/beta cell validation data (y-axis). (B) Enrichment (log odds ratio) and standard errors (x-axis) of SNPs grouped by IEP percentile (y-axis) in MPRA signals from MIN6 beta cells, K562, and HepG2 (color).

We sought to evaluate the islet specificity of IEP scores and compared the enrichment of SNPs in islet MPRA data to K562 and HepG2 MPRA data (Methods; Fig. 3B). We observed a substantial and progressive enrichment of SNPs in all MPRA data and a stronger enrichment in islet MPRA signals than in K562 or HepG2 (*P*<0.05, z-test). Together, these results suggest that IEP scores capture a broad range of effects, from islet-specific regulatory programs to regulatory programs common across these biospecimens.

### Candidate causal SNPs at genetic loci associated with T2D and glycemic traits

We used TREDNet to refine 99% credible sets of SNPs for 1,243 non-coding genetic signals associated with T2D (31) and glycemic traits including blood glucose levels (32), blood glucose levels after fasting (33), and glycated hemoglobin (HbA1c; (32)). First, we used T2D credible sets from two studies to empirically derive a IEP score cutoff to prioritize candidate causal SNPs: a European ancestry study (6) and a trans-ancestry study (31). We calculated the ratio of the highest IEP score and the second highest IEP score (IEP ratio_1:2_) across all SNPs in the 99% credible set. Because trans-ancestry credible sets have greater power to identify candidate causal SNPs due to different LD patterns, we selected association signals with only one SNP in the credible set from the trans-ancestry analysis. We calculated how many times the IEP ratio_1:2_ correctly nominated the trans-ancestry candidate causal SNP among the multiple SNPs in the credible set identified using the less powered European ancestry study at increasingly stringent IEP ratio_1:2_ thresholds (Methods). At an IEP ratio_1:2_ of >24, we found statistical enrichment (*P*<0.05, hypergeometric test) of IEP ratio_1:2_ refined SNPs from European T2D signals in candidate causal SNPs from the trans-ancestry fine-mapping analysis (Fig. S6).

We applied the IEP ratio_1:2_ threshold of >24 to 99% credible sets for 1,243 non-coding signals associated with T2D (trans-ancestry) and glycemic traits with >1 SNP in the credible set (Fig. 4A). For 101 disease/trait signals, we identified a single candidate causal SNP (Fig. 4B-C; Table S2), spanning 94 total SNPs (i.e., some SNPs were associated with more than one phenotype). To further validate these predictions, we compared the allelic imbalance of the 94 SNPs prioritized by our method to all other SNPs in the 99% credible set using islet chromatin accessibility data (Methods). We found the candidate causal SNPs exhibited greater allele-specific accessibility (*P*=0.001, Wilcoxon rank sum test; Fig. S7), as would be expected if these SNPs perturbed the binding of islet regulatory factors. Although no individual SNP exhibited allelic imbalance after multiple hypothesis correction (likely due to the small number of ATAC-seq samples), several of these SNPs have been shown to exhibit allelic activity in reporter assays conducted in MIN6 beta cells: rs7732130 at the 5:76435004 (*ZBED3/PDE8B*; GRCh37 coordinates) T2D signal (34), rs7933438 at the 11:128040810 (*ETS1*) T2D signal (35), and rs4237150 at the 9:4290085 (*GLIS3*) T2D signal (36). We calculated the overlap of the 94 candidate causal SNPs with predicted TFBSs (based on motifs; Table S3; Methods) and found these SNPs were enriched (FDR<5%) in RFX family binding sites, consistent with previous studies that report RFX6 as an important islet transcription factor for T2D genetic risk (21, 37). In addition, 73 of the 94 candidate SNPs (79%) overlapped a PAS/DAS, a 16.8-fold enrichment compared to random SNPs (*P*=1.6×10^−22^, binomial test), providing an additional level of evidence at these signals.

**Fig. 4.**
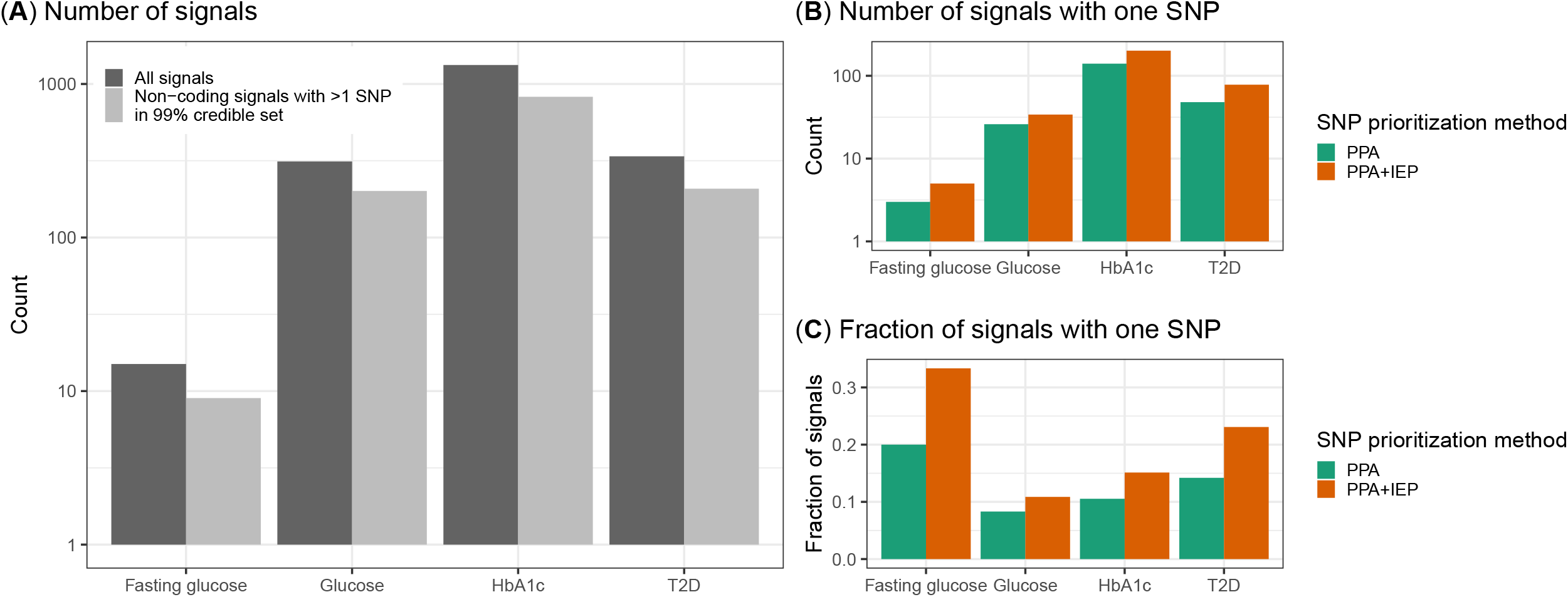
Results of IEP ratio_1:2_ prioritization of 99% credible set SNPs across traits. (A) Total number of independent signals (y-axis) for each disease/trait considered (x-axis). (B) Number of signals with one SNP (y-axis) in the 99% credible set before (green) and after applying the IEP ratio_1:2_ SNP prioritization method (orange) for each disease/trait considered (x-axis). PPA stands for posterior probability of association. (C) Fraction of signals with one SNP (y-axis) in the 99% credible set before (green) and after applying the IEP ratio_1:2_ SNP prioritization method (orange) for each disease/trait considered (x-axis).

To demonstrate the strength of our approach, we (i) highlight three of these signals with extensive evidence from previous studies supporting the predicted candidate causal SNP and (ii) describe the results of an electrophoresis mobility shift assay (EMSA) that we performed for all 99% credible set SNPs at one glucose-associated genetic signal near *PSMA1*.

The 9:4290085 locus near *GLIS3* is associated with T2D, HbA1c, and glucose. Within the 99% T2D credible set, there are two SNPs: rs4237150 and rs1574285 (Fig. S8). Our model identifies the rs4237150 SNP, with the largest T2D posterior probability of association (PPA=0.96), as being the likely causal SNP, where the C allele increases the enhancer probability of the region. rs4237150 overlaps an islet DAS, disrupts NR3C1 and ZNF528 binding motifs (Table S3), and lies in an islet stretch/super enhancer (36). The T2D risk allele (C) exhibits increased allele-specific imbalance in islet ChIP-seq and ATAC-seq data (36). Consistent with the TRENDNet predictions, the C allele of rs4237150 exhibits increased luciferase reporter activity in MIN6 beta cells (36).

Near *DLK1*, there are genetic associations with T2D, HbA1c, and glucose. Within the HbA1c 99% credible set at 14:91785258, there are two SNPs, rs73347525 and rs8004581 (Fig. S9). Our model predicts rs73347525 to be the candidate causal SNP for the HbA1c signal, with the A allele increasing the enhancer probability of the region. rs73347525 is the only SNP in the T2D 99% credible set at this signal, while for glucose it is one of 32 SNPs, among which our model could not make a confident prediction (there are several SNPs in the region with high IEP scores). The T2D risk allele (A) is associated with increased glucose and HbA1c levels, overlaps a beta cell specific ATAC-seq peak (38), and is associated with increased expression of *DLK1* in islets (30). *DLK1* expression patterns are highly specific for islets (30), particularly beta cells (39, 40), and *DLK1* exhibits increased expression in T2D beta cells compared to non-T2D (39). We analyzed the PAS/DAS predictions and found rs73347525 overlaps an islet specific PAS region and most strongly perturbs a motif for ZNF415 (Table S3), where the risk allele (A) results in decreased binding—suggesting that rs73347525 may perturb ZNF415 binding, resulting in increased *DLK1* expression and increased T2D risk.

In addition, near *ZBED3/PDE8B*, there is a strong association with T2D, HbA1c, and glucose. For all three associations, there are three, identical candidate causal SNPs in the 99% credible set (Fig. S10). Our model predicts rs7732130 to be the causal SNP, where the G allele is predicted to increase the probability that the region is an enhancer compared to the alternative allele (A). The G allele is associated with increased T2D risk, increased HbA1c, and increased glucose levels. This SNP intersects an islet PAS and the T2D risk allele (G) is predicted to increase ZNF143 and RFX7 binding affinity (Table S3). Consistent with these findings, the T2D risk allele (G) has been shown to increase both *in vivo* chromatin accessibility in human islets and luciferase reporter activity in MIN6 beta cells (34). Moreover, in human islets, the risk allele (G) is strongly associated with both increased expression of *PDE8B*—a gene with islet-specific expression patterns (21)—and *ZBED3* (30). CRISPR activation and inhibition experiments of the rs7732130 enhancer in human EndoC-βH3 cells, a pancreatic beta cell line, also show effects of this enhancer region on *PDE8B, ZBED3*, and other transcripts within the region (41). Thus, while the effector gene(s) at this locus is unclear, these data cumulatively support rs7732130 as the most likely causal SNP at this locus.

The 11:7117503 locus near *PSMA1* is associated with HbA1c and glucose. For both traits, among all of the 99% credible sets SNPs, our method prioritized rs75336838 as the likely functional variant (Fig. 5A), where the T allele increases the enhancer probability of the region and overlaps an islet DAS region matching several TF binding motifs (Table S3). To test this prediction biochemically, we assessed the effects of the three SNPs in the HbA1c 99% credible set on human beta cell nuclear/transcription factor binding using an EMSA with EndoC-βH3 nuclear extracts. Among the SNPs tested, the candidate causal SNP prioritized by our model, rs75336838, showed the most striking allelic differences in the binding of transcription factors/complexes contained in human EndoC-βH3 nuclear extracts, where the T allele (associated with increased glucose levels) exhibited increased binding (Fig. 5B, arrows). While additional work is needed to complete our molecular understanding at this signal, the nomination of a biochemically-validated likely causal variant should aid future studies in dissecting the mechanism of action.

**Fig. 5.**
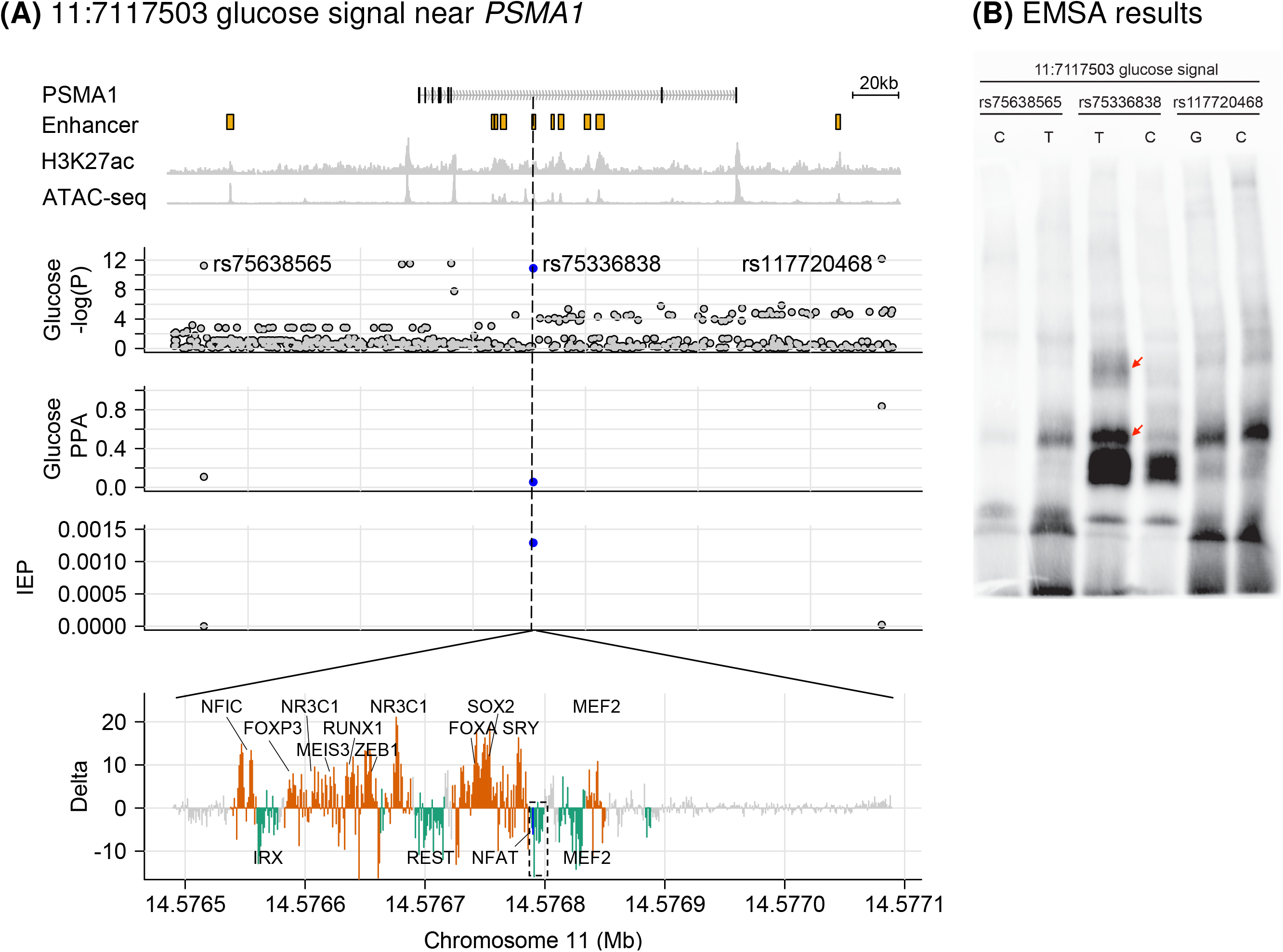
*PSMA1* locus. (A) Locus zoom around the 11:7117503 glucose association (Glucose −*log*_10_(*P*) facet) near *PSMA1*. Top facet shows islet enhancers, called from islet H3K27ac ChIP-seq and ATAC-seq data. rs75336838 (blue) is one of three SNPs in the 99% glucose credible set (PPA facet), has a large IEP score (IEP facet), and occurs in a DAS region (green; PAS regions shown in orange), defined by delta scores from *in silico* saturated mutagenesis (Delta facet). Dashed box indicates the PAS containing the candidate SNP (blue line). (B) Electrophoretic mobility shift assay (EMSA) for all SNPs in the 99% credible set. Red arrows indicate bands of interest.

## Discussion

DL methods have been applied to the problems of genomics extensively. One of the first DL genomics studies implemented a framework, DeepSEA (11), to learn multiple tissue-specific epigenomic signals (e.g., HMs, TFBSs, DHSs) using CNNs in a multi-task learning fashion. The authors later extended the DeepSEA model through the ExPecto (10) model which doubled the number of epigenomic features and increased the depth and breadth of the CNN. Another method, Basset (12), was trained in similar fashion but on DHS regions only. All of these models used an additional layer to predict molecular effects of mutations in DNA regions; however, these effects were calculated across all learned epigenomic features simultaneously and not tailored to detect altered tissue-specific enhancer activity.

Indeed, to date, few studies have applied DL methods to predict enhancers and prioritize candidate causal variants. For instance, Tan et al. trained an ensemble of recurrent neural networks to predict enhancers using features from physicochemical properties of dinucleotides in enhancer regions (14). Yan et al. developed another method, BiRen, that predicts epigenomic signals from DNA sequences using DeepSEA, and combines these predictions with conservation scores of the input sequence to predict enhancers (13). However, neither of these studies performed extensive *post hoc* analyses such as *in silico* mutation analysis or candidate causal variant prioritization. To date, such *post hoc* analyses have been performed primarily using methods that predict epigenomic features directly (10–12, 42) or non-DL enhancer prediction methods, like SVM (15, 43).

In this study, we developed TREDNet, a model that combines the prediction of epigenomic signals and tissue-specific enhancers through a two phase process. Compared to other enhancer prediction methods (13–15), TREDNet consistently improves enhancer detection (Fig. 2A). We found that TREDNet’s enhanced enhancer modeling translates directly to more accurate modeling of signals from *in vitro* MPRA experiments of enhancer regions, compared to previous methods (Fig. 2B). We applied TREDNet to (i) better understand the overall epigenomic regulatory structure of islet enhancers and (ii) refine credible sets from genetic association studies.

By computing the effects of all possible mutations in islet enhancers, we found that we could accurately predict TFBSs from ChIP-seq experiments and TF footprints derived from ATAC-seq data (Fig. S4) as short genomic regions (average length 13.7bp) that greatly alter the overall enhancer probability of a 2kb genomic region, termed PASs and DASs. Previous DL methods for TFBS prediction, such as BPNet (44), have used saliency maps to detect TF motifs (e.g., DeepLIFT (45), TF-MoDISco (46)) and applied the detected motifs to predict TF binding. However, to predict binding sites of a specific TF, such methods require experimental data of the specific TF in question to train the model. By contrast, TREDNet’s peak/dip detection module enables the detection of putative TFBSs of arbitrary TFs throughout the genome. For example, of the 141,215 PASs/DASs detected in islets, 28,081 (20%) do not overlap a known TFBS. We hypothesize that these sites correspond to unmeasured TFBSs or to TFBSs that become bound and active in response to specific stimuli (e.g.,glucose stimulation, stressors like inflammation) and thus are not detected in studies that identify TF binding under baseline conditions. Future studies will be required to test this hypothesis.

Importantly, we used TREDNet predictions to nominate candidate causal mutations at several signals associated with T2D and glycemic traits. For 101 signals (Table S2), we were able to nominate a single candidate causal variant in the 99% credible set from statistical fine-mapping (94 unique SNPs). These predictions include multiple SNPs for which functional allelic effects have been detected previously *in vitro* in islet beta cells (34–36) and one for which we provide biochemical validation. We anticipate that further validation experiments of candidate causal variants nominated in this study will lead to additional insights into the molecular genetic basis of T2D and T2D-related traits.

## Materials and Methods

### Genome annotations

#### ENCODE and NIH Roadmap genomic and epigenomic profiles

We used previously published DNase I hypersensitive sites (DHSs), ChIP-seq peaks of histone marks (HM), and ChIP-seq peaks of transcription factor (TF) binding from the ENCODE (8) and NIH Roadmap (9) studies (1,924 features in total).

#### Islet genomic and epigenomic profiles

We used previously published ATAC-seq data from 33 islets, consisting of 64,129 peaks (30).

We reprocessed previously published H3K27ac ChIP-seq data from two islets (47). Briefly, in order to avoid bias from one specific sample, we downsampled the number of reads from each sample to 26,369,910 reads per sample and merged reads from both samples. We performed a similar process for the input controls for each sample, downsampling to 23,702,108 reads per sample before merging reads across samples. Next, we aligned reads and identified peaks using the ENCODE ChIP-seq processing pipeline (https://github.com/ENCODE-DCC/chip-seq-pipeline2, v1.2.0) with default parameters. We excluded the peaks overlapping Duke blacklisted regions (UCSC browser tables wgEncodeDacMapabilityConsensusExcludable and wgEncodeDacMapabilityConsensusExcludable), resulting in 87,007 H3K27ac peaks.

### Enhancer definitions

We identified 9,918 islet enhancers by selecting ATAC-seq peaks that overlapped an islet H3K27ac ChIP-seq peak (≥1bp) and expanding 1kb up and down the genome from the middle position of the ATAC-seq peak. We considered the entire 2kb region, centered on the ATAC-seq peak, as an enhancer. We removed promoter regions, defined as 1.5kb upstream and 0.5kb downstream (2kb in total) of the transcription start site of known genes from the UCSC genome browser (ftp://hgdownload.soe.ucsc.edu/goldenPath/hg19/database/knownGene.txt.gz).

To define enhancers in HepG2 and K562, we performed the same procedure, except using DHSs rather than ATAC-seq peaks (278,000 DHSs for HepG2 and 342,000 DHSs for K562). We identified 39,892 HepG2 and 42,697 K562 enhancers in total.

### Two phase deep learning model to predict enhancers

We developed a two phase deep learning (DL) classifier, TREDNet, based on convolutional neural networks (CNNs; implemented in keras v2.1.2 and tensorflow-GPU v1.4.1) to predict enhancers from DNA sequence. In the first phase, we used a model with six convolutional layers (∼143 million trainable parameters; Table S4) to predict 1,924 different genomic and epigenomic features simultaneously for a 2kb genomic region. These features included DHSs, TF ChIP-seq peaks, and histone mark ChIP-seq peaks from the ENCODE (8) and NIH Roadmap (9) studies. We tiled the entire human genome using a sliding window of length 2kb and the step length of 200bp. We selected those segments that overlapped at least one of the 1,924 epigenomic features and trained the DL model using these segments. For training, we fit the model on all autosomes, except for chromosomes 8 and 9. During training, we used signals on chromosome 7 as a validation set. We used signals on chromosomes 8 and 9 for testing the final model, evaluating both the area under the receiver operating characteristic curve (auROC) and area under the precision recall curve (auPRC; Fig. S1). Due to the cell/tissue type-specific nature of the features, the class distribution (positive/negative) in the training set was imbalanced across the features, with only 2% of the dataset being positive cases on average. We compared the TREDNet phase one model to previously published, similar epigenomic feature prediction models—ExPecto (10), DeepSEA (11), and Basset (12)—on the data used for TREDNet with the same testing strategy (i.e., testing performance on chromosomes 8 and 9).

For the second phase, we fit three smaller models to predict pancreatic islet, HepG2, and K562 enhancers (one for each cell/tissue type) from the output of the first model (a vector of the 1,924 epigenomic predictions for 2kb sequence segments). The second model consisted of two convolutional layers resulting in ∼12 million trainable parameters (Table S5). For training, we adopted a similar strategy as phase one, using chromosome 7 to validate during training and withholding chromosomes 8 and 9 to evaluate the final model. We tested the model’s performance (auROC and auPRC) using enhancers from chromosomes 8 and 9 withheld from training (Fig. 2A). To benchmark the TREDNet phase two model, we tested previously described enhancer prediction models—BiRen (13), Tan et al. (14), and SVM (15)—on the data used for TREDNet with the same testing strategy (i.e., testing performance on chromosomes 8 and 9). For methods that did not distribute trained models, SVM and BiRen, we trained models using the same data and training strategy as for TREDNet. For Tan et al., the method produces a binary output for each input region based on the scores generated by five different models. To generate auROC and auPRC values, we used the scores generated by each of the five Tan et al. models and picked the best-performing one.

### Analysis of *in vitro* enhancer mutagenesis experiments

We validated TRENDNet enhancer probability scores using data from massively parallel reporter assays (MPRAs) that tested the effect of mutations on reporter expression in enhancer regions near *ZFAND3* and *TCF7L2* in MIN6 and *SORT1* in HepG2 (16). Using the log2 allelic MPRA expression effect estimates distributed by the authors, we filtered for alleles with >=50 unique barcode tags (which maximized the correlation between replicates as reported in Kircher et al. (16)) and for alleles that showed a statistical difference in the MPRA expression read out (FDR<5%, Benjamini-Hochberg procedure (48)). Using the TREDNet enhancer model corresponding to each MPRA experiment (i.e., the islet model for MIN6 and the HepG2 model for HepG2), we calculated the log2 fold change of enhancer probability scores for the alternate allele compared to the reference allele. We performed the same procedure to generate computational allelic scores using the other enhancer modeling methods considered in this study: Tan et al. and SVM. We did not include BiRen because as input the method takes coordinates of genomic regions, not DNA sequences, and therefore is not applicable to model mutagenesis experiments. We also aggregated computational allelic scores reported previously in Kircher et al. (16): DeepSEA, CADD (17), Eigen (18), and FATHMM (19). For each computational allelic score, we calculated the Spearman’s rank-order correlation with the MPRA derived allelic score.

### Calculation of delta scores from *in silico* saturated mutagenesis

For each biospecimen, we performed *in silico* saturated mutagenesis of enhancer regions to evaluate the effects of mutations on the overall enhancer probability score. For each 2kb enhancer region (see “Enhancer definitions”), we calculated a delta score iteratively for each nucleotide position against the GRCh37 reference sequence (i.e., we mutate each nucleotide to all possible mutations but keep the remaining 1,999 nucleotide sequence the same as the reference):

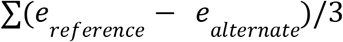

where the *e* term represents the probability that the 2kb sequence is an enhancer, *reference* indicates the GRCh37 reference nucleotide, and *alternate* indicates a non-reference nucleotide. This delta score, generated for each bp in the 2kb enhancer region, predicts the effects of mutations at a specific base on the overall enhancer probability of the region, such that a positive score indicates a negative change in the enhancer probability (enhancer-damaging) and a negative score indicates a positive change in the enhancer probability (enhancer-strengthening).

### Identification and analysis of transcription factor binding sites (TFBSs)

For HepG2 and K562, we used TF ChIP-seq data spanning 77 TFs in HepG2 and 150 TFs in K562 (8). For each TF, in order to extract the TFBS position, we used HOMER v4.11 (49) to scan motifs found in the database packaged with HOMER (http://homer.ucsd.edu/homer/motif/motifDatabase.html), producing a list of predicted TFBSs for each motif in the database. We ranked the motifs in decreasing order of their enrichment *P*-value and kept the TF of the most enriched motif.

Because islets do not have as comprehensive TF ChIP-seq profiles as HepG2 and K562, we used predicted TFBSs derived from ATAC-seq footprints described previously (21). Briefly, for two islet ATAC-seq samples, Varshney et al. scanned for potential transcription factor binding sites (TFBSs) in a haplotype-aware manner using the “find individual motif occurrences” (FIMO) tool (50) with position weight matrices (PWMs) from a previously described database (51) consisting of PWMs from ENCODE (52), JASPAR (53), and Jolma et al. (54). Next, Varshney et al. used CENTIPEDE (55) to call footprints in the islets ATAC-seq data, considering a given motif occurrence bound if both the CENTIPEDE posterior probability was ≥0.99 and the motif’s coordinates were fully contained within an ATAC-seq peak.

For islets, because many of the predicted TFs share similar motif patterns and result in overlapping predicted TFBSs, we reduced redundancy by aggregating islet TF footprints with similar binding motifs. For each TF, we calculated the average delta score across all nucleotides within the predicted binding sites. For the TFs with a positive average delta score across all predicted TFBSs (690 TFs), we selected 156 TFs where the average lower boundary of 95% confidence interval (CI) the binding region was greater than the upper boundary of the CI of the flanking region (defined as 10bp on each side of the TFBS). For the TFs with a negative average delta score across all predicted TFBSs (49 TFs), we selected 10 TFs where the delta scores within the CI were all negative. Using these 166 TFs, we iteratively merged TFs if >40% of their predicted binding sites overlapped and the overlapping regions were greater than half of either binding site. In total, this procedure resulted in 100 non-redundant TFs groups for islets, which we used to analyze delta scores in islet enhancers.

We used TFBSs in islets, K562, and HepG2 to evaluate if delta scores mark TFBSs by comparing the absolute value of delta scores within TFBSs to 20bp regions immediately flanking each TFBS as well as randomly sampled enhancer regions. To generate random delta scores, we shuffled the TFBSs within enhancer regions for each biospecimen 10 times and recorded the absolute value of delta scores within these regions.

In addition, to compare delta scores to evolutionary conservation profiles at TFBSs, we calculated the information content (IC) at each position of PWMs for each TF. For evolutionary conservation profiles, we used phyloP scores generated from 46 vertebrate species (56, 57). Across all TF PWMs for each biospecimen, we calculated the correlation (Spearman’s rho) between the IC of each PWM position and delta/phyloP scores.

### Detection of peak and dip active sites

For each biospecimen, we trained two DL models, one to predict peak active sites (PASs) and one to predict dip active sites (DASs), within enhancers from delta score enhancer profiles (six models total across all three cell lines).

To train the PAS classifier for each biospecimen, we annotated nucleotides within enhancers with 1 (positive set) if the nucleotide overlapped a TFBS and otherwise 0 (control set). We subsequently excluded from the control set (i.e., enhancer nucleotides annotated as 0) genomic regions (i) between any two TFBSs in an enhancer (even if these TFBSs are on the opposite ends of the enhancer), (ii) within 10bp of a TFBS, (iii) within 20bp of an enhancer boundary, and (iv) in an enhancer of less than 50bp. For HepG2 and K562, we used all TFBSs, described in “Identification and analysis of transcription factor binding sites”. For islets, we used the TFBSs from the 166 TFBSs derived from ATAC-seq footprints, described in “Identification and analysis of transcription factor binding sites”.

For each enhancer nucleotide, we derived a series of features from delta score predictions across various window sizes and used these features to predict the location of TFBSs within the enhancer sequence (i.e., the positive or control status of each nucleotide within the enhancer). For each nucleotide, we scanned a series of windows from 10bp in length to 1bp in length. For each window length >7bp, we defined a core region as the 6bp in the center of the window and calculated the following metrics: (i) the average delta score of nucleotides within the window, (ii) the maximum delta score of nucleotides within the window, (iii) the fraction of nucleotides within the window with a positive delta score, and (iv) the fraction of nucleotides within the core region with a positive delta score. For windows of length <6bp, we repeated the same procedure, but using a core region equal to the window size. For each window length, we iteratively scanned around the target nucleotide such that the target nucleotide occupied every position within the window (i.e., we incremented the window position by one base pair for each iteration resulting in 10 sliding windows for a window size of 10bp).

For a single nucleotide, the result of this procedure was a series of four metrics defined across 55 windows (of size 10bp to 1bp). We concatenated these metrics together across all windows to generate a vector of 220 values for each nucleotide, representing a comprehensive description of a local mutational impact of each nucleotide on an enhancer. After iteratively performing this procedure across all nucleotides within enhancers, we then fit a two layer CNN (implemented in keras v2.0.8; architecture described in Table S6) to predict the TFBS annotation status (0 or 1) from the 220 values calculated for each nucleotide. During training, we randomly sampled 20% of the input data as a validation set and excluded chromosomes 8 and 9 entirely. We used signals on chromosomes 8 and 9 for testing the final model, evaluating both the auROC and auPRC (Fig. S4).

To train the DAS classifier, we performed the same procedure, except we selected for TFBSs where the average delta score was negative.

Both the PAS and DAS models were highly accurate (Fig. S4) and resulted in a score for each nucleotide within an enhancer representing the likelihood that the nucleotide overlaps a TFBS. We used these scores to call PAS and DAS regions. For PAS regions, we used the peak model for each biospecimen and labeled any span of nucleotides (one after the other) of length >=3 with a likelihood score >=0.312 as a PAS. We repeated the same procedure with the dip models, calling regions with >=3 consecutive predicted dip nucleotides and a likelihood score >=0.152 as DASs.

To validate the PAS and DAS predictions as regulatory sites within enhancer sequences, we compared the density of SNPs reported to have an allelic effect on transcription in K562 and HepG2 MPRA experiments (28) in DHSs, enhancer regions, and PAS/DAS regions ranked by their average delta score. For comparisons, we calculated the number of MPRA validated SNPs in each genomic region divided by the total length of the region.

### Calculation and validation of islet enhancer perturbation scores

We generated a catalog of predicted effects of 67,226,155 SNPs on islet enhancers using the genome Aggregation Database (gnomAD) v3.0 (29), including SNPs with a minor allele frequency (MAF) >=0.0001. Since gnomAD v3.0 uses GRCh38 coordinates, we lifted these coordinates over to GRCh37 to match the data used to train the model. Across all SNPs, we calculated islet enhancer perturbation (IEP) scores using a two phase procedure. First for each SNP, we calculated the probability, *e*, of either allele falling in an enhancer, given 2kb of the flanking reference sequence (GRCh37) surrounding the SNP. Next, we generated IEP scores:

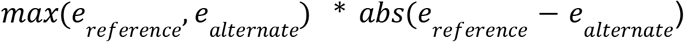

where the *e* term represents the probability of an allele residing in an islet enhancer. For enrichment calculations and subsequent binning, we calculated the percentile rank of IEP scores.

To validate IEP scores, we used previously published islet genetic studies spanning gene/exon expression (30), chromatin accessibility (58), and MPRA data generated from the MIN6 mouse pancreatic islet beta cell line (35). From these data, we generated genome annotations that were subsequently used for enrichment calculations. For the genetic association data (eQTLs, exonQTLs, and caQTLs), we selected all QTLs and marked the genomic location of all SNPs in LD (r^2^>0.8) with the lead SNP (minimum *P*-value at the locus). For the MIN6 MPRA data, we marked the genomic location of SNPs reported to induce activity in either the unstimulated (baseline) or stimulated (endoplasmic reticulum stress) conditions. We did not LD expand the MPRA SNPs because the MPRA data has SNP resolution since the alleles of single SNP were tested in a reporter construct, thereby breaking the LD structure that exists naturally in the human genome. Using these annotations, we calculated the enrichment of SNPs across progressive percentile cutoffs (from 0.05 to 1 in steps of 0.05) using GARFIELD v2.0 (59), a logistic regression method that controls for the distance of each SNP to the nearest gene and the number of SNPs in LD. For these enrichments, we used the 24 million SNPs included in the GARFIELD package.

For islet specificity comparisons, we calculated the enrichment of previously published MPRA data from the K562 and HepG2 cell lines (28). We compared the enrichment coefficients of the MIN6 MPRA data to K562 and HepG2 using a z-test as described in Paternoster et al. (60).

### Refinement of credible sets for T2D and glycemic traits

To empirically define a threshold for prioritizing candidate causal SNPs using IEP scores, we used 99% credible sets from two T2D fine-mapping genetic studies (uniform priors): a European ancestry study (6) and a trans-ancestry study (31). To focus on likely distal regulatory signals, we excluded signals where >=1 SNP fell in a coding region from both datasets. For each T2D-associated signal in each study, we calculated the ratio of the IEP score of the largest IEP score to the second largest IEP score (IEP ratio_1:2_). Next, we treated the trans-ancestry study as a “truth set”, since trans-ancestry studies have greater power to fine-map due to different LD patterns across ancestries, and asked how often we could nominate the trans-ancestry candidate causal SNP using progressive IEP ratio_1:2_ cutoffs in the European ancestry study. To make the trans-ancestry “truth set”, we selected the 11 signals with one SNP in the 99% credible set that were not fine-mapped to a single candidate causal SNP in the European ancestry study. We selected the 179 from the European ancestry study with >1 SNP in the 99% credible set and performed a hypergeometric test (phyper function in R) across progressive IEP ratio_1:2_ thresholds. We selected the minimum IEP ratio_1:2_ cutoff with *P*<0.05, corresponding to an IEP ratio_1:2_ of 24, and applied the cutoff to the trans-ancestry credible sets to identify additional signals with a signal candidate causal SNP.

We applied the IEP ratio_1:2_ of 24, to 99% credible sets for T2D (31) and glycemic traits including blood glucose levels (32), blood glucose levels after fasting (33), and glycated hemoglobin (HbA1c; (32)).

### Allelic imbalance analysis

For allelic imbalance analysis, we collected 24 islet ATAC-seq samples for which SNP genotypes were also available: 1 sample from Varshney et al. (21), 10 samples from Rai et al. (38), and 13 samples from Vinuela et al. (30). We tested for allelic imbalance at all SNPs in the 99% credible sets for T2D and glycemic traits where our model predicts a single candidate causal SNP that previously had >1 candidate causal SNP at each association signal (Table S2). We followed the computational procedure outlined in Greenwald et al. (34). Briefly, we mapped reads using bwa v0.7.17-r1194-dirty (61) and filtered reads with low mapping quality (“-q 30 −M”). We then used WASP v0.3.4 (62) to remove duplicate reads and correct for reference mapping bias. Using a binomial test to assess imbalance on a per sample basis, we tested for allelic imbalance at SNPs that had at least two heterozygotes with two or more reads covering each allele. Finally, we calculated z-scores and used Stouffer’s method to calculate a combined z-score and *P*-value across samples, weighting the scores by the sequencing depth of each sample. We controlled for the number of tests using the Benjamini-Hochberg procedure.

### TFBSs enrichment of candidate causal SNPs

To calculate the enrichment of the candidate causal SNPs in TFBSs, we calculated the fraction of SNPs that overlap TFBSs predicted from ATAC-seq footprints (see “Identification and analysis of transcription factor binding sites”) using both the candidate causal set and a 10-fold control set, derived from islet DHS regions. We computed the fold-enrichment, calculated *P*-values using Fisher’s exact test, and controlled for the number of tests using the Benjamini-Hochberg procedure.

### Calculation of the effects of candidate causal SNPs on TFBS motifs

To calculate the allelic effects of candidate causal SNPs on binding motifs, we used TFBS motifs from ENCODE (52), JASPAR (53), and HOMER (http://homer.ucsd.edu/homer/motif/motifDatabase.html). We ran FIMO (packaged with MEME v4.9.0) with a *P*-value threshold of 0.01, keeping motifs that had hits for the sequences of both alleles. The remaining motifs were sorted in the decreasing order of the value |log(*P*-value_ref_) – log(*P*-value_alt_|).

### Electrophoresis mobility shift assay (EMSA) experiments

For EMSA experiments, we designed 21bp biotin end-labeled complementary oligonucleotides with each SNP allele tested centered at the 11th position of the oligo (Integrated DNA Technologies; Table S7). Each forward and reverse oligo for the biotinylated probes were biotinylated at their 5’ ends. We annealed complementary oligos to create double-stranded probes for each tested sequence. Using the NE-PER Extraction Kit (Thermo Scientific), we prepared nuclear extract from human EndoC-βH3 cells in the proliferating, non-excised state (63) and completed EMSAs using the LightShift Chemiluminescent EMSA kit (Thermo Scientific) according to the manufacturer’s instructions. Each binding reaction contained 1X binding buffer, 1 μg poly dI-dC, 4 μg EndoC-βH3 nuclear extract, and 200 fmol of biotinylated double-stranded probe. We incubated reactions at 25°C for 25 minutes, after which we resolved DNA-protein complexes on a 6% DNA retardation gel (Invitrogen) and detected them by chemiluminescence after transfer and UV crosslinking to a nitrocellulose membrane.

## Data Availability

The models from this study (architecture and weights), islet enhancer locations, PAS/DAS locations, and IEP scores for gnomAD SNPs are available through zenodo (https://doi.org/10.5281/zenodo.6463875).

## Acknowledgements

We thank Stephen CJ Parker, Arushi Varshney, and Chad Krilow for supporting the work presented in this study. This research was funded in part by United States National Institutes of Health grants 1-ZIA-HG000024 (to F.S.C.), 1-ZIA-LM200881-12 (to I.O.), R01DK118011 (to M.L.S.), and the Department of Defense Peer-Reviewed Medical Research Program grant W81XWH-18-0401 (to M.L.S.).

## References

1. M. Claussnitzer, et al., A brief history of human disease genetics. Nature (2020).

2. S. H. Orkin, D. E. Bauer, Emerging genetic therapy for sickle cell disease. Annu. Rev. Med. 70, 257–271 (2019).

3. M. S. Sabatine, PCSK9 inhibitors: clinical evidence and implementation. Nat. Rev. Cardiol. 16, 155–165 (2019).

4. A. Hutchinson, J. Asimit, C. Wallace, Fine-mapping genetic associations. Hum. Mol. Genet. 29, R81–R88 (2020).

5. S. E. Kahn, M. E. Cooper, S. Del Prato, Pathophysiology and treatment of type 2 diabetes: perspectives on the past, present, and future. Lancet 383, 1068–1083 (2014).

6. A. Mahajan, et al., Fine-mapping type 2 diabetes loci to single-variant resolution using high-density imputation and islet-specific epigenome maps. Nat. Genet. 50, 1505–1513 (2018).

7. I. Cebola, Pancreatic islet transcriptional enhancers and diabetes. Curr. Diab. Rep. 19, 145 (2019).

8. ENCODE Project Consortium, An integrated encyclopedia of DNA elements in the human genome. Nature 489, 57–74 (2012).

9. Roadmap Epigenomics Consortium, et al., Integrative analysis of 111 reference human epigenomes. Nature 518, 317–330 (2015).

10. J. Zhou, et al., Deep learning sequence-based ab initio prediction of variant effects on expression and disease risk. Nat. Genet. 50, 1171–1179 (2018).

11. J. Zhou, O. G. Troyanskaya, Predicting effects of noncoding variants with deep learning-based sequence model. Nat. Methods 12, 931–934 (2015).

12. D. R. Kelley, J. Snoek, J. L. Rinn, Basset: learning the regulatory code of the accessible genome with deep convolutional neural networks. Genome Res. 26, 990–999 (2016).

13. B. Yang, et al., BiRen: predicting enhancers with a deep-learning-based model using the DNA sequence alone. Bioinformatics 33, 1930–1936 (2017).

14. K. K. Tan, N. Q. K. Le, H.-Y. Yeh, M. C. H. Chua, Ensemble of deep recurrent neural networks for identifying enhancers via dinucleotide physicochemical properties. Cells 8 (2019).

15. D. Lee, LS-GKM: a new gkm-SVM for large-scale datasets. Bioinformatics 32, 2196–2198 (2016).

16. M. Kircher, et al., Saturation mutagenesis of twenty disease-associated regulatory elements at single base-pair resolution. Nat. Commun. 10, 3583 (2019).

17. M. Kircher, et al., A general framework for estimating the relative pathogenicity of human genetic variants. Nat. Genet. 46, 310–315 (2014).

18. I. Ionita-Laza, K. McCallum, B. Xu, J. D. Buxbaum, A spectral approach integrating functional genomic annotations for coding and noncoding variants. Nat. Genet. 48, 214–220 (2016).

19. H. A. Shihab, et al., An integrative approach to predicting the functional effects of non-coding and coding sequence variation. Bioinformatics 31, 1536–1543 (2015).

20. P. K. Koo, M. Ploenzke, Deep learning for inferring transcription factor binding sites. Current Opinion in Systems Biology 19, 16–23 (2020).

21. A. Varshney, et al., Genetic regulatory signatures underlying islet gene expression and type 2 diabetes. Proc Natl Acad Sci USA 114, 2301–2306 (2017).

22. Y. Zhou, et al., TCF7L2 is a master regulator of insulin production and processing. Hum. Mol. Genet. 23, 6419–6431 (2014).

23. N. Gao, et al., Foxa1 and Foxa2 maintain the metabolic and secretory features of the mature beta-cell. Mol. Endocrinol. 24, 1594–1604 (2010).

24. D. A. Glauser, W. Schlegel, The emerging role of FOXO transcription factors in pancreatic beta cells. J. Endocrinol. 193, 195–207 (2007).

25. C. A. Zahnow, CCAAT/enhancer-binding protein beta: its role in breast cancer and associations with receptor tyrosine kinases. Expert Rev. Mol. Med. 11, e12 (2009).

26. A. Miura, et al., Hepatocyte nuclear factor-4alpha is essential for glucose-stimulated insulin secretion by pancreatic beta-cells. J. Biol. Chem. 281, 5246–5257 (2006).

27. H. Nakabayashi, et al., Clock-controlled output gene Dbp is a regulator of Arnt/Hif-1β gene expression in pancreatic islet β-cells. Biochem. Biophys. Res. Commun. 434, 370–375 (2013).

28. J. van Arensbergen, et al., High-throughput identification of human SNPs affecting regulatory element activity. Nat. Genet. 51, 1160–1169 (2019).

29. K. J. Karczewski, et al., The mutational constraint spectrum quantified from variation in 141,456 humans. Nature 581, 434–443 (2020).

30. A. Viñuela, et al., Genetic variant effects on gene expression in human pancreatic islets and their implications for T2D. Nat. Commun. 11, 4912 (2020).

31. A. Mahajan, et al., Trans-ancestry genetic study of type 2 diabetes highlights the power of diverse populations for discovery and translation. medRxiv (2020) https://doi.org/10.1101/2020.09.22.20198937.

32. N. Sinnott-Armstrong, et al., Genetics of 35 blood and urine biomarkers in the UK Biobank. Nat. Genet. 53, 185–194 (2021).

33. J. Dupuis, et al., New genetic loci implicated in fasting glucose homeostasis and their impact on type 2 diabetes risk. Nat. Genet. 42, 105–116 (2010).

34. W. W. Greenwald, et al., Pancreatic islet chromatin accessibility and conformation reveals distal enhancer networks of type 2 diabetes risk. Nat. Commun. 10, 2078 (2019).

35. S. Khetan, et al., Functional characterization of T2D-associated SNP effects on baseline and ER stress-responsive β cell transcriptional activation. Nat. Commun. 12, 5242 (2021).

36. A. Aylward, J. Chiou, M.-L. Okino, N. Kadakia, K. J. Gaulton, Shared genetic risk contributes to type 1 and type 2 diabetes etiology. Hum. Mol. Genet. (2018) https://doi.org/10.1093/hmg/ddy314.

37. J. T. Walker, et al., RFX6-mediated dysregulation defines human β cell dysfunction in early type 2 diabetes. BioRxiv (2021) https://doi.org/10.1101/2021.12.16.466282.

38. V. Rai, et al., Single-cell ATAC-Seq in human pancreatic islets and deep learning upscaling of rare cells reveals cell-specific type 2 diabetes regulatory signatures. Mol. Metab. 32, 109–121 (2020).

39. N. Lawlor, et al., Single-cell transcriptomes identify human islet cell signatures and reveal cell-type-specific expression changes in type 2 diabetes. Genome Res. 27, 208–222 (2017).

40. J. Li, et al., Single-cell transcriptomes reveal characteristic features of human pancreatic islet cell types. EMBO Rep. 17, 178–187 (2016).

41. I. Miguel-Escalada, et al., Human pancreatic islet three-dimensional chromatin architecture provides insights into the genetics of type 2 diabetes. Nat. Genet. 51, 1137–1148 (2019).

42. A. Wesolowska-Andersen, et al., Deep learning models predict regulatory variants in pancreatic islets and refine type 2 diabetes association signals. eLife 9 (2020).

43. D. Lee, et al., A method to predict the impact of regulatory variants from DNA sequence. Nat. Genet. 47, 955–961 (2015).

44. Ž. Avsec, et al., Base-resolution models of transcription-factor binding reveal soft motif syntax. Nat. Genet. 53, 354–366 (2021).

45. A. Shrikumar, P. Greenside, A. Kundaje, Learning Important Features Through Propagating Activation Differences. arXiv (2017).

46. A. Shrikumar, et al., Technical Note on Transcription Factor Motif Discovery from Importance Scores (TF-MoDISco) version 0.5.6.5. arXiv (2018).

47. S. C. J. Parker, et al., Chromatin stretch enhancer states drive cell-specific gene regulation and harbor human disease risk variants. Proc Natl Acad Sci USA 110, 17921–17926 (2013).

48. Y. Benjamini, Y. Hochberg, Controlling the false discovery rate: A practical and powerful approach to multiple testing. Journal of the Royal Statistical Society. Series B (Methodological) 57, 289–300 (1995).

49. S. Heinz, et al., Simple combinations of lineage-determining transcription factors prime cis-regulatory elements required for macrophage and B cell identities. Mol. Cell 38, 576–589 (2010).

50. C. E. Grant, T. L. Bailey, W. S. Noble, FIMO: scanning for occurrences of a given motif. Bioinformatics 27, 1017–1018 (2011).

51. L. J. Scott, et al., The genetic regulatory signature of type 2 diabetes in human skeletal muscle. Nat. Commun. 7, 11764 (2016).

52. P. Kheradpour, M. Kellis, Systematic discovery and characterization of regulatory motifs in ENCODE TF binding experiments. Nucleic Acids Res. 42, 2976–2987 (2014).

53. A. Mathelier, et al., JASPAR 2016: a major expansion and update of the open-access database of transcription factor binding profiles. Nucleic Acids Res. 44, D110–5 (2016).

54. A. Jolma, et al., DNA-binding specificities of human transcription factors. Cell 152, 327–339 (2013).

55. R. Pique-Regi, et al., Accurate inference of transcription factor binding from DNA sequence and chromatin accessibility data. Genome Res. 21, 447–455 (2011).

56. G. M. Cooper, et al., Distribution and intensity of constraint in mammalian genomic sequence. Genome Res. 15, 901–913 (2005).

57. A. Siepel, K. S. Pollard, D. Haussler, “New Methods for Detecting Lineage-Specific Selection” in Research in Computational Molecular Biology, Lecture notes in computer science., A. Apostolico, C. Guerra, S. Istrail, P. A. Pevzner, M. Waterman, Eds. (Springer Berlin Heidelberg, 2006), pp. 190–205.

58. S. Khetan, et al., Type 2 Diabetes-Associated Genetic Variants Regulate Chromatin Accessibility in Human Islets. Diabetes 67, 2466–2477 (2018).

59. V. Iotchkova, et al., GARFIELD classifies disease-relevant genomic features through integration of functional annotations with association signals. Nat. Genet. 51, 343–353 (2019).

60. R. Paternoster, R. Brame, P. Mazerolle, A. Piquero, Using the correct statistical test for the equality of regression coefficients. Criminology. 36, 859–866 (1998).

61. H. Li, R. Durbin, Fast and accurate long-read alignment with Burrows-Wheeler transform. Bioinformatics 26, 589–595 (2010).

62. B. van de Geijn, G. McVicker, Y. Gilad, J. K. Pritchard, WASP: allele-specific software for robust molecular quantitative trait locus discovery. Nat. Methods 12, 1061–1063 (2015).

63. M. Benazra, et al., A human beta cell line with drug inducible excision of immortalizing transgenes. Mol. Metab. 4, 916–925 (2015).

