## Supplemental figures and tables for "Modeling islet enhancers using deep learning identifies candidate causal variants at loci associated with T2D and glycemic traits"

### Supplementary Figures and Tables

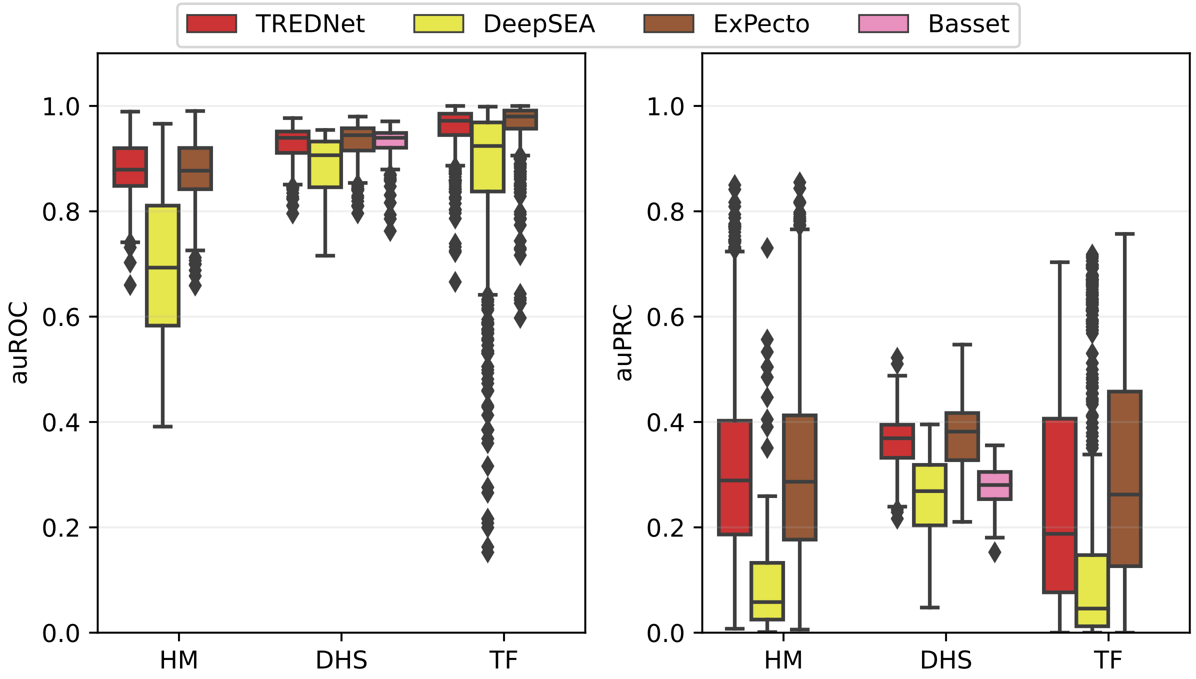


**Fig. S1. Characterization of TREDNet phase one.** Phase one TREDNet peak prediction accuracy for transcription factors (TFs), histone modifications (HMs), and DNase I hypersensitivity sites (DHSs; x-axis) compared to other models (colors) using area under the receiver operating characteristic (auROC; left) and area under the precision recall curve (auPRC; right) metrics (y-axis).

**(A)** Average delta and phyloP scores across FOXA binding sites in islet enhancers

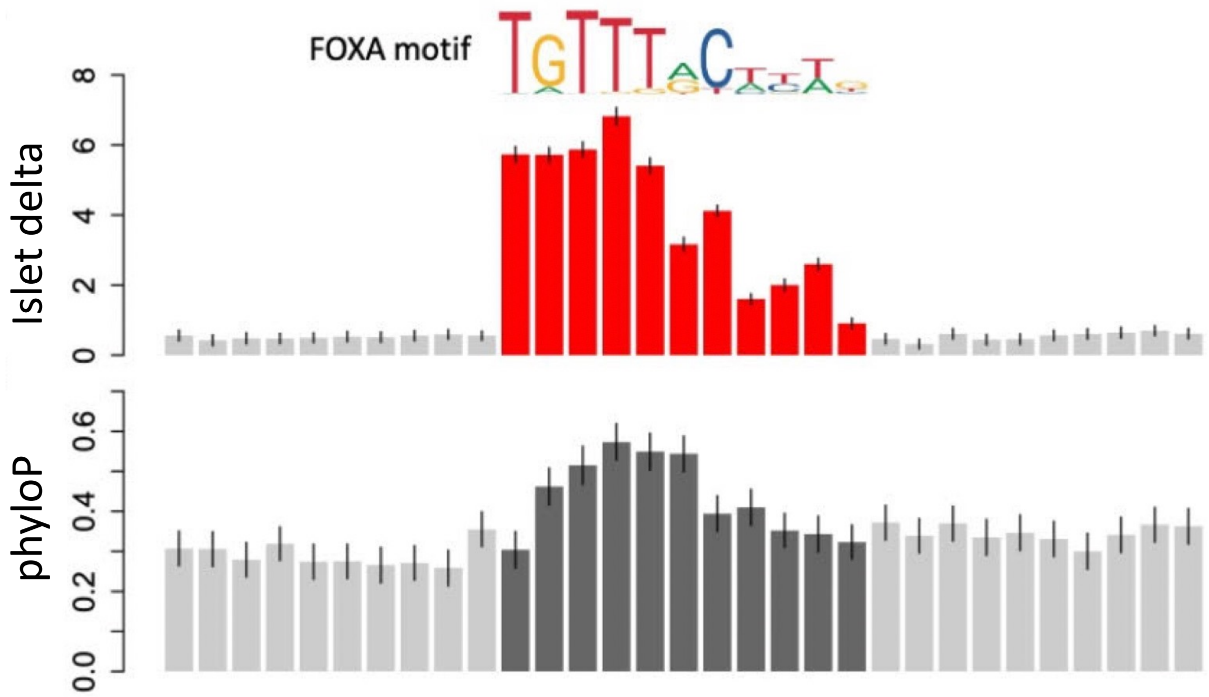


**(B)** Correlation between delta/phyloP scores and information content

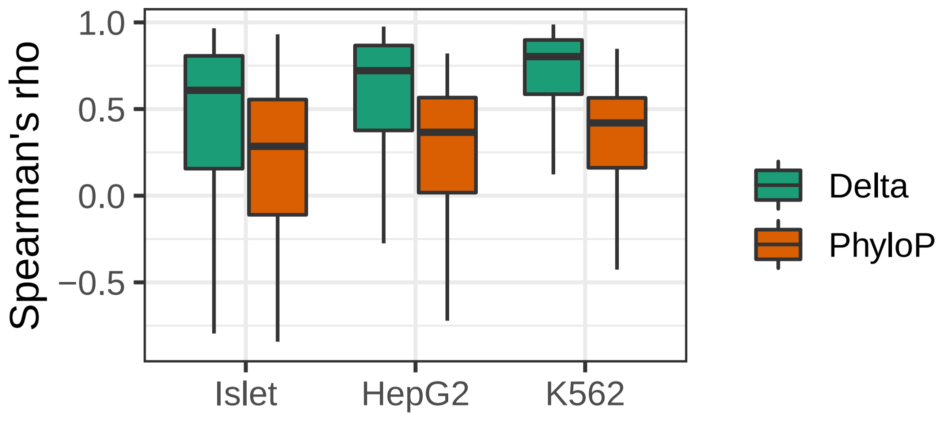


**Fig. S2. Comparison of delta scores and motif information content.** (A) FOXA position weight matrix (PWM; top panel). The average delta score of each position at FOXA transcription factor binding sites (TFBSs), including a 20bp flanking region around the central motif, using the islet TREDNet model (middle panel). The average evolutionary conservation phyloP scores of each position at FOXA TFBSs, including a 20bp flanking region around the central motif (bottom panel). (B) Distribution of correlation coefficients (Spearman's rho; y-axis) between delta/phyloP scores and information content (IC) of each position in PWMs for TFBSs in enhancer regions across biospecimens (x-axis).

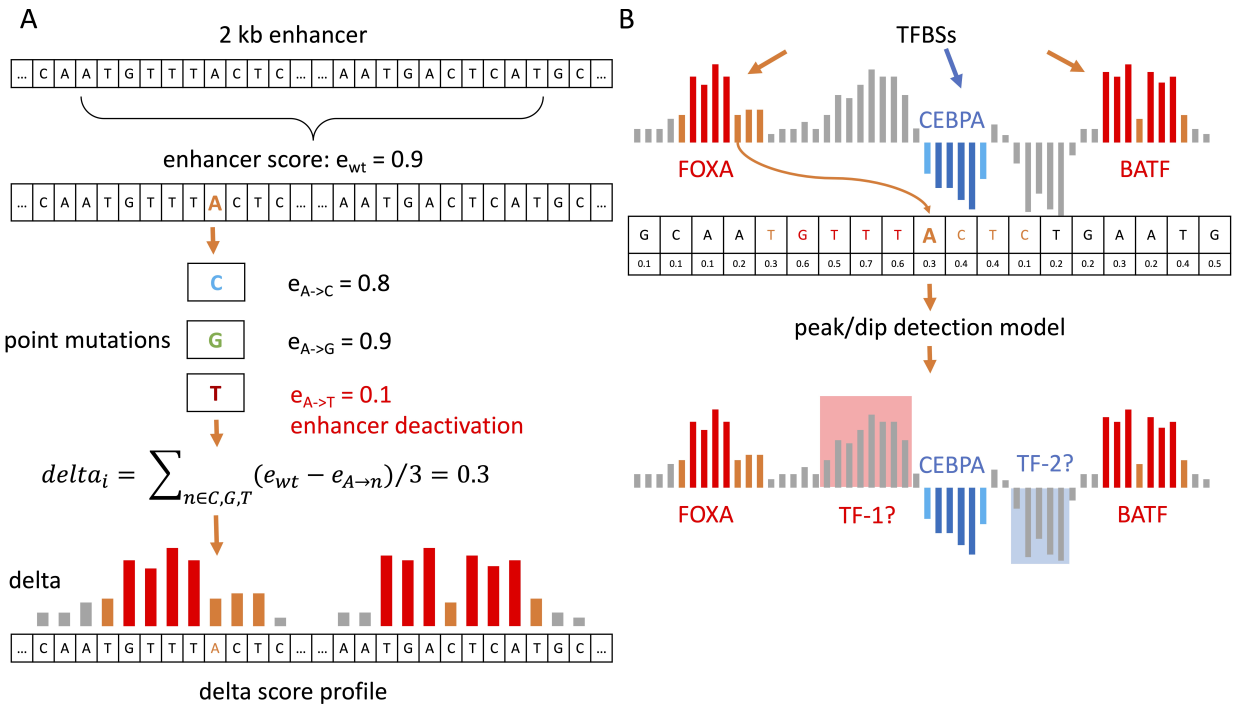


**Fig. S3. Detection framework for peak and dip active sites.** (A) For each 2kb enhancer region, we calculate the TREDNet enhancer probability score ($e$) using the "wildtype" GRCh37 reference sequence ($e_{wt}$). To predict the mutational effect of each nucleotide, we calculate enhancer probability scores for all three non-reference nucleotides at each enhancer position while the remaining enhancer DNA sequence remains unchanged. We generate delta score mutational profiles at each sequence position ($\text{delta}_i$) by computing the average difference between the wildtype and allele-specific enhancer scores. (B) We find that known TFBSs correspond to delta score peaks (red bars; enhancer-damaging) and dips (blue bars; enhancer-strengthening). To identify known and unknown TFBSs (peaks/dips in gray bars) directly from delta scores, we train a second model to annotate each nucleotide position as a peak, dip, or neutral using delta score profiles. TF-1 depicts a novel predicted peak active site (PAS). TF-2 depicts a novel predicted dip active site (DAS). The delta scores in this plot are for illustrative purposes only.

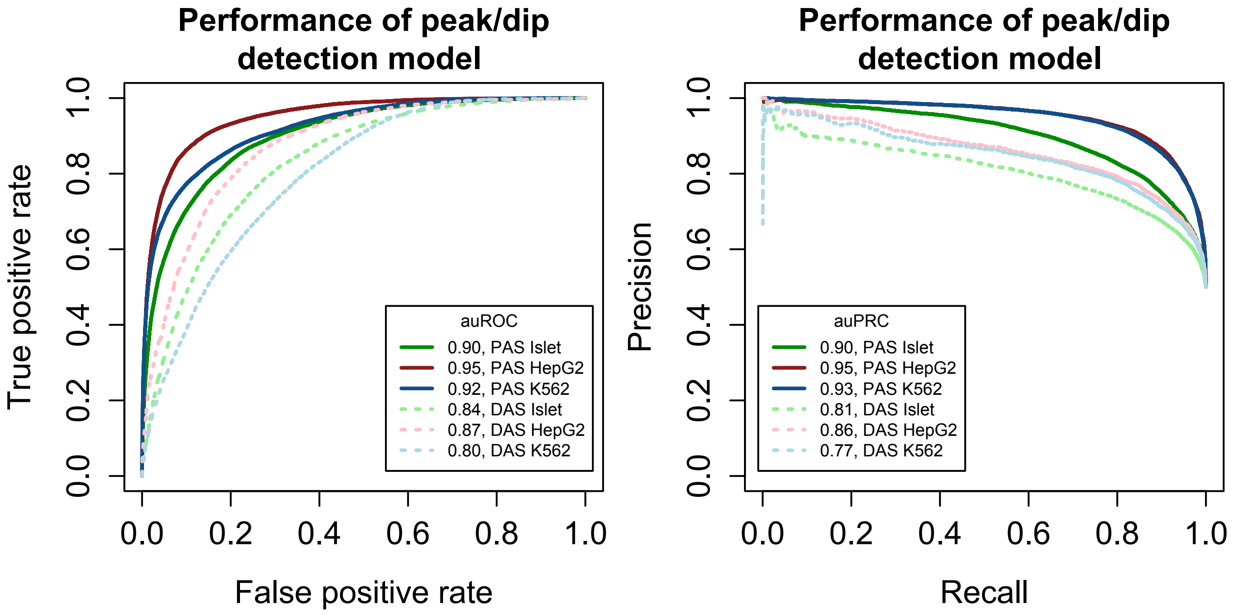


**Fig. S4. Performance of peak and dip detection models.** TFBSs detection performance of peak/dip models (linetype) for each biospecimen (color). Area under the curve listed in the plot legend for receiver operating characteristic (left) and precision recall curves (right).

**(A)** Examples of PASs and DASs at a locus

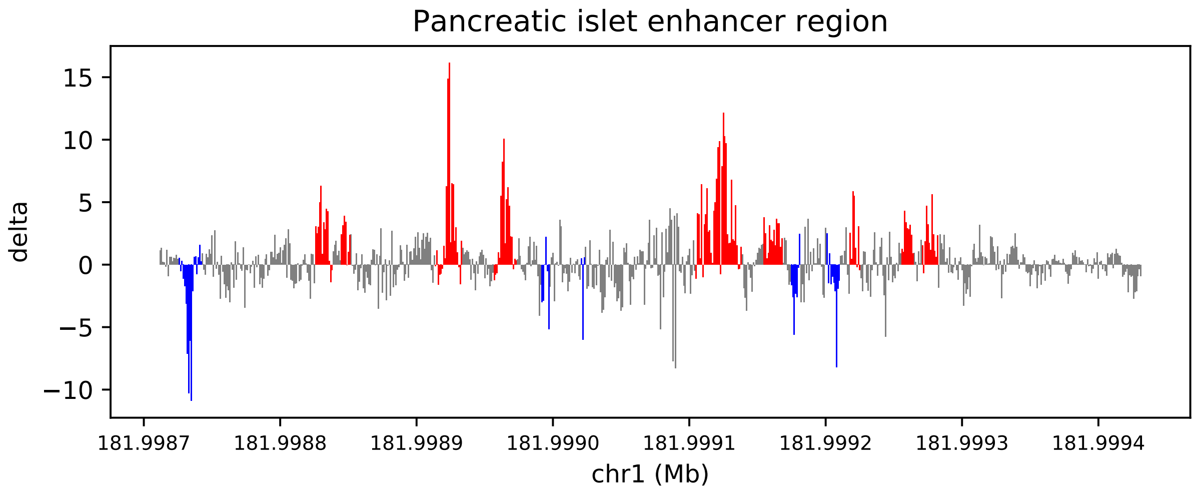


**(B)** Intersection of PASs and DASs

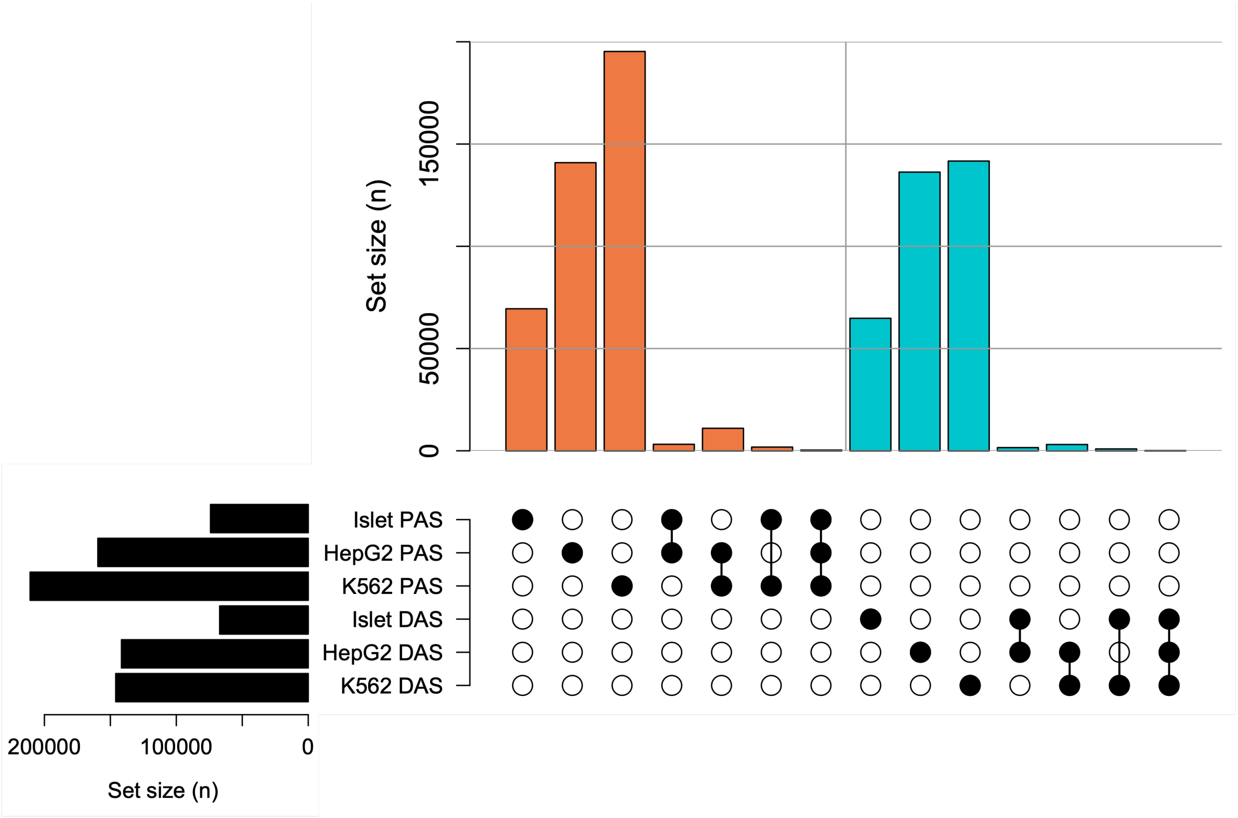


**Fig. S5. PAS and DAS results.** (A) Example islet PASs (blue) and DASs (red) overlaid on the corresponding delta scores (y-axis) in an islet enhancer region (x-axis). (B) Comparison of the overlap ($>= 1$ shared bp) of PASs (orange) and DASs (blue) across biospecimens, where set groups are shown in bubble plots (bottom).

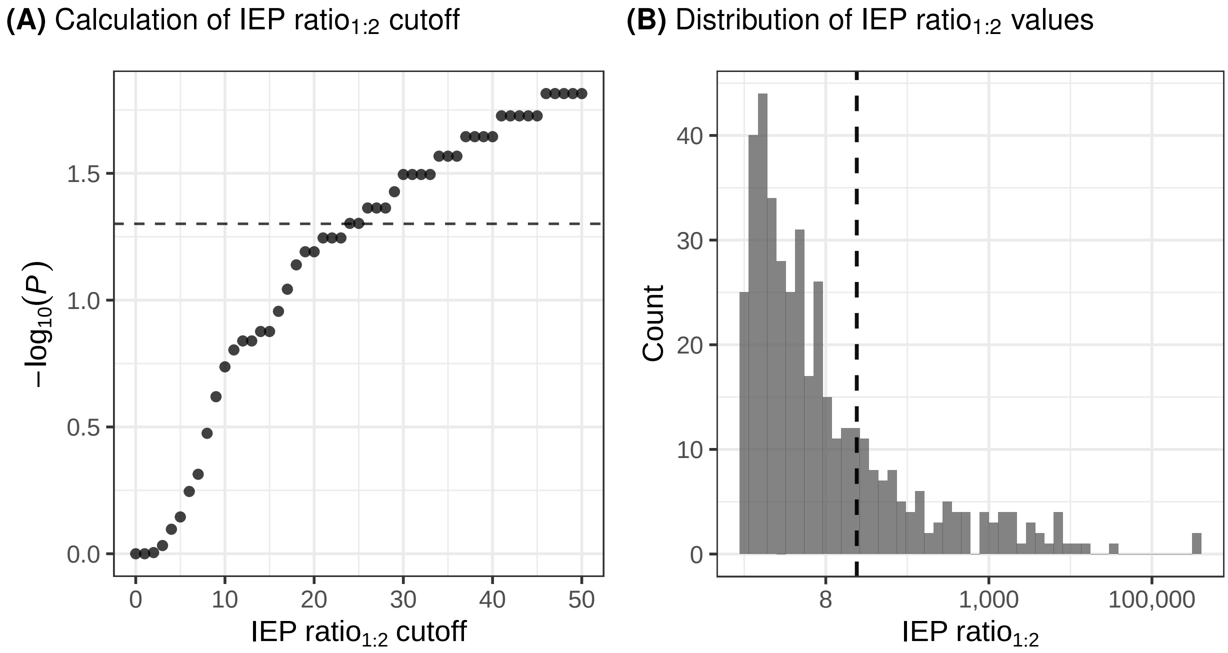


**Fig. S6. IEP $\text{ratio}_{1:2}$ prioritization cutoff.** (A) *P*-values from hypergeometric test (y-axis) evaluating the enrichment of SNPs prioritized by increasing IEP $\text{ratio}_{1:2}$ cutoffs (x-axis) from 99% European T2D credible sets in 99% trans-ancestry T2D credible sets at signals where $> 1$ SNPs are in the 99% European credible set and exactly 1 SNP occurs in the corresponding 99% trans-ancestry credible set. Dashed line at $P = 0.05$. (B) Distribution of IEP $\text{ratio}_{1:2}$ values (x-axis) for each signal in the 99% credible set for all disease/traits considered. Dashed line indicates cutoff derived from panel A.

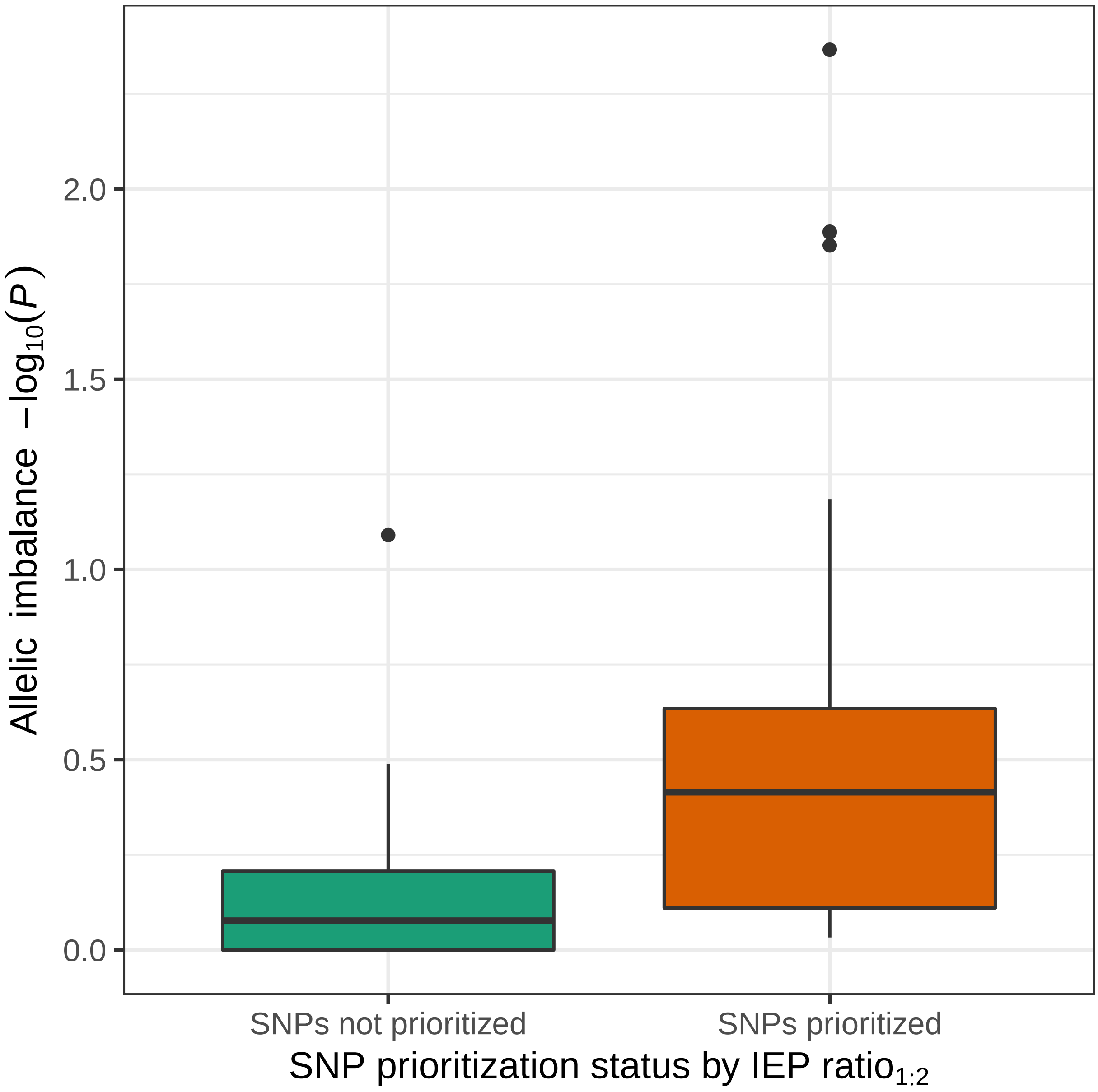


**Fig. S7. Allelic imbalance of IEP ratio$_{1:2}$ prioritized SNPs.** Comparison of the distribution of allelic imbalance *P*-values (y-axis) from islet ATAC-seq in heterozygous individuals for IEP ratio$_{1:2}$ prioritized SNPs (x-axis; orange) and all other SNPs in the 99% credible set (x-axis; green) at association signals where IEP ratio$_{1:2}$ prioritization identified one candidate causal SNP.

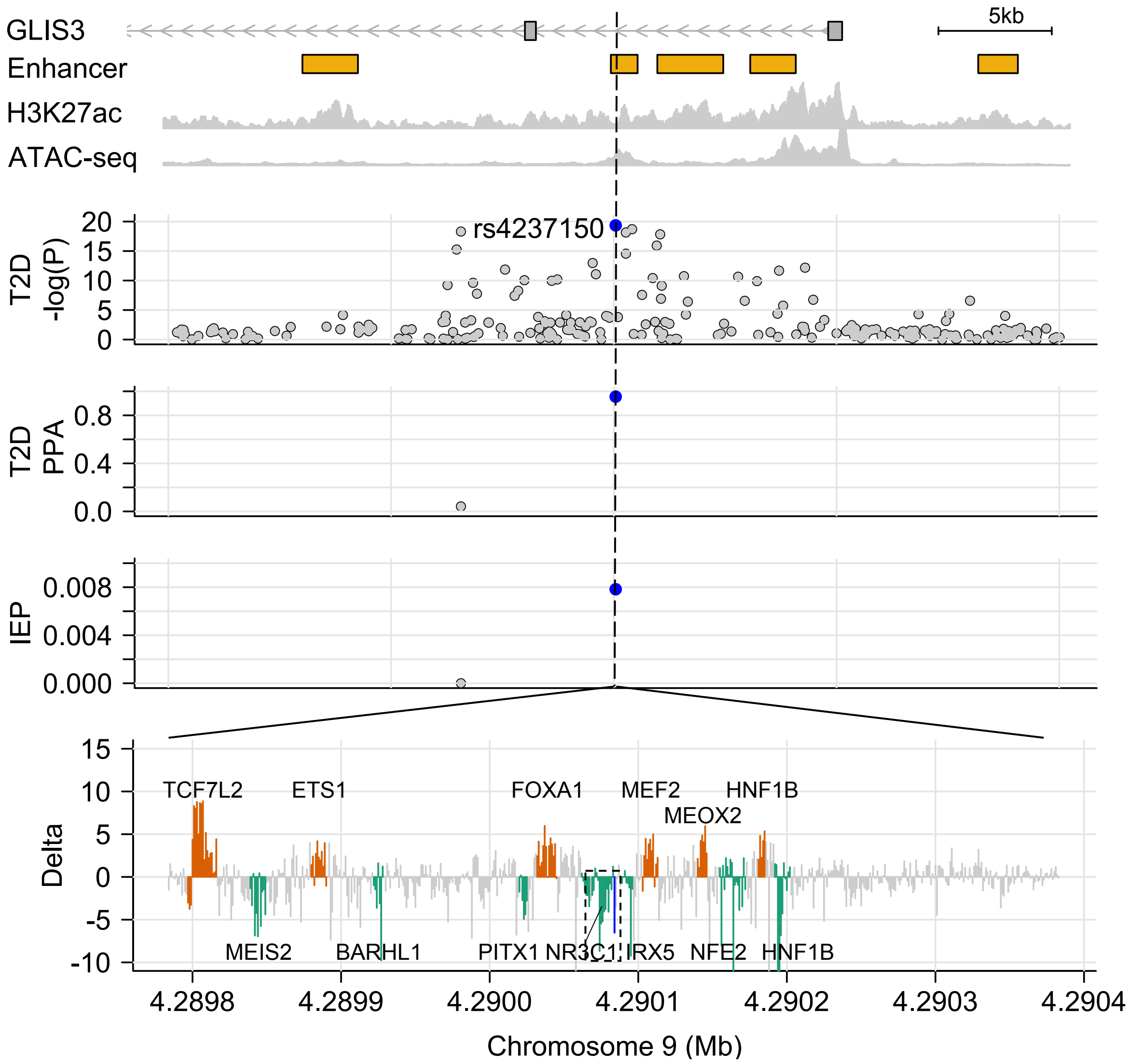


**Fig. S8.** ***GLIS3* locus.** Locus zoom around the 9:4290085 T2D association (T2D $-log_{10}(P)$ facet) in a *GLIS3* intron. Top facet shows islet enhancers, called from islet H3K27ac ChIP-seq and ATAC-seq data. rs4237150 (blue) is one of two SNPs in the 99% T2D credible set (PPA facet), has a large IEP score (IEP facet), and occurs in a DAS region (green; Delta facet). Dashed box indicates the DAS containing the candidate SNP (blue line).

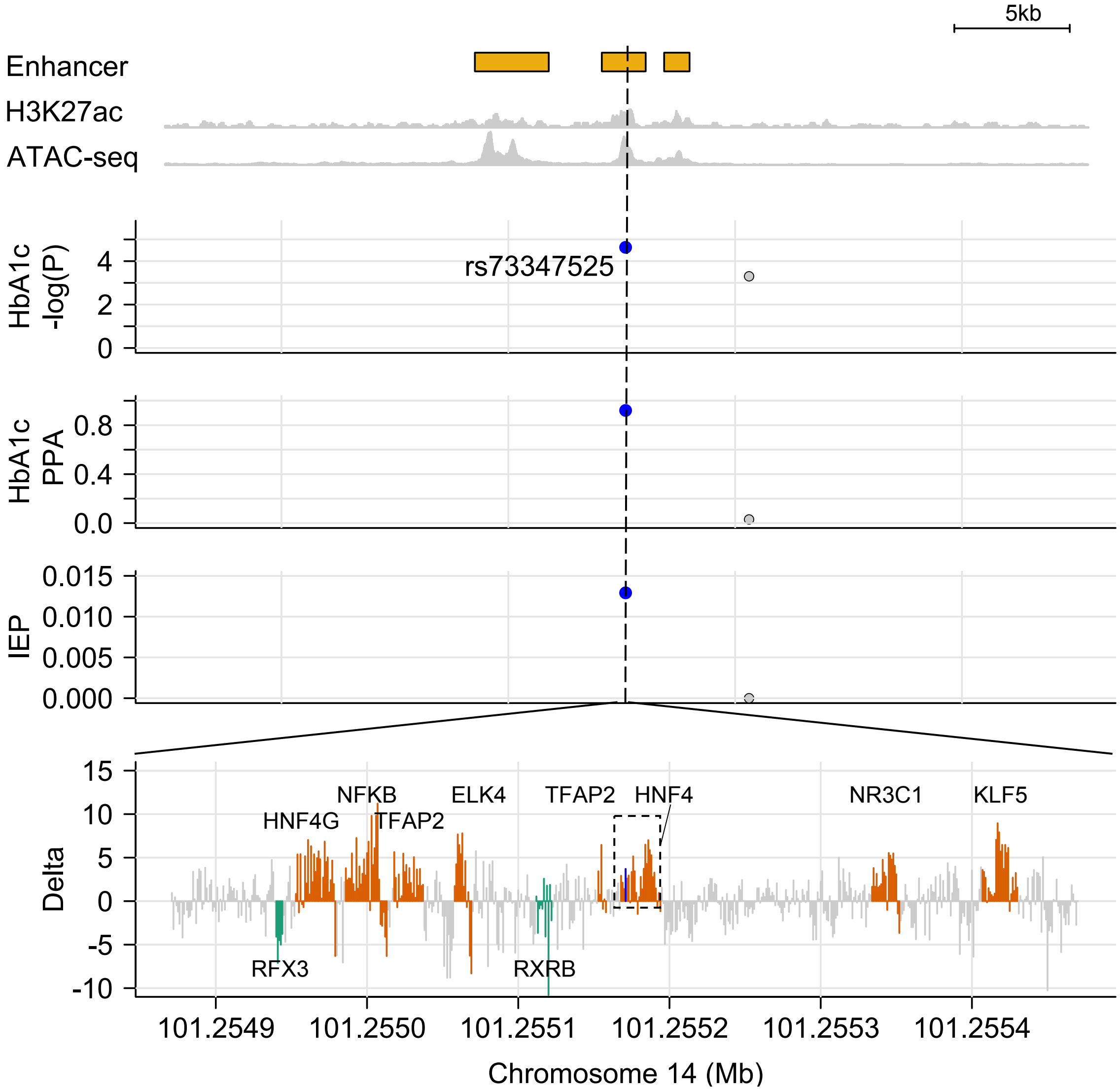


**Fig. S9. *DLK1* locus.** Locus zoom around the 14:101255172 HbA1C association (HbA1C $-log_{10}(P)$ facet), near *DLK1* which is outside of the genomic coordinates shown. Top facet shows islet enhancers, called from islet H3K27ac ChIP-seq and ATAC-seq data. rs73347525 (blue) is one of the 99% HbA1C credible set SNPs (PPA facet), has a large IEP score (IEP facet), and occurs in a PAS region (orange; Delta facet). Dashed box indicates the PAS containing the candidate SNP (blue line).

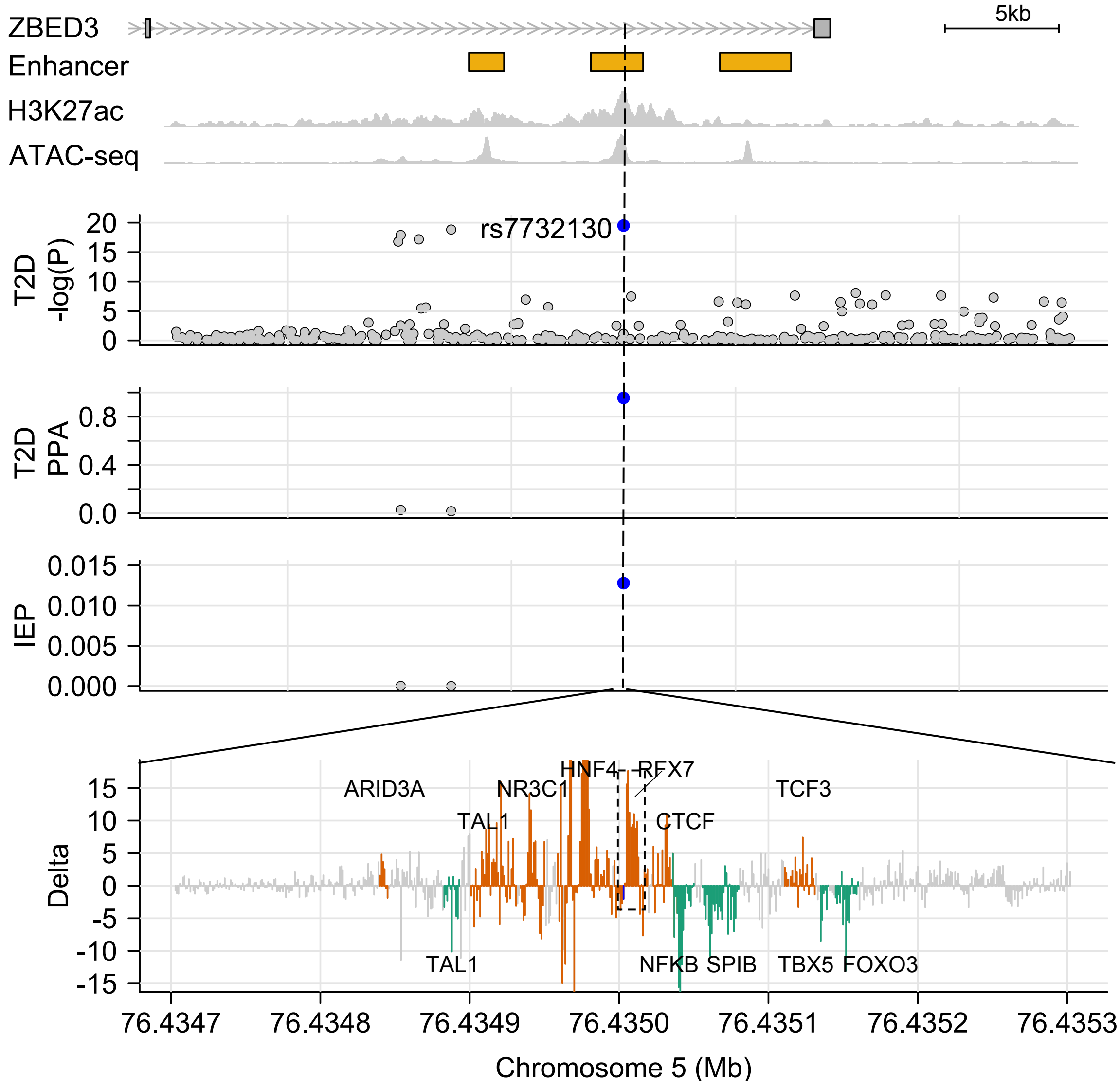


**Fig. S10.** ***ZBED3* locus.** Locus zoom around the 5:76435004 T2D association (T2D $-log_{10}(P)$ facet), at *ZBED3*. Top facet shows islet enhancers, called from islet H3K27ac ChIP-seq and ATAC-seq data. rs7732130 (blue) is one of the 99% T2D credible set SNPs (PPA facet), has a large IEP score (IEP facet), and occurs in a PAS region (orange; Delta facet). Dashed box indicates the PAS containing the candidate SNP (blue line).

| TFBS | Average delta in TFBS motif | Average delta in TFBS flank | $\text{Ratio}_{motif:flank}$ |
|---|---|---|---|
| TCF7L2:BACH2:TRIM28:HMGN3 | 9.0469400 | 0.690110 | 13.109400 |
| FOXA:FOX:FOXC2:FOXC1:FOXF2 | 2.7187200 | 0.274605 | 9.900480 |
| CEBPE:CEBPG:CEBPA:CEBPB | 5.9105600 | 0.644939 | 9.164530 |
| HNF1:HNF1A:HNF1B | 1.4904900 | 0.246813 | 6.038940 |
| DBP | 2.7862300 | 0.548098 | 5.083450 |
| MEF2 | 1.8211600 | 0.365245 | 4.986130 |
| NFE2L2 | 2.8760800 | 0.623274 | 4.614470 |
| GATA | 1.9332000 | 0.454534 | 4.253150 |
| MAFF:MAFG | 2.3234800 | 0.651011 | 3.569030 |
| TCF21:MYF6:MSC:ASCL2:MYOG | 2.0969800 | 0.594118 | 3.529570 |
| NFE2 | 1.8015000 | 0.522697 | 3.446550 |
| KLF16 | 1.3807200 | 0.407841 | 3.385440 |
| ATF4 | 2.4149100 | 0.717540 | 3.365540 |
| IRF | 1.2652200 | 0.386515 | 3.273400 |
| STAT | 1.3456300 | 0.412231 | 3.264260 |
| TEAD4:TEAD1:TEAD3 | 1.8860700 | 0.580423 | 3.249470 |
| EP300 | 1.3889300 | 0.448923 | 3.093920 |
| MYC | 1.5561800 | 0.506209 | 3.074180 |
| BCL | 1.4166200 | 0.481807 | 2.940220 |
| TCF12 | 1.5869300 | 0.542909 | 2.923010 |
| AP1 | 1.3254800 | 0.455766 | 2.908250 |
| MAF | 1.4159600 | 0.490977 | 2.883960 |
| HDAC2 | 0.9255450 | 0.384367 | 2.407970 |
| TFAP2 | 1.2330000 | 0.533990 | 2.309030 |
| ZNF740 | 0.8753080 | 0.384343 | 2.277410 |
| RXRA | 1.0180400 | 0.448128 | 2.271760 |
| BHLHA15:OLIG2 | 0.4721690 | 0.211981 | 2.227410 |
| MYF | 1.2892200 | 0.586876 | 2.196750 |
| TAL1 | 1.1057300 | 0.508764 | 2.173370 |
| NR3C1 | 0.9831560 | 0.458071 | 2.146300 |
| GTF2I | 0.8764100 | 0.409337 | 2.141050 |
| MXI1 | 1.1488400 | 0.548443 | 2.094730 |
| RFX5 | 1.0467800 | 0.499957 | 2.093740 |
| TFCP2 | 1.0914500 | 0.522462 | 2.089050 |
| CACD | 0.9915670 | 0.482966 | 2.053080 |
| NRL | 1.2814000 | 0.636737 | 2.012450 |
| OVOL2 | 0.8429610 | 0.454799 | 1.853480 |
| SREBP | 0.9882180 | 0.533657 | 1.851780 |
| SPI1 | 0.8629100 | 0.468536 | 1.841720 |
| TATA | 0.7920010 | 0.440804 | 1.796720 |
| ELF1 | 0.9690190 | 0.551094 | 1.758360 |
| ETS | 0.8607650 | 0.493842 | 1.743000 |
| EGR1 | 0.8005320 | 0.488085 | 1.640150 |
| ATF3 | 0.9116230 | 0.564967 | 1.613590 |
| CHD2 | 0.8544780 | 0.536357 | 1.593110 |
| E2F | 0.8143250 | 0.516242 | 1.577410 |
| HMGA1 | 0.5886910 | 0.379431 | 1.551510 |
| SMC3 | 0.7754170 | 0.508466 | 1.525010 |
| ZFX | 0.8442480 | 0.560772 | 1.505510 |
| SIRT6 | 0.9468310 | 0.629131 | 1.504980 |
| HNF4 | 0.6240370 | 0.417979 | 1.492990 |

| | | | |
|---|---|---|---|
| SOX17 | 0.4695030 | 0.316569 | 1.483100 |
| SP1 | 0.6308790 | 0.427328 | 1.476330 |
| HEY1 | 0.8010530 | 0.546259 | 1.466430 |
| SRF | 0.7508040 | 0.515783 | 1.455660 |
| MZF1 | 0.5786280 | 0.407201 | 1.420990 |
| BHLHE40 | 0.7330760 | 0.519300 | 1.411660 |
| POU2F2 | 0.6433150 | 0.456267 | 1.409950 |
| NANOG | 0.6906920 | 0.493835 | 1.398630 |
| SIN3A | 0.7477220 | 0.535425 | 1.396500 |
| PKNOX2:TGIF2LX:TGIF2 | 0.6049670 | 0.439624 | 1.376100 |
| SOX9 | 0.5003520 | 0.366266 | 1.366090 |
| TFCP2L1 | 0.8484700 | 0.625706 | 1.356020 |
| CTCF:RAD21 | 0.6552790 | 0.486918 | 1.345770 |
| REST | 0.6843840 | 0.512284 | 1.335950 |
| PAX5 | 0.6703400 | 0.510149 | 1.314010 |
| RUNX2 | 0.6509740 | 0.498049 | 1.307050 |
| ESRRA | 0.6222210 | 0.482093 | 1.290670 |
| REL | 0.6279390 | 0.504035 | 1.245820 |
| YY1 | 0.6264430 | 0.512010 | 1.223500 |
| ZNF143 | 0.6017220 | 0.504918 | 1.191720 |
| EBF1 | 0.5783750 | 0.494279 | 1.170140 |
| HIC1 | 0.6316610 | 0.544796 | 1.159450 |
| NFKB | 0.5295010 | 0.460065 | 1.150930 |
| PAX2 | 0.5726050 | 0.519630 | 1.101950 |
| HEY2 | 0.5197700 | 0.488618 | 1.063760 |
| ZIC2:ZIC1:ZIC3 | 0.5044810 | 0.487959 | 1.033860 |
| NKX2-5 | 0.3991450 | 0.391284 | 1.020090 |
| RHOXF1 | 0.4223190 | 0.424415 | 0.995061 |
| GLI | 0.4230810 | 0.425236 | 0.994932 |
| POU5F1 | 0.2750300 | 0.300068 | 0.916559 |
| SETDB1 | 0.4876430 | 0.533604 | 0.913867 |
| IKZF1 | 0.4340330 | 0.476024 | 0.911788 |
| PBX3 | 0.4051790 | 0.473039 | 0.856545 |
| GCM2:GCM1 | 0.3609310 | 0.446948 | 0.807546 |
| HOXA7 | 0.2466590 | 0.308423 | 0.799743 |
| SOX14 | 0.1939000 | 0.324805 | 0.596974 |
| EN1 | 0.1833610 | 0.363579 | 0.504322 |
| RBPJ | 0.1486800 | 0.358928 | 0.414233 |
| IKZF2 | 0.1660080 | 0.407663 | 0.407219 |
| SIX5 | 0.0604405 | 0.288187 | 0.209727 |
| ZBTB7C:ZBTB7A | 0.0754125 | 0.439864 | 0.171445 |
| ZNF410 | -0.0111956 | 0.334417 | -0.033478 |
| CUX1 | -0.0431143 | 0.372399 | -0.115774 |
| ARID5B | -0.0680515 | 0.303808 | -0.223995 |
| DOBOX4 | -0.1748180 | 0.282215 | -0.619450 |
| PBX1 | -0.2992370 | 0.340565 | -0.878649 |
| YY2 | -0.8091360 | 0.488708 | -1.655660 |
| ONECUT | -0.3668550 | 0.200166 | -1.832750 |
| AP3 | -0.5095790 | 0.141112 | -3.611170 |

**Table S1. Prioritization of islet relevant transcription factors using delta scores.** Islet TF footprints ranked by the ratio of the average delta score within the TF footprint motif to the flanking region. TFBS refers to the predicted TF binding site from islet TF footprints. TFs with similar PWMs are merged (Methods) and separated by ":" in the TFBS column.

| Disease or trait | Signal | Gene | N SNP | IEP top SNP | IEP 2nd top SNP | IEP score ratio | IEP score top SNP | IEP score 2nd top SNP | IEP percentile top SNP | IEP percentile 2nd top SNP |
|---|---|---|---|---|---|---|---|---|---|---|
| T2D | chr9:84308948 | TLE1 | 2 | rs2796441 | rs9410573 | Inf | 1.13E-02 | 0.00E+00 | 0.984 | 0.000 |
| T2D | chr15:38873115 | RASGRP1 | 2 | rs12912777 | rs34715063 | 355601.64 | 1.94E-03 | 5.46E-09 | 0.943 | 0.058 |
| HbA1c | chr6:33796794-53507100_4 | GLP1R | 2 | rs10305514 | rs10305518 | 37912.17 | 2.40E-03 | 6.32E-08 | 0.949 | 0.239 |
| HbA1c | chr14:91785258-105440006_4 | DLK1 | 2 | rs73347525 | rs8004581 | 16732.11 | 1.29E-02 | 7.72E-07 | 0.986 | 0.429 |
| HbA1c | chr2:18233271-37277241_2 | VIT | 2 | rs10206462 | rs2691106 | 12031.96 | 5.54E-04 | 4.60E-08 | 0.896 | 0.213 |
| HbA1c | chr20:22375756-40249273_1 | PXMP4 | 3 | rs13042148 | rs149142833 | 9024.43 | 4.83E-03 | 5.35E-07 | 0.967 | 0.401 |
| Glucose | chr7:40687437-51293550_2 | GCK | 3 | rs2908292 | rs2971672 | 7954.92 | 3.71E-03 | 4.66E-07 | 0.961 | 0.391 |
| HbA1c | chr5:71716874-85560931_2 | ZNF366 | 3 | rs35585881 | rs34216626 | 6931.32 | 1.07E-03 | 1.55E-07 | 0.923 | 0.309 |
| Glucose | chr14:80252374-99785610_1 | FOXN3 | 2 | rs35889227 | rs11626777 | 6383.12 | 1.36E-02 | 2.13E-06 | 0.987 | 0.506 |
| HbA1c | chr2:36365383-55151959_1 | SIX3 | 3 | rs2121564 | rs10205222 | 6262.30 | 2.78E-04 | 4.45E-08 | 0.863 | 0.210 |
| HbA1c | chr12:38542595-57976118_8 | SENP1 | 2 | rs117797076 | rs117523200 | 4999.69 | 2.10E-05 | 4.20E-09 | 0.687 | 0.048 |
| T2D | chr9:4290085 | GLIS3 | 2 | rs4237150 | rs1574285 | 4211.41 | 7.83E-03 | 1.86E-06 | 0.977 | 0.495 |
| HbA1c | chr4:80561055-96171283_1 | ABCG2 | 8 | rs45499402 | rs4148155 | 4041.16 | 2.38E-02 | 5.89E-06 | 0.993 | 0.585 |
| T2D | chr3:54828827 | CACNA2D3 | 3 | rs75088635 | rs111494834 | 3166.90 | 3.53E-03 | 1.11E-06 | 0.959 | 0.456 |
| HbA1c | chr17:57914080-77759691_4 | CCDC47 | 4 | rs75646162 | rs72845888 | 2943.81 | 1.53E-02 | 5.19E-06 | 0.988 | 0.575 |
| HbA1c | chr3:175462743-186763651_2 | ST6GAL1 | 2 | rs6780016 | rs3936289 | 2887.83 | 4.71E-05 | 1.63E-08 | 0.749 | 0.124 |
| HbA1c | chr11:221870-17419500_10 | LMO1 | 2 | rs4758317 | rs110420 | 2339.79 | 1.31E-02 | 5.59E-06 | 0.986 | 0.581 |
| HbA1c | chr2:161007292-178850138_6 | ABCB11 | 2 | rs478333 | rs2947987 | 2154.15 | 1.14E-05 | 5.30E-09 | 0.638 | 0.057 |
| HbA1c | chr16:7769193-24862718_2 | ABCC1 | 3 | rs504348 | rs71378214 | 2066.12 | 1.12E-03 | 5.41E-07 | 0.924 | 0.402 |
| T2D | chr9:22301092 | CDKN2A-CDKN2B | 4 | rs1575972 | rs7045760 | 1964.45 | 4.10E-03 | 2.09E-06 | 0.963 | 0.504 |
| HbA1c | chr12:38542595-57976118_7 | R3HDM2 | 4 | rs4760278 | rs7484541 | 1811.16 | 2.60E-03 | 1.43E-06 | 0.951 | 0.475 |
| Fasting glucose | rs17168486 | DGKB | 2 | rs17168486 | rs11980500 | 1553.35 | 1.21E-03 | 7.79E-07 | 0.927 | 0.429 |
| HbA1c | chr10:3136070-21712524_1 | CDC123 | 4 | rs7394200 | rs7077792 | 1433.94 | 1.96E-05 | 1.37E-08 | 0.682 | 0.111 |
| HbA1c | chr19:360319-10567212_4 | ARHGAP45 | 3 | rs12974537 | rs35532684 | 1358.22 | 1.81E-05 | 1.33E-08 | 0.675 | 0.109 |
| T2D | chr4:6293237 | WFS1 | 36 | rs4234731 | rs13103357 | 1324.54 | 9.24E-03 | 6.98E-06 | 0.981 | 0.599 |
| HbA1c | chr3:4803184-21599247_3 | PPARG | 2 | rs4518111 | rs4135247 | 1268.03 | 4.45E-05 | 3.51E-08 | 0.745 | 0.189 |
| HbA1c | chr7:36762307-52275656_3 | GCK | 4 | rs2908292 | rs2971671 | 1113.73 | 3.71E-03 | 3.33E-06 | 0.961 | 0.540 |
| HbA1c | chr14:80124163-99783600_1 | FOXN3 | 3 | rs35889227 | rs10873398 | 1047.29 | 1.36E-02 | 1.30E-05 | 0.987 | 0.649 |
| T2D | chr13:54107583 | OLFM4 | 5 | rs12429545 | rs4477562 | 863.72 | 7.41E-03 | 8.58E-06 | 0.976 | 0.615 |
| Glucose | chr5:67102574-81961482_1 | ZBED3 | 3 | rs7732130 | rs4457054 | 797.58 | 1.28E-02 | 1.60E-05 | 0.986 | 0.666 |
| HbA1c | chr5:67098268-86045601_1 | ZBED3 | 3 | rs7732130 | rs4457054 | 797.58 | 1.28E-02 | 1.60E-05 | 0.986 | 0.666 |
| T2D | chr5:76435004 | ZBED3 | 3 | rs7732130 | rs4457054 | 797.58 | 1.28E-02 | 1.60E-05 | 0.986 | 0.666 |
| HbA1c | chr12:459593-14109457_6 | LTBR | 3 | rs10466905 | rs12296430 | 519.65 | 9.09E-04 | 1.75E-06 | 0.916 | 0.491 |
| HbA1c | chr1:149784689-167872017_5 | OR6Y1 | 2 | rs189857927 | rs555280966 | 501.78 | 3.09E-05 | 6.16E-08 | 0.717 | 0.237 |
| T2D | chr3:124921457 | SLC12A8 | 3 | rs649961 | rs569255 | 480.04 | 1.94E-02 | 4.03E-05 | 0.991 | 0.738 |
| HbA1c | chr17:945530-13945508_2 | PFAS | 2 | rs2313286 | rs4791663 | 467.81 | 1.91E-04 | 4.08E-07 | 0.842 | 0.381 |

| | | | | | | | | | | |
|---|---|---|---|---|---|---|---|---|---|---|
| HbA1c | chr12:38542595-57976118_3 | PFKM | 2 | rs140895856 | rs139395863 | 445.34 | 2.12E-04 | 4.77E-07 | 0.848 | 0.393 |
| HbA1c | chr11:2140530-12207149_3 | PPFIBP2 | 16 | rs10743026 | rs7949659 | 411.52 | 3.21E-02 | 7.81E-05 | 0.996 | 0.785 |
| HbA1c | chr3:56856309-74768389_3 | SHQ1 | 3 | rs13080180 | rs13085136 | 396.56 | 1.43E-02 | 3.61E-05 | 0.988 | 0.729 |
| HbA1c | chr2:233963437-241496445_2 | ILKAP | 4 | rs10929270 | rs4663862 | 378.01 | 3.41E-05 | 9.02E-08 | 0.725 | 0.267 |
| T2D | chr12:4521511 | CCND2 | 7 | 12:4288000:T:C | 12:4301300:C:T | 343.43 | 3.96E-04 | 1.15E-06 | 0.880 | 0.459 |
| HbA1c | chr7:1011402-15893576_1 | FAM220A | 3 | rs13226769 | rs35076783 | 335.44 | 1.05E-05 | 3.14E-08 | 0.632 | 0.179 |
| HbA1c | chr11:55267156-69977915_4 | MYEOV | 2 | rs148893083 | rs188584129 | 334.73 | 1.07E-05 | 3.18E-08 | 0.633 | 0.181 |
| HbA1c | chr2:165514409-184356242_2 | SCRN3 | 3 | rs10203361 | rs7570614 | 288.39 | 9.40E-05 | 3.26E-07 | 0.798 | 0.364 |
| HbA1c | chr7:18200482-36754918_3 | KIAA0087 | 15 | rs2286177 | rs62446872 | 282.91 | 1.54E-02 | 5.45E-05 | 0.989 | 0.760 |
| T2D | chr10:114552267 | TCF7L2 | 4 | rs10787461 | rs941826 | 232.86 | 2.34E-02 | 1.01E-04 | 0.993 | 0.802 |
| HbA1c | chr6:125423968-144260129_1 | HBS1L | 5 | rs7776054 | rs9389268 | 214.26 | 4.71E-05 | 2.20E-07 | 0.749 | 0.335 |
| T2D | chr3:23457080 | UBE2E2 | 6 | rs17012829 | rs11926494 | 212.81 | 2.42E-03 | 1.14E-05 | 0.949 | 0.638 |
| HbA1c | chr17:51634879-67464569_1 | YPEL2 | 2 | rs12600858 | rs2060779 | 194.59 | 5.05E-03 | 2.60E-05 | 0.968 | 0.704 |
| HbA1c | chr5:148867578-168166431_2 | TIMD4 | 11 | rs12657266 | rs58198139 | 190.15 | 6.03E-04 | 3.17E-06 | 0.900 | 0.536 |
| T2D | chr10:114715598 | TCF7L2 | 3 | rs2104598 | rs11196169 | 161.36 | 1.53E-02 | 9.48E-05 | 0.989 | 0.799 |
| HbA1c | chr19:41748220-58664418_2 | APOC1 | 2 | rs584007 | rs3826688 | 150.44 | 2.71E-03 | 1.80E-05 | 0.952 | 0.675 |
| T2D | chr6:40409243 | LRFN2 | 2 | rs34298980 | rs34045288 | 146.10 | 1.29E-03 | 8.83E-06 | 0.929 | 0.618 |
| T2D | chr10:114344288 | TCF7L2 | 2 | rs12243296 | rs192497142 | 137.33 | 4.70E-03 | 3.42E-05 | 0.966 | 0.725 |
| HbA1c | chr17:7672395-27554378_5 | PFAS | 4 | rs10468467 | rs9891267 | 134.12 | 1.78E-06 | 1.33E-08 | 0.492 | 0.109 |
| Glucose | chr11:400109-17518525_2 | PSMA1 | 4 | rs78959242 | rs76277448 | 131.26 | 4.84E-06 | 3.69E-08 | 0.570 | 0.194 |
| HbA1c | chr17:71006408-81049792_1 | FN3K | 7 | rs7208565 | rs12947062 | 122.32 | 1.86E-04 | 1.52E-06 | 0.841 | 0.480 |
| HbA1c | chr1:149784689-167872017_2 | HORMAD1 | 2 | rs56057831 | rs77209899 | 121.40 | 1.24E-06 | 1.03E-08 | 0.465 | 0.091 |
| T2D | chr17:36056076 | HNF1B | 4 | rs12938438 | rs10962 | 106.74 | 7.17E-05 | 6.71E-07 | 0.779 | 0.418 |
| HbA1c | chr1:16516110-33342430_4 | RPS6KA1 | 3 | rs4970489 | rs4443935 | 100.87 | 2.20E-04 | 2.18E-06 | 0.850 | 0.507 |
| HbA1c | chr13:21251235-36045677_3 | LINC00426 | 4 | rs1337939 | rs1337940 | 94.26 | 2.15E-03 | 2.28E-05 | 0.946 | 0.694 |
| T2D | chr2:234191103 | ATG16L1-DGKD | 4 | rs117809958 | rs117447187 | 94.23 | 1.25E-04 | 1.33E-06 | 0.816 | 0.470 |
| HbA1c | chr9:126208837-140199175_2 | GFI1B | 2 | rs73554552 | rs60757417 | 86.86 | 1.37E-05 | 1.58E-07 | 0.653 | 0.310 |
| T2D | chr14:79944099 | NRXN3 | 11 | rs7156625 | rs7144011 | 81.07 | 2.02E-02 | 2.49E-04 | 0.992 | 0.857 |
| T2D | chr6:164133001 | QKI | 7 | rs4709746 | rs17630640 | 76.22 | 3.55E-02 | 4.66E-04 | 0.996 | 0.888 |
| HbA1c | chr17:945530-13945508_3 | SMG6 | 2 | rs7213347 | rs2873195 | 69.22 | 1.47E-05 | 2.13E-07 | 0.659 | 0.332 |
| HbA1c | chr6:100884618-118992323_1 | SLC22A16 | 2 | rs72939920 | rs12194000 | 65.60 | 1.08E-06 | 1.64E-08 | 0.454 | 0.125 |
| T2D | chr10:71466578 | VPS26A-NEUROG3 | 15 | rs2616132 | rs10762307 | 64.54 | 1.84E-02 | 2.85E-04 | 0.991 | 0.864 |
| HbA1c | chr19:36748817-54972918_1 | CD33 | 3 | rs34813869 | rs1354106 | 63.33 | 2.74E-04 | 4.32E-06 | 0.862 | 0.561 |
| T2D | chr6:50788778 | TFAP2B | 3 | rs3798519 | rs2206277 | 59.67 | 3.57E-04 | 5.99E-06 | 0.875 | 0.587 |
| HbA1c | chr13:107220834-115107493_4 | ATP11A | 3 | rs142329603 | rs113245682 | 57.64 | 4.90E-04 | 8.49E-06 | 0.890 | 0.614 |
| Glucose | chr11:7117503-23425585_1 | PSMA1 | 3 | rs75336838 | rs117720468 | 57.39 | 1.29E-03 | 2.25E-05 | 0.929 | 0.692 |
| HbA1c | chr11:7598559-21202416_2 | PSMA1 | 5 | rs75336838 | rs117720468 | 57.39 | 1.29E-03 | 2.25E-05 | 0.929 | 0.692 |
| HbA1c | chr2:36863789-52883790_2 | FBXO11 | 34 | rs74646951 | rs11687989 | 54.55 | 1.62E-03 | 2.97E-05 | 0.937 | 0.714 |
| T2D | chr10:122834572 | WDR11 | 3 | rs11199755 | rs11199753 | 49.28 | 7.87E-04 | 1.60E-05 | 0.911 | 0.665 |

| | | | | | | | | | | |
|---|---|---|---|---|---|---|---|---|---|---|
| T2D | chr7:127250831 | GCC1-PAX4-LEP | 3 | rs72607746 | rs12669223 | 45.97 | 4.91E-04 | 1.07E-05 | 0.890 | 0.633 |
| T2D | chr11:128040810 | ETS1 | 14 | rs7933438 | rs7931773 | 45.76 | 1.10E-01 | 2.41E-03 | 1.000 | 0.949 |
| HbA1c | chr17:51634879-67464569_7 | YPEL2 | 9 | rs112412433 | rs112197279 | 45.16 | 2.14E-03 | 4.74E-05 | 0.946 | 0.750 |
| HbA1c | chr5:87068177-103510995_4 | GLRX | 6 | rs2546197 | rs12523597 | 44.93 | 5.14E-03 | 1.14E-04 | 0.969 | 0.811 |
| Glucose | chr11:365897-11800779_1 | KCNQ1 | 2 | rs4930011 | rs234864 | 44.51 | 5.00E-03 | 1.12E-04 | 0.968 | 0.810 |
| HbA1c | chr16:1484338-20380004_4 | ABCC1 | 2 | rs184499898 | rs45623833 | 42.36 | 3.47E-07 | 8.19E-09 | 0.369 | 0.078 |
| HbA1c | chr3:117101604-133501529_1 | SLC12A8 | 2 | rs1416218 | rs684030 | 41.23 | 6.74E-03 | 1.64E-04 | 0.974 | 0.833 |
| HbA1c | chr3:39480518-57952834_3 | CDHR4 | 3 | rs114424909 | rs142613277 | 39.62 | 2.94E-03 | 7.43E-05 | 0.955 | 0.782 |
| HbA1c | chr19:35485269-54814602_3 | SIGLEC5 | 2 | rs73050880 | rs112456998 | 36.33 | 8.59E-08 | 2.36E-09 | 0.264 | 0.031 |
| Fasting glucose | rs17390909 | ELK3 | 5 | rs2268501 | rs17331697 | 36.17 | 8.59E-03 | 2.37E-04 | 0.979 | 0.854 |
| T2D | chr19:13038415 | FARSA-ZNF799 | 10 | rs2974752 | rs2242517 | 35.44 | 5.59E-03 | 1.58E-04 | 0.970 | 0.831 |
| Glucose | chr10:106575427-121378575_2 | TIAL1 | 8 | rs79612474 | rs41287142 | 35.28 | 1.46E-02 | 4.13E-04 | 0.988 | 0.882 |
| HbA1c | chr8:36842055-49957895_2 | ANK1 | 4 | rs72638983 | rs72638977 | 34.54 | 3.76E-04 | 1.09E-05 | 0.878 | 0.634 |
| HbA1c | chr2:36863789-52883790_4 | ZFP36L2 | 7 | rs112694524 | rs77552263 | 33.74 | 2.09E-03 | 6.21E-05 | 0.945 | 0.769 |
| T2D | chr4:106048291 | TET2 | 4 | rs11729069 | rs17035289 | 31.70 | 1.63E-03 | 5.15E-05 | 0.937 | 0.756 |
| HbA1c | chr3:176710191-196233136_1 | ST6GAL1 | 5 | rs1981767 | rs9814673 | 30.62 | 5.43E-03 | 1.77E-04 | 0.970 | 0.838 |
| HbA1c | chr7:17545963-33138472_2 | IGF2BP3 | 6 | rs12700421 | rs12700423 | 29.62 | 1.64E-02 | 5.52E-04 | 0.989 | 0.896 |
| HbA1c | chr19:41748220-58664418_5 | ZC3H4 | 4 | rs62136859 | rs1532127 | 29.42 | 4.50E-06 | 1.53E-07 | 0.564 | 0.308 |
| Glucose | chr19:36509463-54873286_2 | ZC3H4 | 6 | rs62136859 | rs1532127 | 29.42 | 4.50E-06 | 1.53E-07 | 0.564 | 0.308 |
| T2D | chr3:63897215 | PSMD6-ADAMTS9 | 10 | rs6785040 | rs2292662 | 29.13 | 7.08E-03 | 2.43E-04 | 0.975 | 0.855 |
| HbA1c | chr13:107606835-115109852_1 | ATP11A | 3 | rs76533333 | rs12876143 | 28.84 | 1.56E-03 | 5.39E-05 | 0.936 | 0.759 |
| HbA1c | chr20:33191665-51625096_3 | PLTP | 4 | rs12185776 | rs58847685 | 28.12 | 1.30E-04 | 4.63E-06 | 0.819 | 0.566 |
| T2D | chr2:227100490 | IRS1 | 29 | rs2943654 | rs2673128 | 27.72 | 4.97E-03 | 1.79E-04 | 0.968 | 0.838 |
| HbA1c | chr3:134658128-150395262_2 | ATP1B3 | 9 | rs11539489 | rs55704642 | 27.52 | 2.65E-03 | 9.64E-05 | 0.952 | 0.800 |
| T2D | chr3:170724883 | SLC2A2 | 9 | rs1905505 | rs6804915 | 25.75 | 7.94E-04 | 3.08E-05 | 0.911 | 0.717 |
| T2D | chr11:2856658 | INS-IGF2-KCNQ1 | 2 | rs4930011 | rs2237895 | 24.97 | 5.00E-03 | 2.00E-04 | 0.968 | 0.845 |

**Table S2. Signals where IEP ratio$_{1:2}$ refined 99% credible set SNPs to one SNP.** Signals with a single SNP identified by IEP ratio$_{1:2}$ that previously had $> 1$ SNPs in the 99% credible set. Gene column refers to the nearest gene except in the case of the T2D data, for which we used the gene reported by the original study. N SNP column refers to the number of SNPs in the original 99% credible set (uniform prior). Note: IEP percentiles are rounded.

| Number | SNP | Ref | Alt | High score allele | Ref score | Alt score | PAS/DAS FPR 5% | PAS/DAS FPR 10% | TF motif overlap | Increased binding allele | Log difference | Motif database |
|---|---|---|---|---|---|---|---|---|---|---|---|---|
| 1 | rs4970489 | T | C | T | 0.028478 | 0.020762 | DAS islet | DAS islet | | | | |
| | | | | | | | | DAS K562 | Gli2(Zf) | C | 3.096273503 | HOMER |
| | | | | | | | | DAS HepG2 | GLI3(Zf) | C | 2.835239687 | HOMER |
| | | | | | | | | | ESRRA_disc3 | C | 2.513480022 | ENCODE |
| | | | | | | | | | GLIS3(Zf) | C | 2.26659149 | HOMER |
| | | | | | | | | | ZIC3_1 | C | 2.240891524 | ENCODE |
| | | | | | | | | | GLI2_1 | C | 2.148906 | ENCODE |
| 2 | rs56057831 | A | T | A | 0.002942 | 0.002519 | DAS islet | DAS islet | | | | |
| | | | | | | | DAS K562 | DAS K562 | STAT_disc2 | A | 3.105875853 | ENCODE |
| | | | | | | | DAS HepG2 | DAS HepG2 | PRDM1_disc2 | A | 3.084881004 | ENCODE |
| | | | | | | | | | AP1_disc3 | A | 3.075774981 | ENCODE |
| | | | | | | | | | FOSL2::JUND | A | 2.481396335 | JASPAR |
| | | | | | | | | | Fos(bZIP) | A | 2.143113679 | HOMER |
| | | | | | | | | | JUN(var.2) | A | 2.058555651 | JASPAR |
| | | | | | | | | | FOS::JUN | A | 2.020873188 | JASPAR |
| 3 | rs189857927 | A | G | G | 0.030573 | 0.031552 | DAS K562 | DAS K562 | | | | |
| | | | | | | | DAS HepG2 | DAS HepG2 | FOXB1_3 | G | 1.732971395 | ENCODE |
| 4 | rs7394200 | T | C | T | 0.017668 | 0.016557 | | DAS islet | | | | |
| | | | | | | | | DAS K562 | IRF_known1 | T | 2.972475411 | ENCODE |
| | | | | | | | | DAS HepG2 | IRF_known3 | T | 2.642484247 | ENCODE |
| | | | | | | | | | IRF_known2 | T | 2.610411732 | ENCODE |
| | | | | | | | | | IRF_known21 | T | 2.574518808 | ENCODE |
| 5 | rs2616132 | G | A | G | 0.284545 | 0.219826 | DAS islet | DAS islet | | | | |
| | | | | | | | | DAS K562 | RFX3 | G | 2.69704899 | JASPAR |
| | | | | | | | | | TOPORS_1 | G | 2.405515474 | ENCODE |
| 6 | rs12243296 | A | G | G | 0.186022 | 0.208544 | DAS islet | DAS islet | | | | |
| | | | | | | | | DAS K562 | ZNF317(Zf) | G | 3.732861917 | HOMER |
| | | | | | | | | | MEIS2_4 | G | 2.496597974 | ENCODE |
| | | | | | | | | | MEIS3_5 | G | 2.47248413 | ENCODE |
| | | | | | | | | | SNAI1 | G | 2.467928313 | JASPAR |
| | | | | | | | | | MEIS3_3 | G | 2.458357833 | ENCODE |
| | | | | | | | | | TBX21_6 | G | 2.417424353 | ENCODE |
| | | | | | | | | | ZEB2(Zf) | G | 2.381653616 | HOMER |
| | | | | | | | | | ZSCAN16_1 | G | 2.283253636 | ENCODE |
| | | | | | | | | | SNAI2_1 | G | 2.004775326 | ENCODE |
| 7 | rs10787461 | A | G | A | 0.42461 | 0.369475 | PAS islet | PAS islet | | | | |
| | | | | | | | PAS HepG2 | PAS K562 | MYF_1 | G | 3.407552374 | ENCODE |
| | | | | | | | | PAS HepG2 | Sox17(HMG) | A | 3.2988296 | HOMER |
| | | | | | | | | | Oct4:Sox17(POU,Homeobox,HMG) | A | 3.252885479 | HOMER |
| | | | | | | | | | FOXJ3_3 | G | 2.850901326 | ENCODE |
| | | | | | | | | | FOXJ1_1 | A | 2.706519979 | ENCODE |
| | | | | | | | | | SOX17_4 | A | 2.670871198 | ENCODE |
| | | | | | | | | | FOXJ3_6 | G | 2.622625904 | ENCODE |

| | | | | | | | | | | | | |
|---|---|---|---|---|---|---|---|---|---|---|---|---|
| | | | | | | | | | SOX18_2 | A | 2.510067804 | ENCODE |
| | | | | | | | | | SOX21_2 | A | 2.455707495 | ENCODE |
| | | | | | | | | | SOX3_1 | A | 2.44392421 | ENCODE |
| | | | | | | | | | SOX10_8 | A | 2.372716917 | ENCODE |
| | | | | | | | | | SOX4_2 | A | 2.349837978 | ENCODE |
| | | | | | | | | | FOXJ2_3 | G | 2.330394372 | ENCODE |
| | | | | | | | | | SOX10_3 | A | 2.324207557 | ENCODE |
| | | | | | | | | | SOX2_2 | A | 2.317605595 | ENCODE |
| | | | | | | | | | SOX8_4 | A | 2.302585093 | ENCODE |
| | | | | | | | | | SOX7_3 | A | 2.302585093 | ENCODE |
| | | | | | | | | | SOX8_7 | A | 2.272345208 | ENCODE |
| | | | | | | | | | SOX9_7 | A | 2.225081532 | ENCODE |
| | | | | | | | | | Sox11 | A | 2.196891854 | JASPAR |
| | | | | | | | | | SOX11_2 | A | 2.195191761 | ENCODE |
| | | | | | | | | | ZNF136 | A | 2.140857844 | JASPAR |
| | | | | | | | | | SRY_5 | A | 2.119535999 | ENCODE |
| | | | | | | | | | SOX15_2 | A | 2.100241416 | ENCODE |
| 8 | rs2104598 | G | A | G | 0.805454 | 0.78646 | | PAS islet | | | | |
| | | | | | | | | PAS HepG2 | HOXA13_4 | A | 3.220457687 | ENCODE |
| | | | | | | | | | NKX6-3 | A | 2.531218137 | JASPAR |
| | | | | | | | | | NFATC1_1 | G | 2.072509105 | ENCODE |
| 9 | rs79612474 | C | T | C | 0.63966 | 0.616856 | PAS islet | PAS islet | | | | |
| | | | | | | | | DAS K562 | PITX2_2 | C | 3.810769272 | ENCODE |
| | | | | | | | | DAS HepG2 | PITX1_3 | C | 3.337827768 | ENCODE |
| | | | | | | | | | Mecom | C | 3.325248486 | JASPAR |
| | | | | | | | | | Otx2(Homeobox) | C | 3.268637334 | HOMER |
| | | | | | | | | | RUNX1_8 | C | 3.251411615 | ENCODE |
| | | | | | | | | | DOBOX5_1 | C | 3.248302826 | ENCODE |
| | | | | | | | | | OTX2_3 | C | 3.181834553 | ENCODE |
| | | | | | | | | | POU2F2_known9 | T | 3.091042453 | ENCODE |
| | | | | | | | | | FOXP3_1 | T | 2.643212682 | ENCODE |
| | | | | | | | | | GSC_1 | C | 2.48365743 | ENCODE |
| | | | | | | | | | GSC(Homeobox) | C | 2.292174357 | HOMER |
| | | | | | | | | | FOXD3_1 | T | 2.170815815 | ENCODE |
| | | | | | | | | | SIX5_disc3 | T | 2.062791529 | ENCODE |
| | | | | | | | | | FOXD3_2 | T | 2.008878906 | ENCODE |
| 10 | rs11199755 | C | T | C | 0.185268 | 0.181022 | | PAS islet | | | | |
| | | | | | | | | | KLF3(Zf) | C | 3.246373074 | HOMER |
| | | | | | | | | | Sp2(Zf) | C | 2.75565966 | HOMER |
| | | | | | | | | | EGR1_known12 | C | 2.525059524 | ENCODE |
| | | | | | | | | | RREB1_2 | C | 2.492938821 | ENCODE |
| | | | | | | | | | RREB1 | C | 2.489732279 | JASPAR |
| | | | | | | | | | SP2 | C | 2.455789251 | JASPAR |
| | | | | | | | | | CCNT2_disc2 | C | 2.365042448 | ENCODE |
| | | | | | | | | | RREB1_1 | C | 2.339323919 | ENCODE |
| 11 | rs4930011 | C | G | C | 0.156832 | 0.124978 | DAS islet | PAS HepG2 | | | | |

| | | | | | | | | | | | | |
|---|---|---|---|---|---|---|---|---|---|---|---|---|
| | | | | | | | | DAS islet | EGR1_disc7 | C | 4.028531344 | ENCODE |
| | | | | | | | | | ZSCAN4 | C | 3.803162311 | JASPAR |
| | | | | | | | | | PBX3_disc3 | G | 3.250943961 | ENCODE |
| | | | | | | | | | EGR1 | C | 3.026358931 | JASPAR |
| | | | | | | | | | KLF9 | C | 2.947315174 | JASPAR |
| | | | | | | | | | EGR1_known12 | C | 2.873433242 | ENCODE |
| | | | | | | | | | EGR1_known9 | C | 2.81979424 | ENCODE |
| | | | | | | | | | ZSCAN4_3 | C | 2.817213003 | ENCODE |
| | | | | | | | | | EGR3_3 | C | 2.780620894 | ENCODE |
| | | | | | | | | | EGR3_1 | C | 2.7120445 | ENCODE |
| | | | | | | | | | KLF9 | C | 2.590267165 | JASPAR |
| | | | | | | | | | RREB1 | C | 2.547940028 | JASPAR |
| | | | | | | | | | RREB1_2 | C | 2.541477001 | ENCODE |
| | | | | | | | | | EGR3_2 | C | 2.479482059 | ENCODE |
| | | | | | | | | | EGR4_2 | C | 2.47940911 | ENCODE |
| | | | | | | | | | EGR4 | C | 2.459888887 | JASPAR |
| | | | | | | | | | FOXA1:AR(Forkhead,NR) | G | 2.364698918 | HOMER |
| | | | | | | | | | RREB1 | C | 2.254323635 | JASPAR |
| | | | | | | | | | RREB1_2 | C | 2.249265455 | ENCODE |
| 12 | rs10743026 | C | G | G | 0.386128 | 0.456547 | DAS islet | DAS islet | | | | |
| | | | | | | | DAS K562 | DAS K562 | FOXG1_1 | G | 1.976288184 | ENCODE |
| 13 | rs4758317 | C | A | C | 0.413518 | 0.381916 | DAS HepG2 | DAS HepG2 | | | | |
| | | | | | | | | | OSR2 | C | 5.370790015 | JASPAR |
| | | | | | | | | | SPDEF_4 | A | 2.157954807 | ENCODE |
| 14 | rs78959242 | T | C | C | 0.02426 | 0.024458 | | PAS islet | | | | |
| | | | | | | | | PAS HepG2 | GATA1 | T | 5.60199978 | JASPAR |
| 15 | rs75336838 | C | T | T | 0.13511 | 0.144061 | PAS HepG2 | PAS HepG2 | | | | |
| | | | | | | | DAS islet | DAS islet | NFAT(RHD) | T | 4.860563982 | HOMER |
| | | | | | | | | DAS K562 | ZNF75D | C | 3.734136046 | JASPAR |
| | | | | | | | | | SIX5_disc2 | C | 3.698448905 | ENCODE |
| | | | | | | | | | ZNF143_disc1 | C | 3.569078254 | ENCODE |
| | | | | | | | | | RBPJ_1 | C | 3.215123475 | ENCODE |
| | | | | | | | | | Sox11 | T | 2.839302131 | JASPAR |
| | | | | | | | | | SOX18_2 | T | 2.834275855 | ENCODE |
| | | | | | | | | | NFAT5 | T | 2.741135926 | JASPAR |
| | | | | | | | | | SOX10_8 | T | 2.699206176 | ENCODE |
| | | | | | | | | | SOX7_3 | T | 2.676595249 | ENCODE |
| | | | | | | | | | SOX2_2 | T | 2.586226756 | ENCODE |
| | | | | | | | | | SOX21_2 | T | 2.577251797 | ENCODE |
| | | | | | | | | | SOX11_2 | T | 2.571497063 | ENCODE |
| | | | | | | | | | SOX21 | T | 2.530741186 | JASPAR |
| | | | | | | | | | SRY_5 | T | 2.508437147 | ENCODE |
| | | | | | | | | | SOX8_4 | T | 2.497424868 | ENCODE |
| | | | | | | | | | GFY(?) | C | 2.486277452 | HOMER |
| | | | | | | | | | SOX9_7 | T | 2.407255716 | ENCODE |
| | | | | | | | | | SOX17_4 | T | 2.401256621 | ENCODE |

| | | | | | | | | | | | | |
|---|---|---|---|---|---|---|---|---|---|---|---|---|
| | | | | | | | | | RBPJ_2 | C | 2.381458073 | ENCODE |
| | | | | | | | | | SOX3_1 | T | 2.363209715 | ENCODE |
| | | | | | | | | | SOX4_2 | T | 2.160614832 | ENCODE |
| | | | | | | | | | NFAT:AP1(RHD,bZIP) | T | 2.156733216 | HOMER |
| | | | | | | | | | THAP11 | C | 2.137070655 | JASPAR |
| | | | | | | | | | SOX10_3 | T | 2.11971991 | ENCODE |
| | | | | | | | | | SOX8_7 | T | 2.10650472 | ENCODE |
| | | | | | | | | | SRY_5 | T | 2.095210034 | ENCODE |
| | | | | | | | | | SOX2_2 | T | 2.029291758 | ENCODE |
| 16 | rs148893083 | C | T | C | 0.016996 | 0.016369 | DAS HepG2 | DAS HepG2 | | | | |
| | | | | | | | | | HNF4a(NR),DR1 | C | 2.277325426 | HOMER |
| 17 | rs7933438 | G | A | G | 0.639608 | 0.467173 | PAS islet | PAS islet | | | | |
| | | | | | | | PAS K562 | PAS K562 | GATA_disc2 | G | 2.816530844 | ENCODE |
| | | | | | | | PAS HepG2 | PAS HepG2 | Fos(bZIP) | G | 2.092289684 | HOMER |
| 18 | 12:4288000:T:C | T | C | C | 0.094517 | 0.098531 | | PAS K562 | | | | |
| | | | | | | | | | ZKSCAN3_1 | C | 2.254693805 | ENCODE |
| 19 | rs10466905 | G | A | G | 0.036248 | 0.011166 | PAS islet | PAS islet | | | | |
| | | | | | | | PAS K562 | PAS K562 | DUX4 | A | 5.801974501 | JASPAR |
| | | | | | | | PAS HepG2 | PAS HepG2 | AP1_known1 | G | 5.020181906 | ENCODE |
| | | | | | | | | | AP1_known4 | G | 4.652335654 | ENCODE |
| | | | | | | | | | DUXA | A | 4.531350274 | JASPAR |
| | | | | | | | | | DUXA_1 | A | 4.440470231 | ENCODE |
| | | | | | | | | | DUX4(Homeobox) | A | 4.251664817 | HOMER |
| | | | | | | | | | AP1_known2 | G | 4.251187034 | ENCODE |
| | | | | | | | | | AP1_known3 | G | 3.718860286 | ENCODE |
| | | | | | | | | | Duxbl(Homeobox) | A | 3.322416504 | HOMER |
| | | | | | | | | | POU6F2_2 | A | 3.143652404 | ENCODE |
| | | | | | | | | | GATA_disc6 | G | 3.111493064 | ENCODE |
| | | | | | | | | | PHOX2B_2 | A | 2.822506282 | ENCODE |
| | | | | | | | | | POU6F2_2 | A | 2.792133318 | ENCODE |
| | | | | | | | | | ALX4_3 | A | 2.112372219 | ENCODE |
| | | | | | | | | | ALX4_4 | A | 2.047692843 | ENCODE |
| 20 | rs117797076 | G | A | G | 0.036711 | 0.036139 | | DAS HepG2 | | | | |
| | | | | | | | | | T_2 | A | 3.21237526 | ENCODE |
| | | | | | | | | | Tbx6(T-box) | A | 2.75605942 | HOMER |
| | | | | | | | | | CTCF_disc6 | A | 2.620420204 | ENCODE |
| | | | | | | | | | Tbr1(T-box) | A | 2.320143405 | HOMER |
| | | | | | | | | | TBR1_1 | A | 2.00847213 | ENCODE |
| 21 | rs140895856 | C | T | T | 0.081348 | 0.083881 | PAS islet | PAS islet | | | | |
| | | | | | | | | PAS K562 | RHOXF1_2 | T | 3.824351967 | ENCODE |
| | | | | | | | | PAS HepG2 | RHOXF1_1 | T | 3.501563238 | ENCODE |
| | | | | | | | | | CHD2_disc2 | T | 3.088776591 | ENCODE |
| | | | | | | | | | E2F_disc5 | T | 3.065831034 | ENCODE |
| 22 | rs4760278 | C | A | C | 0.123359 | 0.102314 | PAS islet | PAS islet | | | | |
| | | | | | | | PAS K562 | PAS K562 | ATF3_known3 | C | 2.417075403 | ENCODE |
| | | | | | | | PAS HepG2 | PAS HepG2 | SOX21_5 | A | 2.353522029 | ENCODE |

| | | | | | | | | | | | | |
|---|---|---|---|---|---|---|---|---|---|---|---|---|
| | | | | | | | | | SOX10_10 | A | 2.037250319 | ENCODE |
| 23 | rs2268501 | C | T | C | 0.815791 | 0.805267 | DAS HepG2 | DAS islet | | | | |
| | | | | | | | | DAS HepG2 | HIF2a(bHLH) | T | 3.013505873 | HOMER |
| | | | | | | | | | NFAT:AP1(RHD,bZIP) | C | 2.134998196 | HOMER |
| 24 | rs1337939 | T | C | T | 0.480531 | 0.476055 | | | | | | |
| | | | | | | | | | IRF_known18 | C | 2.011388078 | ENCODE |
| 25 | rs12429545 | G | A | A | 0.352101 | 0.372017 | | PAS islet | | | | |
| | | | | | | | | DAS K562 | MYC_disc6 | G | 3.139323193 | ENCODE |
| 26 | rs76533333 | A | G | G | 0.094495 | 0.108793 | | DAS islet | | | | |
| | | | | | | | | | HNF4_known24 | G | 2.744417845 | ENCODE |
| | | | | | | | | | NKX2-1_1 | G | 2.717018871 | ENCODE |
| | | | | | | | | | NR2F6_1 | G | 2.516159193 | ENCODE |
| | | | | | | | | | SCRT1 | G | 2.481508176 | JASPAR |
| | | | | | | | | | NR2F6_4 | G | 2.375655661 | ENCODE |
| | | | | | | | | | VDR_1 | G | 2.259765096 | ENCODE |
| 27 | rs142329603 | T | C | C | 0.444308 | 0.445407 | | PAS K562 | | | | |
| | | | | | | | | | ZNF341 | T | 5.191345247 | JASPAR |
| | | | | | | | | | ZNF341(Zf) | T | 4.234796398 | HOMER |
| | | | | | | | | | ZNF189(Zf) | T | 3.772879429 | HOMER |
| | | | | | | | | | Znf263(Zf) | C | 3.525672293 | HOMER |
| | | | | | | | | | GRE(NR),IR3 | T | 3.154337304 | HOMER |
| | | | | | | | | | GRE(NR),IR3 | T | 3.111436306 | HOMER |
| | | | | | | | | | PAX4_5 | T | 2.995732274 | ENCODE |
| | | | | | | | | | NR3C1_known9 | T | 2.975461309 | ENCODE |
| | | | | | | | | | GRE(NR),IR3 | T | 2.967891648 | HOMER |
| | | | | | | | | | NR3C1_known16 | T | 2.741461887 | ENCODE |
| | | | | | | | | | ARE(NR) | T | 2.732360791 | HOMER |
| | | | | | | | | | NR3C1_disc1 | T | 2.697000365 | ENCODE |
| | | | | | | | | | Ar | T | 2.670309873 | JASPAR |
| | | | | | | | | | PR(NR) | T | 2.669516537 | HOMER |
| | | | | | | | | | NR3C1_known16 | T | 2.662977513 | ENCODE |
| | | | | | | | | | PR(NR) | T | 2.654154433 | HOMER |
| | | | | | | | | | ARE(NR) | T | 2.653636779 | HOMER |
| | | | | | | | | | Ar | T | 2.604327151 | JASPAR |
| | | | | | | | | | GRE(NR),IR3 | T | 2.42022056 | HOMER |
| | | | | | | | | | NR3C2 | T | 2.402610969 | JASPAR |
| 28 | rs7156625 | G | A | A | 0.441799 | 0.483507 | PAS islet | PAS islet | | | | |
| | | | | | | | PAS HepG2 | PAS HepG2 | SIX5_disc2 | G | 3.490603969 | ENCODE |
| | | | | | | | DAS K562 | DAS K562 | ZNF143_disc1 | G | 3.409818125 | ENCODE |
| | | | | | | | | | CTCFL_disc1 | G | 3.157423046 | ENCODE |
| | | | | | | | | | RXRA_disc2 | G | 2.709382646 | ENCODE |
| | | | | | | | | | CTCF(Zf) | G | 2.317781177 | HOMER |
| | | | | | | | | | HNF1_2 | A | 2.08133779 | ENCODE |
| 29 | rs35889227 | G | T | G | 0.255772 | 0.202556 | PAS islet | PAS islet | | | | |
| | | | | | | | PAS HepG2 | PAS HepG2 | SOX17_5 | G | 3.075069016 | ENCODE |
| | | | | | | | | DAS K562 | SOX7_4 | G | 3.056819966 | ENCODE |

| | | | | | | | | | | | | |
|---|---|---|---|---|---|---|---|---|---|---|---|---|
| | | | | | | | | | SOX10_10 | G | 2.96686429 | ENCODE |
| | | | | | | | | | SOX21_5 | G | 2.946048853 | ENCODE |
| | | | | | | | | | SOX10_6 | G | 2.907721396 | ENCODE |
| | | | | | | | | | SOX9_9 | G | 2.890371758 | ENCODE |
| | | | | | | | | | SOX1_3 | G | 2.851692903 | ENCODE |
| | | | | | | | | | SOX3_3 | G | 2.774396807 | ENCODE |
| | | | | | | | | | SOX2_3 | G | 2.72861418 | ENCODE |
| | | | | | | | | | SOX8_3 | G | 2.721987709 | ENCODE |
| | | | | | | | | | SOX18_4 | G | 2.705095989 | ENCODE |
| | | | | | | | | | SOX8_8 | G | 2.697687414 | ENCODE |
| | | | | | | | | | SOX2_7 | G | 2.679452794 | ENCODE |
| | | | | | | | | | SOX14_3 | G | 2.604587219 | ENCODE |
| | | | | | | | | | LHX6_5 | T | 2.583298496 | ENCODE |
| | | | | | | | | | LHX6_5 | T | 2.409448239 | ENCODE |
| | | | | | | | | | FOXA_known1 | T | 2.343686769 | ENCODE |
| 30 | rs73347525 | A | G | A | 0.429225 | 0.399129 | PAS islet | PAS islet | | | | |
| | | | | | | | | | ZNF415(Zf) | G | 4.65646348 | HOMER |
| | | | | | | | | | HNF4_disc4 | G | 3.359350272 | ENCODE |
| | | | | | | | | | ZBTB12 | A | 2.884779945 | JASPAR |
| 31 | rs12912777 | C | T | C | 0.293417 | 0.286795 | | DAS islet | | | | |
| | | | | | | | | | Bcl6(Zf) | T | 2.324824705 | HOMER |
| | | | | | | | | | ATF3_known5 | C | 2.141097623 | ENCODE |
| 32 | rs504348 | C | G | G | 0.121522 | 0.130112 | | PAS HepG2 | | | | |
| | | | | | | | | | ESRRA_known8 | C | 2.20255224 | ENCODE |
| 33 | rs184499898 | A | G | G | 0.005922 | 0.00598 | | | | | | |
| | | | | | | | | | RREB1 | G | 2.708050201 | JASPAR |
| | | | | | | | | | RREB1_2 | G | 2.703552136 | ENCODE |
| | | | | | | | | | RREB1_1 | G | 2.510224458 | ENCODE |
| 34 | rs7213347 | G | C | C | 0.016265 | 0.017125 | | | | | | |
| | | | | | | | | | Sox7(HMG) | C | 4.115967026 | HOMER |
| | | | | | | | | | Sox4(HMG) | C | 3.388512222 | HOMER |
| | | | | | | | | | Sox17(HMG) | C | 3.129652998 | HOMER |
| | | | | | | | | | Sox6(HMG) | C | 2.934890614 | HOMER |
| | | | | | | | | | SOX9_3 | C | 2.805484651 | ENCODE |
| | | | | | | | | | SOX8 | C | 2.770016925 | JASPAR |
| | | | | | | | | | SOX9_1 | C | 2.60359262 | ENCODE |
| | | | | | | | | | SOX9_2 | C | 2.145852101 | ENCODE |
| | | | | | | | | | SOX9 | C | 2.145852101 | JASPAR |
| 35 | rs10468467 | A | G | A | 0.003669 | 0.003184 | DAS islet | DAS islet | | | | |
| | | | | | | | DAS K562 | DAS K562 | PROX1_1 | G | 2.864134743 | ENCODE |
| | | | | | | | DAS HepG2 | DAS HepG2 | PROX1 | G | 2.823392085 | JASPAR |
| 36 | rs2313286 | C | G | G | 0.019141 | 0.026371 | PAS K562 | PAS K562 | | | | |
| | | | | | | | DAS islet | DAS islet | Usf2(bHLH) | C | 2.211969757 | HOMER |
| 37 | rs12938438 | C | G | G | 0.03663 | 0.038492 | DAS HepG2 | DAS HepG2 | | | | |
| | | | | | | | | | AMYB(HTH) | G | 5.386983004 | HOMER |
| | | | | | | | | | MYB_5 | G | 3.27512537 | ENCODE |

| | | | | | | | | | | | | |
|---|---|---|---|---|---|---|---|---|---|---|---|---|
| 38 | rs112412433 | C | T | T | 0.718152 | 0.721123 | | PAS islet | | | | |
| | | | | | | | | | TCF7_2 | C | 2.30965226 | ENCODE |
| 39 | rs12600858 | G | A | A | 0.218999 | 0.24005 | PAS HepG2 | PAS islet | | | | |
| | | | | | | | | PAS K562 | GLI2_2 | G | 2.415913778 | ENCODE |
| | | | | | | | | PAS HepG2 | RUNX2_2 | A | 2.380215254 | ENCODE |
| 40 | rs75646162 | G | A | G | 0.265144 | 0.207517 | PAS islet | PAS islet | | | | |
| | | | | | | | PAS K562 | PAS K562 | BPTF_1 | A | 3.654898446 | ENCODE |
| 41 | rs7208565 | C | T | C | 0.032429 | 0.02668 | PAS HepG2 | PAS HepG2 | | | | |
| | | | | | | | | | KLF17 | C | 3.299829401 | JASPAR |
| 42 | rs12974537 | C | T | C | 0.011863 | 0.010339 | DAS K562 | DAS islet | | | | |
| | | | | | | | | DAS K562 | Reverb(NR),DR2 | T | 3.379995174 | HOMER |
| | | | | | | | | | REST_disc9 | C | 3.028414921 | ENCODE |
| 43 | rs2974752 | G | A | G | 0.214312 | 0.188236 | PAS islet | PAS islet | | | | |
| | | | | | | | | PAS K562 | NRF1_disc1 | G | 3.300101265 | ENCODE |
| | | | | | | | | PAS HepG2 | HES2 | G | 3.17073545 | JASPAR |
| | | | | | | | | | HES2 | G | 2.988512026 | JASPAR |
| | | | | | | | | | MYC_known20 | G | 2.957991946 | ENCODE |
| | | | | | | | | | MYC_known20 | G | 2.75705778 | ENCODE |
| | | | | | | | | | HEY2_2 | G | 2.642652301 | ENCODE |
| | | | | | | | | | HEY2 | G | 2.574598837 | JASPAR |
| | | | | | | | | | EGR1_disc3 | G | 2.523296539 | ENCODE |
| | | | | | | | | | HES7 | G | 2.349976881 | JASPAR |
| | | | | | | | | | SOHLH2 | G | 2.130115833 | JASPAR |
| 44 | rs584007 | A | G | A | 0.205884 | 0.192706 | DAS K562 | PAS islet | | | | |
| | | | | | | | | PAS HepG2 | CPEB1_1 | A | 2.624126199 | ENCODE |
| | | | | | | | | DAS K562 | ZFP3(Zf) | G | 2.361425593 | HOMER |
| 45 | rs62136859 | A | G | A | 0.014113 | 0.013794 | DAS K562 | PAS islet | | | | |
| | | | | | | | DAS HepG2 | DAS K562 | TBX20_5 | A | 2.469546606 | ENCODE |
| 46 | rs34813869 | A | G | G | 0.081391 | 0.084626 | PAS K562 | DAS HepG2 | | | | |
| | | | | | | | | | TLX2_1 | A | 2.654890833 | ENCODE |
| | | | | | | | | | ZNF165(Zf) | G | 2.342237865 | HOMER |
| | | | | | | | | | VENTX_2 | A | 2.335758867 | ENCODE |
| | | | | | | | | | ZSCAN16_1 | G | 2.311228189 | ENCODE |
| | | | | | | | | | VENTX_2 | A | 2.268968482 | ENCODE |
| | | | | | | | | | EGR1_disc4 | G | 2.243808657 | ENCODE |
| | | | | | | | | | TFAP4_1 | A | 2.113067898 | ENCODE |
| 47 | rs73050880 | A | G | G | 0.001908 | 0.001952 | | | | | | |
| | | | | | | | | | RBPJ:Ebox(?,bHLH) | G | 1.511212886 | HOMER |
| 48 | rs10206462 | C | T | T | 0.07925 | 0.08571 | | PAS islet | | | | |
| | | | | | | | | | BATF_disc2 | C | 3.587696667 | ENCODE |
| | | | | | | | | | IRF_disc6 | C | 3.164196796 | ENCODE |
| | | | | | | | | | PRDM1_known1 | T | 2.682074715 | ENCODE |
| | | | | | | | | | NFKB_known6 | T | 2.582650982 | ENCODE |
| | | | | | | | | | ZNF684 | C | 2.255225623 | JASPAR |
| | | | | | | | | | NFkB-p65(RHD) | T | 2.15910337 | HOMER |
| 49 | rs112694524 | G | A | G | 0.822565 | 0.820019 | PAS islet | PAS islet | | | | |

| | | | | | | | | | | | | |
|---|---|---|---|---|---|---|---|---|---|---|---|---|
| | | | | | | | | PAS K562 | SIN3A_disc4 | A | 3.065659288 | ENCODE |
| | | | | | | | | | MYF_1 | A | 2.97134082 | ENCODE |
| | | | | | | | | | TATA_disc7 | G | 2.374990937 | ENCODE |
| | | | | | | | | | REST_known3 | A | 2.253794929 | ENCODE |
| | | | | | | | | | TFAP2_known4 | G | 2.152772815 | ENCODE |
| | | | | | | | | | HIC1_1 | G | 2.118628595 | ENCODE |
| | | | | | | | | | RAD21_disc8 | G | 2.086361985 | ENCODE |
| | | | | | | | | | TFAP4_1 | A | 2.032188657 | ENCODE |
| 50 | rs2121564 | C | G | G | 0.028745 | 0.036395 | DAS islet | DAS islet | | | | |
| | | | | | | | DAS K562 | DAS K562 | CREB3L2_2 | G | 3.925575972 | ENCODE |
| | | | | | | | DAS HepG2 | DAS HepG2 | CREB3L1_3 | G | 3.721624535 | ENCODE |
| | | | | | | | | | Plagl1 | G | 3.559028949 | JASPAR |
| | | | | | | | | | CREB3L2_2 | G | 3.435499976 | ENCODE |
| | | | | | | | | | CREB3L1_3 | G | 3.389221191 | ENCODE |
| | | | | | | | | | SP1_known3 | G | 3.013353875 | ENCODE |
| | | | | | | | | | MYC_known14 | G | 2.856470206 | ENCODE |
| | | | | | | | | | CREB3L1_4 | G | 2.732969523 | ENCODE |
| | | | | | | | | | HIC1_3 | C | 2.470644203 | ENCODE |
| | | | | | | | | | ATF3_disc3 | C | 2.065309659 | ENCODE |
| 51 | rs74646951 | C | A | C | 0.07839 | 0.05773 | PAS islet | PAS islet | | | | |
| | | | | | | | PAS K562 | PAS K562 | GATA5 | C | 4.651690202 | JASPAR |
| | | | | | | | PAS HepG2 | PAS HepG2 | GATA_disc1 | C | 4.05862648 | ENCODE |
| | | | | | | | | | Gata4(Zf) | C | 3.513224589 | HOMER |
| | | | | | | | | | HDAC2_disc1 | C | 2.630758527 | ENCODE |
| | | | | | | | | | Gata6(Zf) | C | 2.605635196 | HOMER |
| | | | | | | | | | Gata1(Zf) | C | 2.321816455 | HOMER |
| | | | | | | | | | Gata2(Zf) | C | 2.210893735 | HOMER |
| | | | | | | | | | GATA4 | C | 2.186374561 | JASPAR |
| | | | | | | | | | GATA_known15 | C | 2.18311144 | ENCODE |
| | | | | | | | | | GATA_known21 | C | 2.088099604 | ENCODE |
| | | | | | | | | | GATA_known17 | C | 2.071473372 | ENCODE |
| | | | | | | | | | GATA_known22 | C | 2.007615807 | ENCODE |
| 52 | rs478333 | G | A | G | 0.013148 | 0.012279 | DAS islet | DAS islet | | | | |
| | | | | | | | DAS K562 | DAS K562 | E2A(bHLH),near_PU.1 | G | 3.966417077 | HOMER |
| | | | | | | | DAS HepG2 | DAS HepG2 | TCF3 | G | 3.356745619 | JASPAR |
| | | | | | | | | | TCF12(var.2) | G | 3.298502646 | JASPAR |
| | | | | | | | | | Slug(Zf) | G | 3.242532021 | HOMER |
| | | | | | | | | | SNAI1 | G | 2.455092387 | JASPAR |
| | | | | | | | | | ZEB1 | G | 2.31261024 | JASPAR |
| | | | | | | | | | SNAI3 | G | 2.301348234 | JASPAR |
| 53 | rs10203361 | G | A | A | 0.03134 | 0.034098 | DAS islet | DAS islet | | | | |
| | | | | | | | DAS K562 | DAS K562 | SP1_known9 | G | 2.949716036 | ENCODE |
| | | | | | | | DAS HepG2 | DAS HepG2 | KLF17 | G | 2.710630848 | JASPAR |
| | | | | | | | | | SREBP_known2 | G | 2.644555132 | ENCODE |
| | | | | | | | | | KLF16_1 | G | 2.530800506 | ENCODE |
| | | | | | | | | | Srebp2(bHLH) | G | 2.492441669 | HOMER |

| | | | | | | | | | | | | |
|---|---|---|---|---|---|---|---|---|---|---|---|---|
| | | | | | | | | | KLF16 | G | 2.445393496 | JASPAR |
| | | | | | | | | | SP8_1 | G | 2.307099773 | ENCODE |
| | | | | | | | | | SP8 | G | 2.270099638 | JASPAR |
| | | | | | | | | | SP3 | G | 2.008077055 | JASPAR |
| 54 | rs2943654 | C | T | T | 0.194378 | 0.217248 | | | | | | |
| | | | | | | | | | HMBOX1_1 | C | 1.941571747 | ENCODE |
| 55 | rs117809958 | T | A | T | 0.05081 | 0.04835 | PAS islet | PAS islet | | | | |
| | | | | | | | | PAS HepG2 | MAF_known6 | A | 6.466508178 | ENCODE |
| | | | | | | | | | MAF_known8 | A | 5.91162166 | ENCODE |
| | | | | | | | | | Mafb | A | 4.69245297 | JASPAR |
| | | | | | | | | | MAF_known10 | A | 4.618115351 | ENCODE |
| | | | | | | | | | MafF(bZIP) | A | 4.598676657 | HOMER |
| | | | | | | | | | NRL_1 | A | 4.173632838 | ENCODE |
| | | | | | | | | | MAFG | A | 3.976167127 | JASPAR |
| | | | | | | | | | MAFF | A | 3.888855946 | JASPAR |
| | | | | | | | | | Tbx6(T-box) | A | 3.786202832 | HOMER |
| | | | | | | | | | TBX6 | A | 3.556720745 | JASPAR |
| | | | | | | | | | TBX3 | A | 3.498615044 | JASPAR |
| | | | | | | | | | MAF_known4 | A | 3.299637182 | ENCODE |
| | | | | | | | | | MAF_known5 | A | 3.150419866 | ENCODE |
| | | | | | | | | | MafA(bZIP) | A | 3.031420003 | HOMER |
| | | | | | | | | | SIX5_known5 | A | 2.730858897 | ENCODE |
| | | | | | | | | | TBX18 | A | 2.723128092 | JASPAR |
| | | | | | | | | | TBX4_1 | A | 2.637845942 | ENCODE |
| | | | | | | | | | TBX4 | A | 2.637845942 | JASPAR |
| | | | | | | | | | Tbx5(T-box) | A | 2.368398312 | HOMER |
| | | | | | | | | | MAFF_1 | A | 2.196886797 | ENCODE |
| | | | | | | | | | TBX5_4 | A | 2.056402751 | ENCODE |
| | | | | | | | | | TBX5 | A | 2.056402751 | JASPAR |
| 56 | rs10929270 | T | A | T | 0.015716 | 0.013546 | DAS K562 | PAS islet | | | | |
| | | | | | | | DAS HepG2 | DAS K562 | ESR1 | T | 3.390225762 | JASPAR |
| | | | | | | | | DAS HepG2 | ETS:RUNX(ETS,Runt) | A | 2.8963598 | HOMER |
| | | | | | | | | | ESRRA_known6 | T | 2.754570217 | ENCODE |
| | | | | | | | | | STAT_disc7 | A | 2.616045056 | ENCODE |
| | | | | | | | | | ESRRA_known6 | T | 2.412210965 | ENCODE |
| | | | | | | | | | ZNF460 | T | 2.240179527 | JASPAR |
| | | | | | | | | | TATA_disc8 | T | 2.114706563 | ENCODE |
| 57 | rs13042148 | C | T | C | 0.124361 | 0.08552 | PAS islet | PAS islet | | | | |
| | | | | | | | PAS K562 | PAS K562 | SP1_known8 | C | 6.758909651 | ENCODE |
| | | | | | | | PAS HepG2 | PAS HepG2 | SP9 | C | 6.502290171 | JASPAR |
| | | | | | | | | | KLF14 | C | 6.06828664 | JASPAR |
| | | | | | | | | | SP4 | C | 5.984591668 | JASPAR |
| | | | | | | | | | KLF14_1 | C | 5.529098707 | ENCODE |
| | | | | | | | | | KLF11 | C | 5.476836637 | JASPAR |
| | | | | | | | | | SP3 | C | 5.368698163 | JASPAR |
| | | | | | | | | | KLF7_1 | C | 5.302069716 | ENCODE |

| | | | |
|---|---|---|---|
| SP4_1 | C | 5.271565181 | ENCODE |
| KLF13 | C | 5.168724538 | JASPAR |
| SP8 | C | 4.980329945 | JASPAR |
| SP8_1 | C | 4.938439003 | ENCODE |
| KLF16 | C | 4.875659858 | JASPAR |
| KLF13_1 | C | 4.851005149 | ENCODE |
| KLF16_1 | C | 4.774358743 | ENCODE |
| SP1_known9 | C | 4.714696585 | ENCODE |
| KLF2 | C | 4.682589785 | JASPAR |
| KLF6 | C | 4.510791249 | JASPAR |
| KLF10 | C | 4.352404379 | JASPAR |
| HNF4_known6 | T | 4.211291827 | ENCODE |
| KLF9 | C | 4.077781873 | JASPAR |
| SP1 | C | 4.040855877 | JASPAR |
| SP4_2 | C | 4.016226783 | ENCODE |
| KLF1(Zf) | C | 3.950174771 | HOMER |
| Klf9(Zf) | C | 3.912023005 | HOMER |
| KLF14(Zf) | C | 3.781512256 | HOMER |
| KLF4_1 | C | 3.735305551 | ENCODE |
| Sp5(Zf) | C | 3.708073901 | HOMER |
| KLF10(Zf) | C | 3.686307657 | HOMER |
| SP1_known2 | C | 3.515342727 | ENCODE |
| KLF12_2 | C | 3.477011753 | ENCODE |
| Klf12 | C | 3.409744442 | JASPAR |
| KLF3(Zf) | C | 3.401566453 | HOMER |
| SP1_known6 | C | 3.390919799 | ENCODE |
| KLF6(Zf) | C | 3.3437798 | HOMER |
| Sp1(Zf) | C | 3.293711467 | HOMER |
| Klf4(Zf) | C | 3.280360909 | HOMER |
| KLF5(Zf) | C | 3.267450495 | HOMER |
| SP1_known5 | C | 3.175783682 | ENCODE |
| KLF17 | C | 3.152966996 | JASPAR |
| SP1_known4 | C | 3.018004909 | ENCODE |
| IRF_disc4 | C | 2.916144836 | ENCODE |
| KLF3 | C | 2.86277444 | JASPAR |
| SP2_disc3 | C | 2.788280052 | ENCODE |
| Sp2(Zf) | C | 2.776417699 | HOMER |
| HIC1_4 | T | 2.553899521 | ENCODE |
| RXRB_2 | T | 2.490763182 | ENCODE |
| PAX9_1 | T | 2.473928173 | ENCODE |
| PAX5(Paired,Homeobox) | T | 2.451823615 | HOMER |
| RXRA_known14 | T | 2.346617363 | ENCODE |
| RXRG_1 | T | 2.33408376 | ENCODE |
| Rxra | T | 2.3067692 | JASPAR |
| RXRA_known12 | T | 2.281751006 | ENCODE |
| PAX9 | T | 2.243161673 | JASPAR |

| | | | | | | | | | | | | |
|---|---|---|---|---|---|---|---|---|---|---|---|---|
| | | | | | | | | | RXRB_1 | T | 2.217085784 | ENCODE |
| | | | | | | | | | NR2F6_3 | T | 2.215336327 | ENCODE |
| | | | | | | | | | Nr2f6 | T | 2.205421345 | JASPAR |
| | | | | | | | | | TR4(NR),DR1 | T | 2.188268265 | HOMER |
| | | | | | | | | | RXRG | T | 2.183039942 | JASPAR |
| | | | | | | | | | RXRA_known10 | T | 2.170866937 | ENCODE |
| | | | | | | | | | HNF4_known19 | T | 2.145447107 | ENCODE |
| | | | | | | | | | PPARA_1 | T | 2.135763783 | ENCODE |
| | | | | | | | | | NR2F6_2 | T | 2.129832987 | ENCODE |
| | | | | | | | | | Egr2(Zf) | C | 2.073355096 | HOMER |
| | | | | | | | | | RXRB | T | 2.045274118 | JASPAR |
| | | | | | | | | | HNF4_known23 | T | 2.039642627 | ENCODE |
| | | | | | | | | | PPARD | T | 2.001380995 | JASPAR |
| 58 | rs12185776 | C | G | C | 0.069386 | 0.067508 | DAS K562 | DAS K562 | | | | |
| | | | | | | | | | NKX3-1_4 | G | 2.330684004 | ENCODE |
| 59 | rs4518111 | A | C | C | 0.050576 | 0.051441 | | PAS islet | | | | |
| | | | | | | | | DAS HepG2 | ZNF384 | A | 5.981414211 | JASPAR |
| | | | | | | | | | ZNF384 | A | 5.611667186 | JASPAR |
| | | | | | | | | | HDAC2_disc6 | A | 5.515454894 | ENCODE |
| | | | | | | | | | HDAC2_disc6 | A | 5.176804814 | ENCODE |
| | | | | | | | | | HDAC2_disc6 | A | 5.118227954 | ENCODE |
| | | | | | | | | | ZNF35_1 | A | 4.823651724 | ENCODE |
| | | | | | | | | | HDAC2_disc6 | A | 4.297098158 | ENCODE |
| | | | | | | | | | FOXJ3_8 | A | 3.432895757 | ENCODE |
| | | | | | | | | | HDAC2_disc6 | A | 3.328330512 | ENCODE |
| | | | | | | | | | STAT1::STAT2 | C | 3.174350746 | JASPAR |
| | | | | | | | | | Foxd3 | C | 3.045535096 | JASPAR |
| | | | | | | | | | FOXD3_2 | C | 3.041669369 | ENCODE |
| | | | | | | | | | FOXD3_1 | C | 3.018873802 | ENCODE |
| | | | | | | | | | FoxD3(forkhead) | C | 2.736481868 | HOMER |
| | | | | | | | | | FOXJ3_3 | A | 2.722578302 | ENCODE |
| | | | | | | | | | ZNF384 | A | 2.647678073 | JASPAR |
| | | | | | | | | | FOXJ3_8 | A | 2.515605637 | ENCODE |
| | | | | | | | | | FOXJ3_4 | C | 2.5060676 | ENCODE |
| | | | | | | | | | HDAC2_disc6 | A | 2.387955464 | ENCODE |
| | | | | | | | | | FOXJ2_3 | A | 2.308691982 | ENCODE |
| | | | | | | | | | PAX5_known1 | C | 2.263307317 | ENCODE |
| | | | | | | | | | FOXJ3_6 | A | 2.248682875 | ENCODE |
| | | | | | | | | | FOXP1_1 | A | 2.138135661 | ENCODE |
| 60 | rs17012829 | C | T | T | 0.149219 | 0.163974 | | DAS islet | | | | |
| | | | | | | | | DAS K562 | NR3C1_known6 | C | 2.785059846 | ENCODE |
| | | | | | | | | DAS HepG2 | SIX5_disc3 | T | 2.009482953 | ENCODE |
| 61 | rs114424909 | C | G | G | 0.123447 | 0.143895 | DAS islet | DAS islet | | | | |
| | | | | | | | DAS HepG2 | DAS HepG2 | T1ISRE(IRF) | G | 3.317315898 | HOMER |
| | | | | | | | | | IRF7 | G | 2.903864438 | JASPAR |
| | | | | | | | | | IRF_known17 | G | 2.828158573 | ENCODE |

| | | | | | | | | | | | | |
|---|---|---|---|---|---|---|---|---|---|---|---|---|
| | | | | | | | | | IRF_known1 | G | 2.757840866 | ENCODE |
| | | | | | | | | | Sox3 | G | 2.342858992 | JASPAR |
| 62 | rs75088635 | A | G | G | 0.159955 | 0.179606 | | DAS islet | | | | |
| | | | | | | | | DAS K562 | GCM1_2 | A | 2.356887249 | ENCODE |
| 63 | rs6785040 | T | C | C | 0.409713 | 0.42633 | DAS K562 | DAS HepG2 | | | | |
| | | | | | | | | | ZNF143_disc4 | C | 4.426528486 | ENCODE |
| | | | | | | | | | AP1_disc10 | C | 4.154163057 | ENCODE |
| | | | | | | | | | REST_disc5 | C | 3.988624399 | ENCODE |
| | | | | | | | | | YY2 | C | 3.502755611 | JASPAR |
| | | | | | | | | | BCL_disc9 | C | 3.423358953 | ENCODE |
| | | | | | | | | | YY1_disc5 | C | 2.893341122 | ENCODE |
| | | | | | | | | | YY1(Zf) | T | 2.758006299 | HOMER |
| | | | | | | | | | NR3C1_disc4 | T | 2.467664843 | ENCODE |
| | | | | | | | | | EGR1_disc6 | C | 2.178766801 | ENCODE |
| | | | | | | | | | YY2_2 | C | 2.081278089 | ENCODE |
| | | | | | | | | | E2F_disc8 | C | 2.054804237 | ENCODE |
| | | | | | | | | | E2F_disc7 | C | 2.039595633 | ENCODE |
| | | | | | | | | | ZFP42 | T | 2.031032402 | JASPAR |
| 64 | rs13080180 | C | T | C | 0.317879 | 0.272845 | PAS islet | PAS islet | | | | |
| | | | | | | | PAS K562 | PAS K562 | FOXA_known2 | T | 3.039407337 | ENCODE |
| | | | | | | | PAS HepG2 | PAS HepG2 | FOXC1_1 | C | 2.968562368 | ENCODE |
| | | | | | | | | | FOXJ3_6 | T | 2.410381663 | ENCODE |
| | | | | | | | | | FOXA_known3 | T | 2.31833345 | ENCODE |
| | | | | | | | | | FOXJ3_8 | T | 2.273764654 | ENCODE |
| | | | | | | | | | NFATC2_1 | T | 2.169233832 | ENCODE |
| | | | | | | | | | NFATC2 | T | 2.169233832 | JASPAR |
| 65 | rs1416218 | C | G | G | 0.187612 | 0.218471 | | | | | | |
| | | | | | | | | | GLIS3(Zf) | C | 4.035155691 | HOMER |
| | | | | | | | | | Zfp809(Zf) | G | 3.647155611 | HOMER |
| | | | | | | | | | p53(p53) | G | 3.516772491 | HOMER |
| | | | | | | | | | ZNF528 | C | 3.437565026 | JASPAR |
| | | | | | | | | | ZNF460 | C | 2.550087088 | JASPAR |
| | | | | | | | | | ZNF768(Zf) | C | 2.442004628 | HOMER |
| | | | | | | | | | TFCP2L1_1 | G | 2.327156354 | ENCODE |
| | | | | | | | | | Zfp809(Zf) | G | 2.176726056 | HOMER |
| 66 | rs649961 | T | C | C | 0.212595 | 0.28138 | | PAS islet | | | | |
| | | | | | | | | PAS K562 | HNF4_disc3 | T | 3.069989044 | ENCODE |
| | | | | | | | | | TCF21_1 | C | 2.949310437 | ENCODE |
| 67 | rs11539489 | C | G | G | 0.38198 | 0.388803 | | DAS K562 | | | | |
| | | | | | | | | | Sp2(Zf) | G | 2.376693065 | HOMER |
| 68 | rs1905505 | G | A | A | 0.058743 | 0.070076 | DAS islet | DAS islet | | | | |
| | | | | | | | | DAS HepG2 | TFCP2_1 | G | 2.94284281 | ENCODE |
| 69 | rs1981767 | G | A | G | 0.306473 | 0.288754 | PAS islet | PAS islet | | | | |
| | | | | | | | | PAS K562 | YY1_known3 | G | 2.675152498 | ENCODE |
| 70 | rs6780016 | C | T | C | 0.02834 | 0.026677 | PAS K562 | PAS HepG2 | | | | |
| | | | | | | | | | KLF17 | T | 3.047502294 | JASPAR |

| | | | | | | | | | | | |
|---|---|---|---|---|---|---|---|---|---|---|---|
| | | | | | | | | FOXO4_5 | T | 2.505784235 | ENCODE |
| | | | | | | | | HIC1_1 | C | 2.360572351 | ENCODE |
| 71 | rs4234731 | A | G | A | 0.383202 | 0.359077 | DAS islet | DAS islet | | | |
| | | | | | | | DAS HepG2 | DAS K562 | GLIS3(Zf) | A | 2.211251815 | HOMER |
| | | | | | | | | DAS HepG2 | CTCF-SatelliteElement(Zf?) | A | 2.151624991 | HOMER |
| | | | | | | | | | GLIS2_1 | A | 2.039363318 | ENCODE |
| 72 | rs45499402 | G | C | G | 0.212494 | 0.100503 | PAS islet | PAS islet | | | |
| | | | | | | | PAS K562 | PAS K562 | ZNF281_1 | G | 5.411986535 | ENCODE |
| | | | | | | | PAS HepG2 | PAS HepG2 | SP1_disc3 | G | 5.045183102 | ENCODE |
| | | | | | | | | | ZNF740_1 | G | 4.881527109 | ENCODE |
| | | | | | | | | | EGR1_known3 | G | 4.837124367 | ENCODE |
| | | | | | | | | | ZNF281_1 | G | 4.733003557 | ENCODE |
| | | | | | | | | | CCNT2_disc2 | G | 4.670309488 | ENCODE |
| | | | | | | | | | SP2 | G | 3.811257558 | JASPAR |
| | | | | | | | | | ZNF219_1 | G | 3.700713912 | ENCODE |
| | | | | | | | | | GTF2I_1 | G | 3.658941366 | ENCODE |
| | | | | | | | | | KLF16 | G | 3.654908097 | JASPAR |
| | | | | | | | | | KLF16_1 | G | 3.621908828 | ENCODE |
| | | | | | | | | | SPZ1_1 | G | 3.571562728 | ENCODE |
| | | | | | | | | | HF1H3B_1 | G | 3.344524995 | ENCODE |
| | | | | | | | | | ZNF740_4 | G | 3.311585223 | ENCODE |
| | | | | | | | | | CACBP_1 | G | 3.284180923 | ENCODE |
| | | | | | | | | | Zfp281(Zf) | G | 3.277038129 | HOMER |
| | | | | | | | | | ZNF740_2 | G | 3.228064251 | ENCODE |
| | | | | | | | | | KLF14(Zf) | G | 3.22444687 | HOMER |
| | | | | | | | | | GLIS1 | G | 3.138194955 | JASPAR |
| | | | | | | | | | ZNF740 | G | 3.003279479 | JASPAR |
| | | | | | | | | | GLIS3_1 | G | 2.977040141 | ENCODE |
| | | | | | | | | | Zfp281(Zf) | G | 2.961902409 | HOMER |
| | | | | | | | | | SP4_1 | G | 2.936643357 | ENCODE |
| | | | | | | | | | ZNF740 | G | 2.917828604 | JASPAR |
| | | | | | | | | | WT1_1 | G | 2.737109128 | ENCODE |
| | | | | | | | | | HF1H3B_1 | G | 2.688863717 | ENCODE |
| | | | | | | | | | ZNF143_disc3 | G | 2.621160191 | ENCODE |
| | | | | | | | | | SP1_known7 | G | 2.607406675 | ENCODE |
| | | | | | | | | | Klf9(Zf) | G | 2.538365228 | HOMER |
| | | | | | | | | | GLIS1_1 | G | 2.512989619 | ENCODE |
| | | | | | | | | | SP4_2 | G | 2.489171049 | ENCODE |
| | | | | | | | | | SP1_known8 | G | 2.467977588 | ENCODE |
| | | | | | | | | | MZF1_2 | G | 2.457906048 | ENCODE |
| | | | | | | | | | Znf281 | G | 2.438508259 | JASPAR |
| | | | | | | | | | RREB1_1 | G | 2.436864709 | ENCODE |
| | | | | | | | | | KLF14_1 | G | 2.43299311 | ENCODE |
| | | | | | | | | | SP9 | G | 2.267630078 | JASPAR |
| | | | | | | | | | KLF14 | G | 2.241271908 | JASPAR |
| | | | | | | | | | SP4 | G | 2.235574383 | JASPAR |

| | | | | | | | | | | | | |
|---|---|---|---|---|---|---|---|---|---|---|---|---|
| | | | | | | | | | SP3 | G | 2.13470422 | JASPAR |
| | | | | | | | | | SP1_known9 | G | 2.068970242 | ENCODE |
| | | | | | | | | | SP8_1 | G | 2.065130027 | ENCODE |
| | | | | | | | | | ZNF281_1 | G | 2.049749752 | ENCODE |
| | | | | | | | | | SP8 | G | 2.042349999 | JASPAR |
| | | | | | | | | | KLF11 | G | 2.034857635 | JASPAR |
| | | | | | | | | | ZNF740_3 | G | 2.029387953 | ENCODE |
| | | | | | | | | | ZNF740_1 | G | 2.022795721 | ENCODE |
| 73 | rs11729069 | C | G | G | 0.192851 | 0.200973 | | DAS K562 | | | | |
| | | | | | | | | | RARG_8 | G | 2.30982276 | ENCODE |
| 74 | rs35585881 | G | A | A | 0.174187 | 0.180135 | | | | | | |
| | | | | | | | | | TGIF1_2 | A | 1.312706004 | ENCODE |
| 75 | rs7732130 | G | A | G | 0.444827 | 0.416061 | PAS islet | PAS islet | | | | |
| | | | | | | | DAS HepG2 | DAS K562 | ZNF143 | G | 3.817234429 | JASPAR |
| | | | | | | | | DAS HepG2 | ZNF143_known1 | G | 3.215915783 | ENCODE |
| | | | | | | | | | RFX7_1 | G | 2.860913051 | ENCODE |
| | | | | | | | | | ZNF143_known2 | G | 2.8479427 | ENCODE |
| 76 | rs2546197 | T | G | G | 0.212669 | 0.234596 | | DAS islet | | | | |
| | | | | | | | | DAS HepG2 | PAX1_1 | G | 1.487728259 | ENCODE |
| 77 | rs12657266 | C | T | C | 0.083453 | 0.076229 | PAS HepG2 | PAS islet | | | | |
| | | | | | | | | PAS HepG2 | MYOD1_1 | T | 3.049106734 | ENCODE |
| | | | | | | | | | BHLHE40_disc1 | T | 2.786011743 | ENCODE |
| | | | | | | | | | SIRT6_disc1 | T | 2.363769782 | ENCODE |
| | | | | | | | | | TCF4 | T | 2.149455342 | JASPAR |
| | | | | | | | | | TFAP2C(var.2) | C | 2.13905145 | JASPAR |
| | | | | | | | | | TFAP2A | C | 2.122096717 | JASPAR |
| 78 | rs10305514 | G | T | G | 0.155389 | 0.139972 | DAS K562 | DAS K562 | | | | |
| | | | | | | | DAS HepG2 | DAS HepG2 | ZNF684 | G | 2.505322706 | JASPAR |
| 79 | rs34298980 | T | C | C | 0.230408 | 0.23588 | DAS islet | DAS islet | | | | |
| | | | | | | | DAS K562 | DAS K562 | SMAD4_1 | T | 2.567738052 | ENCODE |
| | | | | | | | DAS HepG2 | DAS HepG2 | HINFP_3 | C | 2.051944292 | ENCODE |
| 80 | rs3798519 | A | C | C | 0.044694 | 0.051619 | | DAS islet | | | | |
| | | | | | | | | DAS HepG2 | RARA::RXRA | C | 3.784446406 | JASPAR |
| | | | | | | | | | RXRA_known8 | C | 3.525374169 | ENCODE |
| 81 | rs72939920 | A | T | A | 0.003721 | 0.003431 | PAS islet | PAS islet | | | | |
| | | | | | | | DAS K562 | DAS K562 | Smad4 | T | 1.69813421 | JASPAR |
| 82 | rs7776054 | A | G | A | 0.036979 | 0.035704 | DAS HepG2 | DAS HepG2 | | | | |
| | | | | | | | | | SOX7_2 | A | 2.799758628 | ENCODE |
| | | | | | | | | | SOX21_3 | A | 2.677972746 | ENCODE |
| | | | | | | | | | SOX21_3 | A | 2.594721516 | ENCODE |
| | | | | | | | | | SRY_6 | A | 2.46010467 | ENCODE |
| | | | | | | | | | SOX7_2 | A | 2.454474607 | ENCODE |
| | | | | | | | | | SRY_6 | A | 2.443406919 | ENCODE |
| | | | | | | | | | SOX8_2 | A | 2.035455393 | ENCODE |
| | | | | | | | | | SOX9_4 | A | 2.010689954 | ENCODE |
| 83 | rs4709746 | C | T | C | 0.49737 | 0.425999 | | PAS islet | | | | |

| | | | | | | | | | | | | |
|---|---|---|---|---|---|---|---|---|---|---|---|---|
| | | | | | | | | PAS HepG2 | Ptf1a(var.3) | T | 5.861995448 | JASPAR |
| | | | | | | | | | MYOD1_3 | T | 5.578304817 | ENCODE |
| | | | | | | | | | Rbpjl | T | 5.103892788 | JASPAR |
| | | | | | | | | | ZEB2(Zf) | T | 4.756937074 | HOMER |
| | | | | | | | | | ZEB1_known3 | T | 4.382026635 | ENCODE |
| | | | | | | | | | TCF4_1 | T | 3.899068166 | ENCODE |
| | | | | | | | | | TCF3_2 | T | 3.620112597 | ENCODE |
| | | | | | | | | | EGR4_1 | C | 3.547508501 | ENCODE |
| | | | | | | | | | MYF6_1 | T | 3.497001511 | ENCODE |
| | | | | | | | | | SNAI3 | T | 3.491632247 | JASPAR |
| | | | | | | | | | EGR3_1 | C | 3.362597208 | ENCODE |
| | | | | | | | | | ID4_1 | T | 3.23044881 | ENCODE |
| | | | | | | | | | TCF3_1 | T | 3.140914283 | ENCODE |
| | | | | | | | | | Slug(Zf) | T | 3.062275664 | HOMER |
| | | | | | | | | | EGR1_disc1 | C | 2.978844056 | ENCODE |
| | | | | | | | | | Snail1(Zf) | T | 2.967909768 | HOMER |
| | | | | | | | | | SNAI2 | T | 2.863880142 | JASPAR |
| | | | | | | | | | MYOD1 | T | 2.773189503 | JASPAR |
| | | | | | | | | | E2A(bHLH),near_PU.1 | T | 2.691695313 | HOMER |
| | | | | | | | | | EGR4_2 | C | 2.633679658 | ENCODE |
| | | | | | | | | | EGR4 | C | 2.631939862 | JASPAR |
| | | | | | | | | | TBX5_3 | T | 2.591454109 | ENCODE |
| | | | | | | | | | TBX5_4 | T | 2.556389795 | ENCODE |
| | | | | | | | | | TBX5 | T | 2.556389795 | JASPAR |
| | | | | | | | | | EGR1_known10 | C | 2.548810452 | ENCODE |
| | | | | | | | | | EGR3_3 | C | 2.530594834 | ENCODE |
| | | | | | | | | | EGR1_known1 | C | 2.478924578 | ENCODE |
| | | | | | | | | | EGR2 | C | 2.435481766 | JASPAR |
| | | | | | | | | | EGR1_known8 | C | 2.419622138 | ENCODE |
| | | | | | | | | | EGR1_known7 | C | 2.28460259 | ENCODE |
| | | | | | | | | | EGR1_known11 | C | 2.276439813 | ENCODE |
| | | | | | | | | | MYF6_2 | T | 2.273451943 | ENCODE |
| | | | | | | | | | GRHL2 | T | 2.26371094 | JASPAR |
| | | | | | | | | | EGR3_2 | C | 2.112639611 | ENCODE |
| | | | | | | | | | EGR1_known6 | C | 2.085854757 | ENCODE |
| | | | | | | | | | TBX15_2 | T | 2.070552594 | ENCODE |
| | | | | | | | | | EGR1_known2 | C | 2.062612238 | ENCODE |
| 84 | rs13226769 | C | G | G | 0.00487 | 0.006493 | DAS islet | DAS islet | | | | |
| | | | | | | | DAS K562 | DAS K562 | TRIM28_disc2 | C | 3.169803931 | ENCODE |
| | | | | | | | DAS HepG2 | DAS HepG2 | KLF13_1 | C | 2.997555428 | ENCODE |
| | | | | | | | | | ZFP42 | C | 2.38253633 | JASPAR |
| | | | | | | | | | SMAD_1 | C | 2.020464532 | ENCODE |
| 85 | rs17168486 | C | T | T | 0.161831 | 0.168991 | DAS islet | DAS islet | | | | |
| | | | | | | | DAS HepG2 | DAS K562 | RFX3_4 | T | 2.391780672 | ENCODE |
| | | | | | | | | DAS HepG2 | RFX3 | T | 2.313821166 | JASPAR |
| | | | | | | | | | MYC_disc4 | T | 2.268937059 | ENCODE |

| | | | | | | | | | | | | |
|---|---|---|---|---|---|---|---|---|---|---|---|---|
| | | | | | | | | | RFX2_1 | T | 2.248242352 | ENCODE |
| | | | | | | | | | RFX2_1 | T | 2.243195293 | ENCODE |
| | | | | | | | | | RFX2_3 | T | 2.170467128 | ENCODE |
| | | | | | | | | | RFX2_3 | T | 2.132934596 | ENCODE |
| | | | | | | | | | RFX2 | T | 2.069573041 | JASPAR |
| | | | | | | | | | RFX2 | T | 2.062686815 | JASPAR |
| | | | | | | | | | RFX3_2 | T | 2.001244272 | ENCODE |
| 86 | rs12700421 | C | G | C | 0.325961 | 0.27579 | PAS islet | PAS islet | | | | |
| | | | | | | | | | FOXP3_1 | C | 3.111651376 | ENCODE |
| | | | | | | | | | TP63 | G | 2.190265195 | JASPAR |
| | | | | | | | | | TP73_1 | G | 2.127179628 | ENCODE |
| | | | | | | | | | RAD21_disc6 | C | 2.041529496 | ENCODE |
| | | | | | | | | | TP73 | G | 2.008911391 | JASPAR |
| 87 | rs2286177 | T | C | T | 0.370513 | 0.328905 | DAS K562 | PAS islet | | | | |
| | | | | | | | | DAS K562 | NR2F2_1 | C | 2.600419537 | ENCODE |
| | | | | | | | | | RFX3_1 | T | 2.598172345 | ENCODE |
| | | | | | | | | | RFX5_known5 | T | 2.493781254 | ENCODE |
| | | | | | | | | | EGR1_disc4 | C | 2.230553965 | ENCODE |
| | | | | | | | | | RFX1 | T | 2.009952515 | JASPAR |
| 88 | rs2908292 | C | T | C | 0.153968 | 0.129866 | DAS HepG2 | PAS islet | | | | |
| | | | | | | | | PAS K562 | RUNX1_2 | T | 2.257927942 | ENCODE |
| 89 | rs72607746 | C | T | T | 0.123945 | 0.127785 | DAS islet | DAS HepG2 | | | | |
| | | | | | | | DAS K562 | | GFI1B_1 | T | 4.089466504 | ENCODE |
| 90 | rs72638983 | G | T | G | 0.039249 | 0.029665 | DAS HepG2 | PAS islet | | | | |
| | | | | | | | | PAS K562 | IRF_disc1 | G | 3.685112971 | ENCODE |
| | | | | | | | | PAS HepG2 | E2F_disc4 | G | 2.940565892 | ENCODE |
| | | | | | | | | | ZNF423_1 | G | 2.441171256 | ENCODE |
| | | | | | | | | | RFX5_disc2 | G | 2.283936534 | ENCODE |
| | | | | | | | | | CEBPB_disc2 | G | 2.126908079 | ENCODE |
| | | | | | | | | | NFY_known6 | G | 2.106036415 | ENCODE |
| | | | | | | | | | NFY_known3 | G | 2.067937717 | ENCODE |
| 91 | rs4237150 | G | C | C | 0.096983 | 0.149395 | DAS islet | DAS islet | | | | |
| | | | | | | | DAS HepG2 | DAS HepG2 | NR3C1_known7 | G | 3.450522448 | ENCODE |
| | | | | | | | | | ZNF528(Zf) | G | 2.312585176 | HOMER |
| 92 | rs1575972 | T | A | A | 0.225602 | 0.242512 | DAS islet | DAS islet | | | | |
| | | | | | | | DAS HepG2 | DAS HepG2 | EN1_2 | A | 3.461529306 | ENCODE |
| 93 | rs2796441 | G | A | G | 0.206639 | 0.151748 | PAS islet | PAS islet | | | | |
| | | | | | | | DAS K562 | DAS K562 | POU4F2 | A | 3.068115281 | JASPAR |
| | | | | | | | DAS HepG2 | DAS HepG2 | POU4F2_2 | A | 2.886756051 | ENCODE |
| | | | | | | | | | LHX9_3 | A | 2.670806028 | ENCODE |
| | | | | | | | | | HESX1_2 | A | 2.504857436 | ENCODE |
| | | | | | | | | | BCL6B | A | 2.199400857 | JASPAR |
| | | | | | | | | | BCL6B_2 | A | 2.03441786 | ENCODE |
| 94 | rs73554552 | C | A | A | 0.027729 | 0.028216 | DAS islet | PAS K562 | | | | |
| | | | | | | | | DAS islet | FOXA_known2 | C | 2.379450673 | ENCODE |
| | | | | | | | | | ZNF317 | A | 2.371408315 | JASPAR |

**Table S3. Disruption of transcription factor motifs by prioritized SNPs.** Difference in the binding predictions of transcription factors with motifs that overlap prioritized SNPs. Difference measured by the absolute value of the difference in the log of *P*-values for binding at each allele (log difference column). The “high score allele” column refers to the allele with the greatest enhancer probability score from TRENDNet.

| Layer no | Type | Activation | Parameters |
|---|---|---|---|
| 1 | Convolution | ReLU | filters=320, kernel size=8 |
| 2 | Convolution | ReLU | filters=320, kernel size=8 |
| 3 | Dropout | | probability=0.2 |
| 4 | Max Pooling | | pool size=4, stride=4 |
| 5 | Convolution | ReLU | filters=480, kernel size=8 |
| 6 | Convolution | ReLU | filters=480, kernel size=8 |
| 7 | Dropout | | probability=0.2 |
| 8 | Max Pooling | | pool size=4, stride=4 |
| 9 | Convolution | ReLU | filters=640, kernel size=8 |
| 1 | Convolution | ReLU | filters=640, kernel size=8 |
| 11 | Dropout | | probability=0.2 |
| 12 | Dense | | units=100 |
| 13 | Dense | | units=50 |
| 14 | Sigmoid | | |

**Table S4. TREDNet phase one network architecture**. All kernels were subject to *maximum normalization* value of 0.9. Total trained parameters: ∼143 million. Model was trained with *Adadelta* optimizer using *binary cross entropy* as a cost function.

| Layer no | Type | Activation | Parameters |
|---|---|---|---|
| 1 | Convolution | ReLU | filters=64, kernel size=4 |
| 2 | Batch Normalization | | |
| 3 | Max Pooling | | pool size=2, stride=2 |
| 4 | Dropout | | probability=0.4 |
| 5 | Convolution | ReLU | filters=128, kernel size=2 |
| 6 | Dropout | | probability=0.4 |
| 7 | Dense | | units=100 |
| 8 | Dense | | Units=50 |
| 9 | Dense | | units=1 |
| 10 | Sigmoid | | |

**Table S5. TREDNet phase two network architecture**. All kernels were subject to *maximum normalization* value of 0.9. Total trained parameters: $\sim$12 million. Model was trained with *RMSProp* optimizer using *binary cross entropy* as a cost function.

| Layer no | Type | Activation | Parameters |
|---|---|---|---|
| 1 | Convolution | ReLU | filters=256, kernel size=1 |
| 2 | Batch Normalization | | |
| 3 | Dropout | | probability=0.2 |
| 4 | Convolution | ReLU | filters=256, kernel size=1 |
| 5 | Batch Normalization | | |
| 6 | Dropout | | probability=0.5 |
| 7 | Dense | ReLU | units=100 |
| 8 | Dense | | units=1 |
| 9 | Sigmoid | | |

**Table S6. Peak/dip detection network architecture**. All kernels were subject to *maximum normalization* value of 1.0. Model was trained with *ADAM* optimizer using *binary cross entropy* as a cost function.

| Primer Name | Sequence | rsID | Allele | SNP info | Genomic region | Padding (5' and 3') | Strand | Repeat Masking |
|---|---|---|---|---|---|---|---|---|
| rs75638565_C_For | GAGGCTTTTCCGTTCTGGCCT | rs75638565 | C | hg19_dbSnp153 | chr11:14430386-14430406 | 10 | + | none |
| rs75638565_C_Rev | AGGCCAGAACGGAAAAGCCTC | | | | | | | |
| rs75638565_T_For | GAGGCTTTTCTGTTCTGGCCT | | T | | | | | |
| rs75638565_T_Rev | AGGCCAGAACAGAAAAGCCTC | | | | | | | |
| rs75336838_C_For | AATTACATTTCCCACTTTATG | rs75336838 | C | hg19_dbSnp153 | chr11:14576781-14576801 | 10 | + | none |
| rs75336838_C_Rev | CATAAAGTGGGAAATGTAATT | | | | | | | |
| rs75336838_T_For | AATTACATTTTCCACTTTATG | | T | | | | | |
| rs75336838_T_Rev | CATAAAGTGGAAAATGTAATT | | | | | | | |
| rs117720468_G_For | TCCAAGTTTTGGATTCTCTGA | rs117720468 | G | hg19_dbSnp153 | chr11:14731656-14731676 | 10 | + | none |
| rs117720468_G_Rev | TCAGAGAATCCAAAACTTGGA | | | | | | | |
| rs117720468_C_For | TCCAAGTTTTCGATTCTCTGA | | C | | | | | |
| rs117720468_C_Rev | TCAGAGAATCGAAAACTTGGA | | | | | | | |

**Table S7. Probes used for EMSA experiments**. Each forward and reverse oligo for the biotinylated probes were 5' biotinylated.
